# Genetic analyses of common infections in the Avon Longitudinal Study of Parents and Children cohort

**DOI:** 10.1101/2021.06.18.21259147

**Authors:** Amanda H.W. Chong, Ruth E. Mitchell, Gibran Hemani, George Davey Smith, Rebecca C. Richmond, Lavinia Paternoster

**Affiliations:** MRC Integrative Epidemiology Unit, Population Health Sciences, Bristol Medical School, University of Bristol, Bristol, United Kingdom

**Keywords:** Infection, ALSPAC, Genetics, Antibody, HLA

## Abstract

The burden of infections on an individual and public health is profound. Many observational studies have shown a link between infections and the pathogenesis of disease; however a greater understanding of the role of host genetics is essential. Children from the longitudinal birth cohort, the Avon Longitudinal Study of Parents and Children, had 14 antibodies measured in plasma at age 7: Alpha-casein protein, beta-casein protein, cytomegalovirus, Epstein-Barr virus, feline herpes virus, *Helicobacter pylori*, herpes simplex virus 1, influenza virus subtype H1N1, influenza virus subtype H3N2, measles virus, *Saccharomyces cerevisiae*, Theiler’s virus, *Toxoplasma gondii*, and SAG1 protein domain, a surface antigen of *Toxoplasma gondii* measured for greater precision. We performed genome-wide association analyses of antibody levels against 14 infections (N = 357 – 5010) and identified three genome-wide signals (*P* < 5×10^−8^), two associated with measles virus antibodies and one with *Toxoplasma gondii* antibodies. In an association analysis focused on the human leukocyte antigen (HLA) region of the genome, we further detected 15 HLA alleles at a two-digit resolution and 23 HLA alleles at a four-digit resolution associated with five antibodies, with eight HLA alleles associated with Epstein-Barr virus antibodies showing strong evidence of replication in UK Biobank. Our findings provide a useful resource for any future studies using the ALSPAC infection data and contributes to the understanding of the architecture of host genetics related to infections.

## Introduction

The individual and public health burden of infectious diseases can be substantial. Beyond the immediate impact of infections, exposure to infections has been associated with the development of noncommunicable diseases such as cardiovascular disease, cancer, and autoimmune disease. For example, common infections such as Epstein-Barr virus has been implicated with nasopharyngeal carcinoma(1) and multiple sclerosis(2, 3); *Helicobacter pylori* has been linked to myocardial infarction(4, 5) and ischaemic heart disease(6); and cytomegalovirus has been shown to have a role in the development of atherosclerosis(7). To date, examples of large genome-wide association studies (GWAS) of infections have been performed in COVID-19 Host Genetics investigating people with measured SARS-CoV-2 infection(8), 23andMe examining common infections using retrospective self-reporting(9), UK Biobank using serological measurements of antibody response and seropositivity to antigens(10), and similarly in the Rotterdam Study and Study of Health in Pomerania cohorts using serological measures for people infected with *Helicobacter pylori*(11).

In the current study, we investigate the genetic architecture of antibodies against 14 infections in the Avon Longitudinal Study of Parents and Children (ALSPAC) cohort by: (1) Investigating the options for analysing antibody titers to model antibody levels in terms of whether using measures as a continuous variable or thresholding to define infection status is most appropriate; (2) Identifying genome-wide single nucleotide polymorphisms (SNPs) strongly associated with the antibodies; (3) Identifying HLA alleles strongly associated with the antibodies; (4) Assessing consistency of genetic signals associated with the antibodies at different time points in ALSPAC and in an independent cohort (UK Biobank). (5) Evaluating whether any of the genetic signals identified overlap with SARS-CoV-2 infection (COVID-19 Host Genetics Initiative).

## Materials and Methods

### Study sample

Pregnant women resident in Avon, UK with expected dates of delivery 1st April 1991 to 31st December 1992 were invited to take part in the ALSPAC study(12, 13). The initial number of pregnancies enrolled is 14,541 (for these at least one questionnaire has been returned or a “Children in Focus” clinic had been attended by 19/07/99). Of these initial pregnancies, there was a total of 14,676 foetuses, resulting in 14,062 live births and 13,988 children who were alive at one year of age.

When the oldest children were approximately seven years of age, an attempt was made to bolster the initial sample with eligible cases who had failed to join the study originally. As a result, when considering variables collected from the age of seven onwards (and potentially abstracted from obstetric notes) there are data available for more than the 14,541 pregnancies mentioned above. The number of new pregnancies not in the initial sample (known as Phase I enrolment) that are currently represented on the built files and reflecting enrolment status at the age of 24 is 913 (456, 262 and 195 recruited during Phases II, III and IV respectively), resulting in an additional 913 children being enrolled. The phases of enrolment are described in more detail in the cohort profile paper and its update(12, 13). The total sample size for analyses using any data collected after the age of seven is therefore 15,454 pregnancies, resulting in 15,589 foetuses. Of these 14,901 were alive at one year of age.

A 10% sample of the ALSPAC cohort, known as the Children in Focus (CiF) group, attended clinics at the University of Bristol at various time intervals between 4 to 61 months of age. The CiF group were chosen at random from the last 6 months of ALSPAC births (1432 families attended at least one clinic). Excluded were those mothers who had moved out of the area or were lost to follow-up, and those partaking in another study of infant development in Avon.

The study website contains details of all the data that is available through a fully searchable data dictionary and variable search tool (http://www.bristol.ac.uk/alspac/researchers/our-data/). Ethical approval for the study was obtained from the ALSPAC Ethics and Law Committee and the Local Research Ethics Committees. Consent for biological samples has been collected in accordance with the Human Tissue Act (2004).

### Phenotype measurement

ALSPAC children were invited to participate in clinical assessments at around seven years of age (“Focus @ 7”) between September 1998 and October 2000(14). Attendees at this clinic were invited to give blood samples. Additional blood samples were also available from the “Children in Focus” (CiF) group, a 10% randomly selected subset of ALSPAC children at around five years of age between September 1998 and October 2000, as well as “Focus @ 11+” at 11 years collected between January 2003 and January 2005, and “TeenFocus 3” at 15 years collected between October 2006 and November 2008(14). Antibodies against 14 infections were assessed at these four clinical time points.

Whole blood samples were collected and processed by centrifugation at 3500rpm, for 10 minutes at 4-5°C (14). Subsequently, the plasma fraction from the whole blood was aliquoted out and temporarily stored at −20°C before being stored long term at −70/80°C. In preparation for analysis, EDTA plasma samples were plated out into 96-well plates. Enzyme-linked immunosorbent assay (ELISA) was performed using a specific antigen for each infection of interest to measure IgG antibody titers or IgA antibody titers, in the case for *Sacchraromyces cerevisiae* (14). Briefly, this method involved microtiter plates coated with antigens reacting with a sequence of diluted human EDTA plasma, enzyme labelled anti-human IgG and enzyme substrate, with a plate wash separating each reaction step. Next, a microplate colourimeter was used to measure the optical density of enzyme-substrate reactions.

The antibodies examined in this analysis are against the following antigens: *Toxoplasma gondii* (*T*.*gondii*), surface antigen 1 (SAG1) protein domain of *T*.*gondii*, cytomegalovirus (CMV), Epstein-Barr virus (EBV), herpes simplex virus type 1 (HSV1), influenza virus subtypes H1N1 and H3N2, measles virus, *Saccharomyces cerevisiae* (*S*.*cerevisiae*), *Helicobacter pylori* (*H*.*pylori*), feline herpes virus (FHV), Theiler’s virus (TV), bovine casein alpha protein (alpha-casein) and beta protein (beta-casein).

For each antibody, measurements of optical density were read directly from the ELISA plate. The ratio to standards were then derived from the standards measured on each plate. These ratios were then standardised to produce z-scores with a mean of two and a standard deviation of one per plate (Ratio to standard minus the mean ratio to standard then divided by the standard deviation per plate, plus two) (14). In addition, the data were further transformed using the rank-based inverse normal approach to generate normal distributions.

Not all participants were measured for all 14 antibodies at every clinical time point. The antibody titers for each antigen at the seven-year time point were analysed in the primary analysis as they represented the largest sample size, and the distribution of z-scores for each infection did not largely differ across time points.

### Genotyping and imputation

ALSPAC children were genotyped by 23andMe using the Illumina HumanHap550 quad chip genome-wide SNP genotyping (Illumina, Inc., San Diego, CA) subcontracting from the Wellcome Trust Sanger Institute, Cambridge, UK and the Laboratory Corporation of America, Burlington, NC, US. Genotypes were called with Illumina GenomeStudio and PLINK(v1.07) (15) was used to perform quality control measures. These measures were performed on an initial set of 9,912 individuals, including children who participated at the Focus @ 7 clinic, and 609,203 directly genotyped SNPs. Individuals were removed from further analysis if they had extreme autosomal heterozygosity, >3% missingness, undetermined X chromosome heterozygosity, and insufficient sample replication (<0.8 identity-by-descent (IBD)). In addition, population stratification was assessed using multidimensional scaling of genome-wide identity-by-state pairwise distances using HapMap v2 (release 22) European (CEU), Han Chinese (CHB), Japanese (JPT) and Yoruba (YRI) populations as references, with individuals with non-European ancestry excluded. SNPs were removed if they had a minor allele frequency less than 1%, a call rate of <95%, or displayed a Hardy-Weinberg equilibrium P value of less than 5×10^−7^. Cryptic relatedness was assessed as the proportion of identity by descent (IBD >0.1) described previously (16).

After quality control steps, a total of 9,115 individuals and 500,527 SNPs passed these filters. ALSPAC children were phased using ShapeIt v2(17) to phase the HRC panel (39,235,157 SNPs). Genotype imputation was performed with Michigan Imputation Server(18) using the Haplotype Reference Consortium (HRCr1.1) panel of approximately 31,00 phased whole genomes.

Two software packages, HLA*IMP:03(19) and SNP2HLA(20), were used to impute classical human leukocyte antigens for within the major histocompatibility complex (MHC) located on chromosome six with a range of approximately 4 Mb. A comparison of the imputations was performed by comparing the allele dosages obtained for classical two- and four-digit resolution HLA alleles (for HLA-A, HLA-B, HLA-C, HLA-DPB1, HLA-DRB1) and subsequent association analyses using antibodies measured in ALSPAC children.

SNP2HLA_package_v1.0.3(20) was utilised and software dependencies Beagle (version 3.0.4), PLINK (v1.07)(15) and beagle2linkage.jar(20) were downloaded. Imputation was performed using the pre-constructed Type 1 Diabetes Genetics Consortium (T1DGC) reference panel which consists of 5225 unrelated individuals of European ancestry, and we imputed 126 classical 2-digit HLA alleles, 298 classical four-digit HLA alleles (at HLA-A, HLA-B, HLA-C, HLA-DPA1, HLA-DPB1, HLA-DQA1, HLA-DQB1 and HLA-DRB1), 176 HLA insertions/deletions, 1,101 HLA intragenic SNPs and 399 HLA amino acids(20). The SNP2HLA output then converted the posterior probabilities for each best-imputed allele to a dosage(20).

For HLA*IMP:03 (v.0.1.0)(19), HLA imputation was performed using its online imputation service. Prior to upload into the automated online imputation service, ALSPAC genotype data was converted to phased haplotype files in five batches due to memory size limitations. Outputs from the online service were converted into a dosage format, similar to the SNP2HLA dosage output, for association testing by calculating the expected dosage of each allele. This was performed on each individual by summing the two posterior probabilities calculated for a given classical allele.

Concordance of HLA imputation using the two packages were compared by assessing the difference in dosages by providing a call threshold, *T*, to determine concordance (−0.5 ≤ *T* ≤ 0.5) or discordance (−0.5 ≥ *T* ≥ 0.5) between the two approaches. The proportion of concordant and discordant HLA alleles at each given HLA loci were then tabulated. In addition, allele frequencies for each HLA loci were calculated for each HLA imputation approach, and values were compared to the T1DGC reference panel.

### Statistical analysis

#### Comparison of treating antibody titer data as continuous or thresholded variables

It would be desirable to be able to stratify individuals into seropositive and seronegative for each antibody analysed. However, determining where to call the threshold for continuous antibody data is complex due to the unavailability of reliable assays on reliably determined controls to establish uninfected population baseline distributions (Figure 1). For example, cases could be defined as: (1) Infected, if it is assumed that everyone who is exposed produces an antibody response; (2) High responders, which includes only a subset of people exposed, and controls are a mixture of low responders and unexposed; (3) A mixture of exposed and unexposed individuals if background levels of unexposed vary substantially. We sought to investigate this challenge by examining whether we could estimate thresholds that best represented seropositivity using several approaches, (1) Ratio to antigen/plate standards ≥ 1, as suggested by Barnes et al 2015 (21); (2) A subjective threshold based on the selecting the trough between two obvious peaks where observed for the ratio to standard distributions; and two methods that used the results of genetic association analyses to determine the best threshold: (3) The threshold that exhibited the most signal for each antibody (as assessed by GWAS Q-Q plots of seven arbitrary thresholds: 5% cases 95% control; 10% cases 90% control; 25% cases 75% control; 50% cases 50% control; 75% cases 25% control; 90% cases 10% control; 95% cases 5% control), and (4) The threshold that best recapitulated GWAS results (at a suggestive p-value of 1×10^−6^) from an analysis where the antibody measures were treated as a continuous variable. (NB - This method was selected in place of performing genetic correlation analyses as genetic correlations could not distinguish between thresholds, and many antibodies lacked the sample size requirements to perform the analyses).

**Figure 1.**
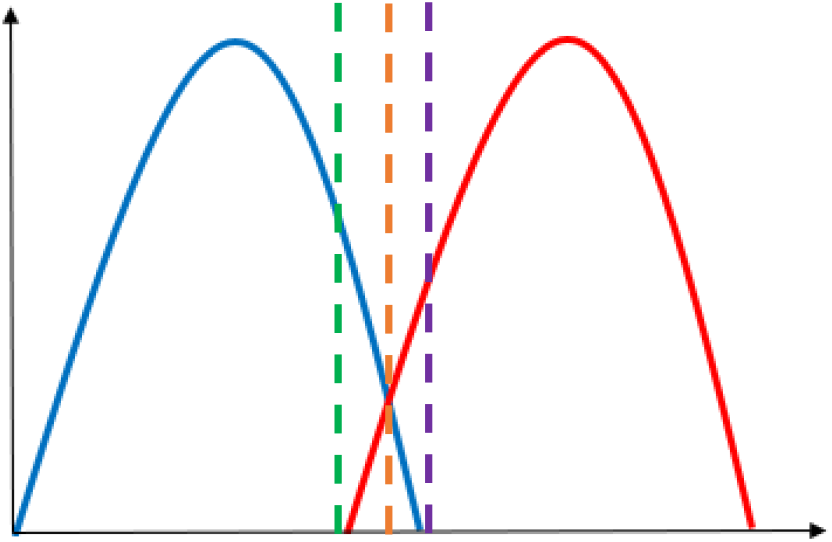
The challenge of thresholding continuous antibody measures. Blue represents the distribution of uninfected individuals; Red represents the distribution of infected individuals. The green, orange, and purple dashed lines represent cut-off points. This figure depicts the theoretical overlap in antibody distribution of uninfected individuals (represented in blue) and infected individuals (represented in red), and the coloured dashed lines denotes how cut-offs may be selected. For example, the green threshold would include all infected individuals as cases, but also include a proportion of uninfected individuals as cases. Alternatively, the purple threshold would result in no uninfected individuals included as cases, but miss some that are infected. The orange threshold may be ideal as it would minimise the proportion of individuals misclassified as either cases or controls. However the true underlying two distributions are not separately observed and so can only ever be estimated from the overall distribution.

### Association testing

Genome-wide association analyses were performed using PLINK (v2.0) (www.cog-genomics.org/plink/2.0/)(21) for each of the 14 ALSPAC antibodies using rank-based inverse normal transformed continuous z-score measures at age seven. In addition, logistic regression was performed applying the various thresholds previously described, for the 14 ALSPAC antibodies at age seven. In addition, linear regression was implemented using PLINK (v2.0) (www.cog-genomics.org/plink/2.0/)(21) on all four ALSPAC clinical time points for all measured antibodies, and these continuous z-score measures were transformed using rank-based inverse normal transformation to make them comparable. Furthermore, stratified association analyses was performed based on measles, mumps and rubella vaccination status using antibodies against measles virus of children at the seven-year clinic. All analyses were carried out using unrelated individuals (IBD < 0.1, corresponds to a relatedness at a first-cousin level) that had available phenotypic and genetic data. Analyses were adjusted for age, sex and the first 10 principal components, and assumed an additive model for genetic effects. SNPs were removed from analyses if they had a minor allele frequency less than 1% and imputation quality (INFO) score of less than 0.8. Genome-wide results were clumped to identify independent loci (using an r2 threshold of 0.1 and a distance of 250 kB in PLINK’s clumping procedure). Genome-wide significance was considered to be a threshold of *P* < 5 × 10^−8^, and we also considered a suggestive threshold of *P* < 1 × 10^−6^.

Association testing of the HLA alleles were performed using the dosage outputs from both imputation methods, HLA*IMP:03(19) and SNP2HLA(20), for each of the 14 antibodies measured in ALSPAC also using the rank-based inverse normal continuous z-score variables from age 7. To aid comparison of methods, classical HLA alleles were only included with the binary markers indicating the presence of the HLA allele or absence of the HLA allele. Probabilistic genotypes were used for each marker to account for uncertainty in imputation. Linear regression analyses was performed using PLINK (v1.90) (www.cog-genomics.org/plink/1.9/)(21) and adjusted for age, sex and the first 10 principal components. To calculate the HLA-wide multiple-testing correction threshold, the Type 1 Diabetes Genetics Consortium (N = 5,225) genetic data was used to produce a correlation matrix of all HLA alleles and MatSpDlite (http://neurogenetics.qimrberghofer.edu.au/matSpDlite/) (22) was used to determine the effective number of HLA alleles. Furthermore, Equation five from Li and Ji et al. 2005 (23) was employed to determine the HLA-wide correction threshold required to keep the Type 1 error rate at 5%.

#### UK Biobank

Comprehensive description of UK Biobank’s cohort profile, serological measures and analyses performed are available in Supplementary Materials. In brief, antibodies against five infections were available for replication: Cytomegalovirus, Epstein-Barr virus, herpes simplex virus 1, *Helicobacter pylori*, and *Toxoplasma gondii*. Analyses were performed using the initial assessment time point for all antigens (N = 9,430). Antibody measurements were rank-based inverse normal transformed for all GWAS and HLA association analyses and beta estimates presented are on this scale. To give an idea of the magnitude of these effects, the main results are also presented using the z-scores of the ratio-to-standard measures, where beta estimates are a change in standard deviation per allele.

#### COVID-19 Host Genetics Initiative

To investigate genetic overlap, ALSPAC antibody association results and the SARS-CoV-2 GWAS meta-analysis findings (C2_ALL_eur) (N = 1,683,784)(8) were compared by performing a look-up of genome-wide and suggestive SNPs from ALSPAC, and genome-wide SNPs from the SARS-CoV-2 GWAS meta-analyses. As the association analyses are presented on different scales, only direction of association and *P-*values were compared.

## Results

### Descriptive characteristics

Distribution of the ratio to standards of the 14 antibodies are shown in Figure 2. Six of the 14 infections had a sample size >1000 individuals (Cytomegalovirus, Epstein-Barr virus, feline herpes virus, *Helicobacter pylori, Toxoplasma gondii*, SAG1 protein domain), and eight had <700 individuals (Alpha-casein protein, Beta-casein protein, herpes simplex virus 1, influenza virus subtype H1N1, influenza virus subtype H3N2, measles virus, *Saccharomyces cerevisiae*) (Table 1). Differences in sample size were because not all individuals were measured for all 14 antibodies. Two of the 14 antibodies showed bimodal z-score distributions (Cytomegalovirus and Epstein-Barr virus, Figure 2), and all antibody distributions showed a strong positive skew. Participants were on average 90 months old (Range: 82 – 113 months) when measures were taken, and approximately half (47-52% dependent on infection) were female(14).

**Table 1.** Suggested antibody thresholds based on percentage of cases using ALSPAC seven-year clinic data compared to published seroprevalence in the United Kingdom. Note:* Only three cut-offs used in selected infections due to smaller sample sizes; ** Published prevalence based on women of child-bearing age; *** No published UK seroprevalence; Seroprevalence percentage in the UK is among individuals of European ancestry and were obtained from different studies with cohorts of ranging ages and data collection time points. Abbreviations: NA, Not applicable due to unimodal distribution or no clumped SNPs that met the suggestive threshold.

| **Antibody (N)** | **Published seroprevalence in UK (%)** | **Ratio to standard ($\geq 1$ seropositive)** | **Subjective threshold based trough in bimodal z-score distribution** | **Threshold with most signal in GWAS (cases %)** | **Threshold that best recapitulated continuous GWAS results (cases %)** |
| --- | --- | --- | --- | --- | --- |
| CMV (5010) | 40-50% (24) | 23.9% | 17% | 25-95 | 50 |
| *H. pylori* (4683) | 27% (25) | 83.7% | NA | 50-95 | 50 |
| *T. gondii* (5010) | <10% (26) ** | 100% | NA | Continuous | 50 |
| EBV (5010) | 85.3% (27) | 46.1% | 36% | 10 | 5-50 |
| FHV (2764) | *** | 65.2% | NA | 5 | 25 |
| SAG1* (1228) | <10% (26) | 100% | NA | 75 | Continuous |
| HSV1* (683) | 61% (28) | 8.3% | NA | Continuous | 25 |
| H1N1* (683) | 0.5-2.2% (29) | 34% | NA | 5 | 25-50 |
| H3N2* (683) | *** | 31.6% | NA | Continuous | 25-75 |
| Measles* (683) | <1%(30) | 82.1% | NA | Continuous | 25-50 |
| Theiler's virus* (683) | *** | 41% | NA | 75 | NA |
| Alpha casein* (683) | *** | 51.2% | NA | Continuous | NA |
| Beta casein* (683) | *** | 47% | NA | Continuous | 25 |
| *S.cerevisiae* * (357) | *** | >1% | NA | Continuous | Continuous |

**Figure 2.**
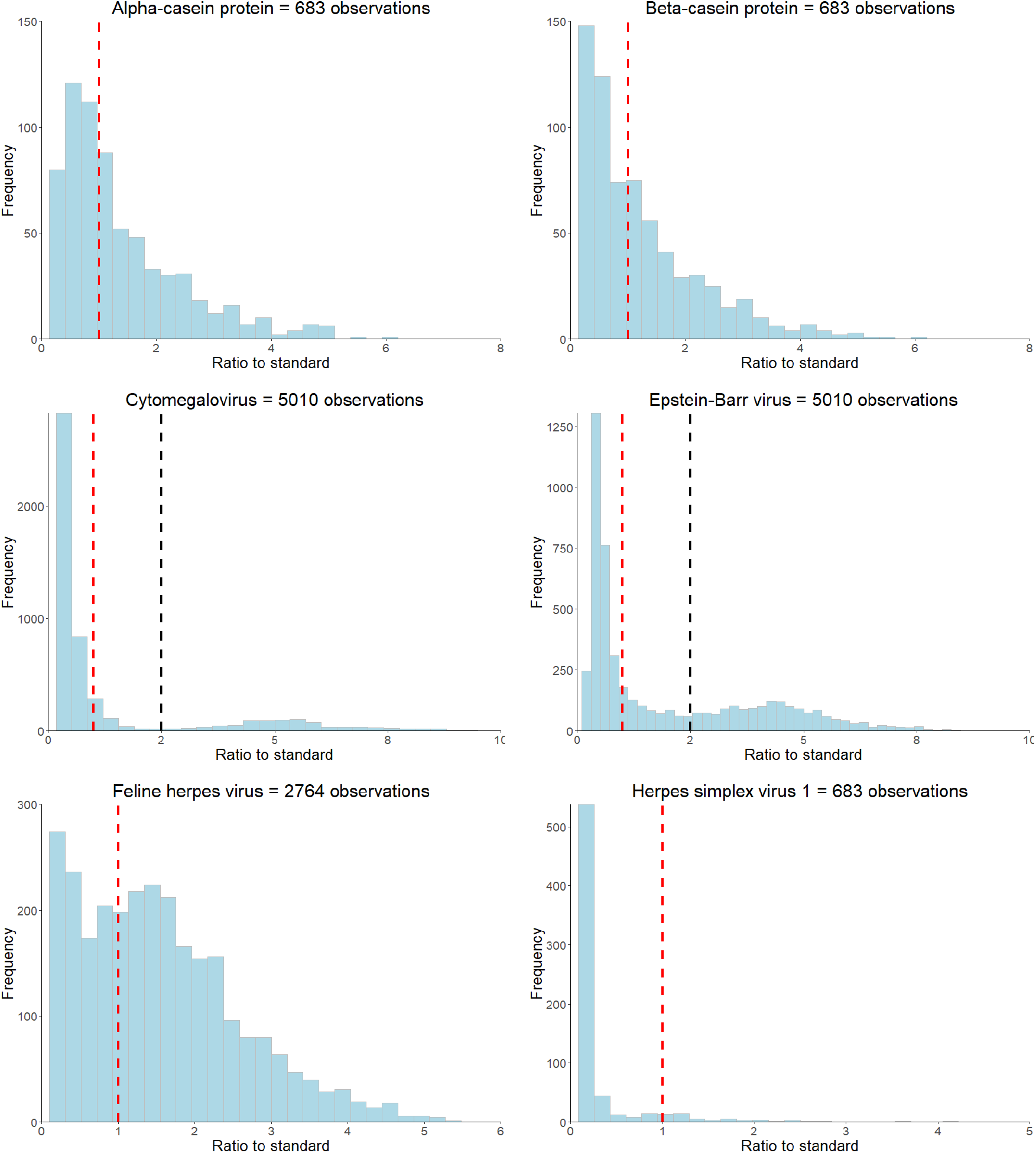
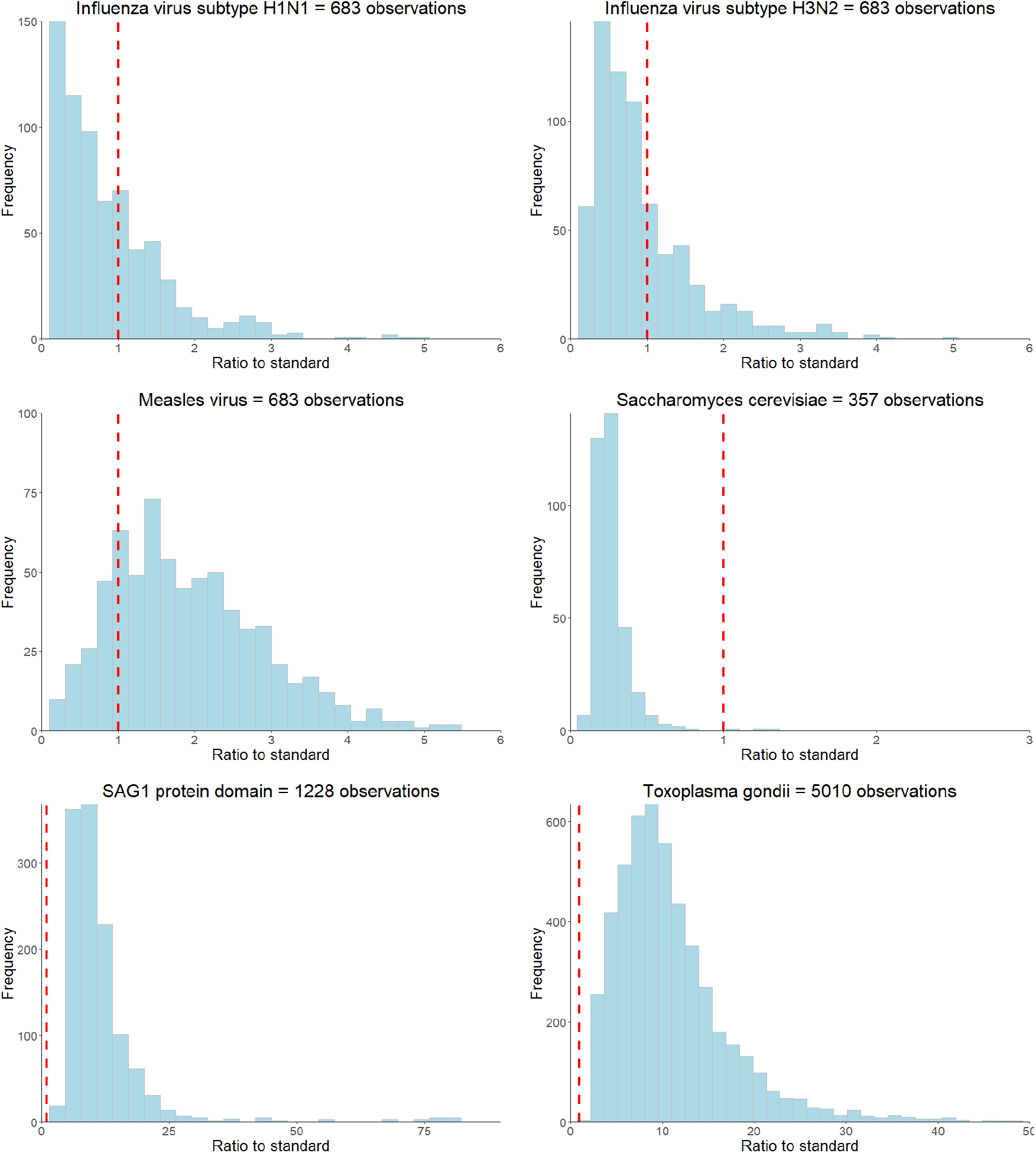
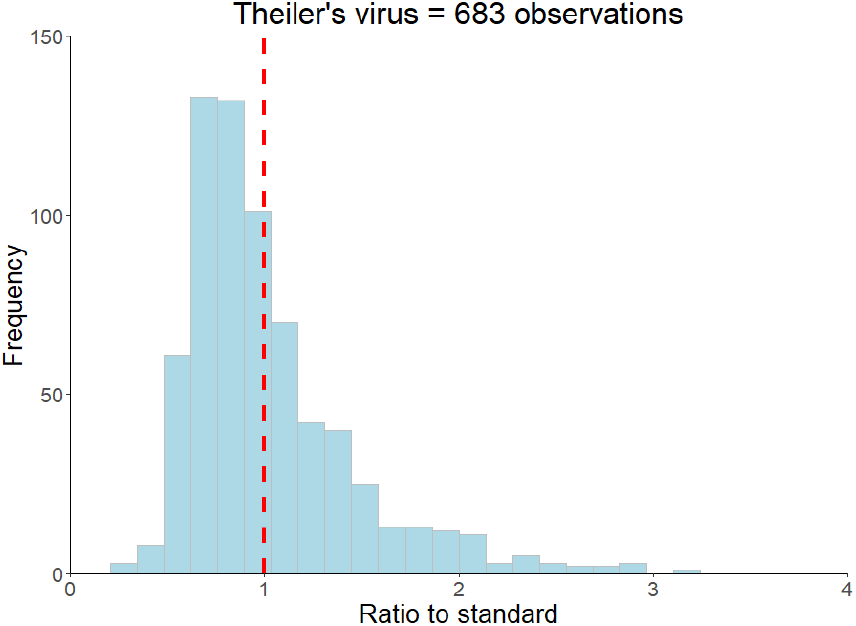
Ratio to standard distributions of the antibodies measured at the ALSPAC seven-year clinic. The red dashed line represents the ratio to standard threshold of >=1 and the black dashed line represents the subjective threshold based on visually identifying the trough in distributions for bimodal antibodies.

### Antibody thresholds

We explored the utility of analysing the 14 antibodies as continuous or thresholded variables in ALSPAC children. Figure 2 and Table 1 shows summary results of each of the tested thresholding approaches (comprehensive results shown in Supplementary Table 1 and Supplementary Figure 1).

Four threshold approaches (as described in the methods) were used: (1) Ratio to standards greater than or equal to 1; (2) A subjective threshold based visually identifying the trough in distributions that were clearly bimodal; (3) The threshold which demonstrated the most signal in a GWAS analysis; (4) The threshold that best recapitulated continuous GWAS results. These were also compared to proportions of seroprevalence measures reported in the literature (Table 1).

Thresholds suggested by the four methods were very inconsistent for all antibody measures and the proportions of seropositive individuals according to the ratio to standards thresholds were not comparable to published seroprevalences in the United Kingdom for each infection (Table 1). Only two measured antibodies had obvious bimodal distributions (Cytomegalovirus and Epstein-Barr virus, Table 1 and Figure 2), but the suggested antibody thresholds were lower than published United Kingdom seroprevalences (17% versus 40-50% for cytomegalovirus, and 36% versus 85% for Epstein-Barr virus), and lower than the ratio to standards approach (Table 1). Seven of the 14 antibodies had most GWAS signals with the rank-based inverse normal transformed continuous measures, as opposed to any of the thresholds (Table 1 and Supplementary Figure 1). Where a thresholded GWAS gave more signal, the suggested thresholds were again inconsistent with the other approaches and published seroprevalences (Table 1 and Supplementary Figure 1).

We compared the top SNP associations (*P* < 1.0×10^−6^) from the continuous GWASs across the thresholded GWASs for each specific antibody, to see if any thresholds gave similar association results on an individual locus basis. The thresholds which gave the most consistent results (to continuous) are shown in Table 1. Again, the suggested thresholds were rarely consistent with any of the other thresholding approaches. The inconsistencies observed between the various methods for selecting a threshold, and the fact that most had more genetic signal when a continuous measure was used, suggest that selecting a threshold to define seropositivity might not be the best way to analyse the data. Therefore, in subsequent analyses we use antibody measures as continuous variables.

### Genome-wide association analyses of antigen-specific antibodies

We performed genome-wide association analyses of all continuous antibody measures at the seven-year clinic in ALSPAC and identified a total of three SNPs with strong associations (*P* < 5 × 10^−8^) to two antibodies (Measles virus or *T*.*gondii*) and 26 suggestive SNPs (*P* < 1 × 10^−6^) associated with 12 antibodies. These SNPs are presented in Table 2 and Table 3, respectively, with Manhattan plots and Q-Q plots illustrated in Supplementary Figure 1 and Figure 2. Furthermore, one association, between rs36020612 and the *T*.*gondii* antibody, was retained after adjusting for the 14 antibodies tested (*P* < 3.6×10^−9^).

**Table 2.** Associations of top SNPs with *P <* 5×10^−8^ measured in the ALSPAC seven-year clinic. Abbreviations: Chr, Chromosome; EA, Effect allele; Beta, Beta estimate for the presence allele; L95, lower 95% confidence interval; U95, Upper 95% confidence interval; P, *p*-value.

| Antibodies (N) | Chr | Position | SNP | EA | Beta | L95 | U95 | P | Gene: Consequence |
| --- | --- | --- | --- | --- | --- | --- | --- | --- | --- |
| Measles (683) | 2 | 151443503 | rs28617484 | C | -0.511 | -0.69 | -0.332 | $3.5 \times 10^{-8}$ | Between *LINC01920* and *LINC02612*: Intergenic |
| | 13 | 78052775 | rs506576 | T | 0.49 | 0.325 | 0.655 | $1.0 \times 10^{-8}$ | Between *LOC105370271* and *SCEL*: Intergenic |
| *T.gondii* (5010) | 11 | 123344435 | rs36020612 | T | 0.202 | 0.147 | 0.258 | $1.2 \times 10^{-12}$ | *GRAMD1B*: Intronic |

**Table 3.** Suggestive SNPs with *P* < 1 × 10^−6^ in measured in the ALSPAC seven-year clinic. Abbreviations: Chr, Chromosome; EA, Effect allele; Beta, Beta estimate for the presence allele; L95, lower 95% confidence interval; U95, Upper 95% confidence interval; P, *p*-value.

| Antibodies (N) | Chr | Position | SNP | EA | Beta | L95 | U95 | P | Gene: Consequence |
| --- | --- | --- | --- | --- | --- | --- | --- | --- | --- |
| Beta-casein (683) | 2 | 126266581 | rs113530838 | T | 0.911 | 0.562 | 1.260 | $4.3 \times 10^{-7}$ | Between *RNU6-259P* and *LOC105373598*: Intergenic |
| | 6 | 128237257 | rs145993168 | A | 1.245 | 0.755 | 1.736 | $8.8 \times 10^{-7}$ | *THEMIS*: Intronic |
| CMV (5010) | 4 | 114752126 | rs10028968 | A | -0.112 | -0.157 | -0.067 | $9.5 \times 10^{-7}$ | Between *CAMK2D* and *LOC105377375*: Intergenic |
| EBV (5010) | 6 | 32192331 | rs3096702 | G | -0.116 | -0.161 | -0.071 | $4.3 \times 10^{-7}$ | *NOTCH4*: 2KB upstream |
| | 7 | 40289816 | rs186721582 | A | 0.429 | 0.264 | 0.594 | $3.6 \times 10^{-7}$ | *SUGCT*: Intronic; *LOC105375245*: Intronic |
| | 10 | 62076002 | rs74807157 | A | 0.269 | 0.164 | 0.375 | $6.2 \times 10^{-7}$ | *ANK3*: Intronic |
| | 15 | 91086484 | rs6416551 | G | -0.210 | -0.286 | -0.134 | $7.3 \times 10^{-8}$ | *CRTC3*: Intronic |
| FHV (2764) | 6 | 32602482 | rs3104369 | C | -0.198 | -0.272 | -0.124 | $2.0 \times 10^{-7}$ | Between *HLA-DRB1* and *LOC107986589*: Intergenic |
|  | 12 | 129882215 | rs117760947 | A | 0.484 | 0.297 | 0.672 | 4.4×10<sup>-7</sup> | *TMEM132D*: Intronic |
|  | 19 | 48319485 | rs6509344 | G | -0.158 | -0.218 | -0.097 | 3.6×10<sup>-7</sup> | Between *TPRX1* and *CRX*: Intergenic |
| *H.pylori* (4683) | 6 | 32672903 | rs35030589 | A | -0.175 | -0.242 | -0.108 | 3.4×10<sup>-7</sup> | Between *HLA-DQB1* and *MTCO3P1*: Intergenic |
| HSV1 (683) | 2 | 121207018 | rs6758808 | T | -0.403 | -0.556 | -0.249 | 3.8×10<sup>-7</sup> | Between *INHBB* and *LINC01101*: Intergenic |
|  | 3 | 52120118 | rs140033700 | A | 1.335 | 0.826 | 1.843 | 3.9×10<sup>-7</sup> | *POC1A*: Intronic |
|  | 9 | 15248194 | rs10961934 | G | 0.533 | 0.326 | 0.741 | 6.4×10<sup>-7</sup> | *TTC39B*: Intronic |
| H1N1 (683) | 4 | 183847475 | rs60526956 | T | 0.376 | 0.232 | 0.520 | 4.3×10<sup>-7</sup> | Between *DCTD* and *LOC105377577*: Intergenic |
|  | 19 | 52351623 | rs7256238 | T | 0.404 | 0.245 | 0.564 | 9.0×10<sup>-7</sup> | *ZNF577*: Intronic |
| H3N2 (683) | 2 | 33610710 | rs58804388 | A | -0.420 | -0.577 | -0.263 | 2.4×10<sup>-7</sup> | *LTBPI*: Intronic |
|  | 10 | 71258686 | rs60590291 | T | -0.590 | -0.820 | -0.360 | 6.7×10<sup>-7</sup> | *TSPAN15*: Intronic |
| Measles (683) | 3 | 190482826 | rs116249622 | T | 0.980 | 0.593 | 1.368 | 9.3×10<sup>-7</sup> | Between *CCT6P4* and *LOC100131685*: Intergenic |
|  | 4 | 149791968 | rs1634083 | G | 0.328 | 0.207 | 0.450 | 1.7×10<sup>-7</sup> | *LOC107986195*: Intronic |
| *S.cerevisiae* (357) | 4 | 41671067 | rs140144775 | A | -1.905 | -2.651 | -1.160 | 9.8×10<sup>-7</sup> | *LIMCH1*: Intronic |
|  | 17 | 7366619 | rs34914463 | C | -0.655 | -0.902 | -0.407 | 4.3×10<sup>-7</sup> | *ZBTB4*: Missense |
| SAG1 (1228) | 10 | 743730 | rs117614721 | G | -0.748 | -1.038 | -0.457 | 5.4×10<sup>-7</sup> | Between *DIP2C* and *LARP4B*: Intergenic |
| *T.gondii* (5010) | 1 | 48630438 | rs10788882 | C | -0.116 | -0.162 | -0.070 | 8.8×10<sup>-7</sup> | *LINC02794*: Intronic; *SKINTIL*: Non-coding transcript variant |
|  | 1 | 161500796 | rs72717048 | G | -0.219 | -0.302 | -0.135 | 2.7×10<sup>-7</sup> | Between *TRL-CAG1-6* and *TRG-TCC2-6*: Intergenic |
|  | 3 | 98620083 | rs190637453 | C | 0.530 | 0.324 | 0.736 | 4.8×10<sup>-7</sup> | *DCBLD2*: Missense; *LOC105374002*: 2KB upstream |

The strongest association for measles virus was rs506576 (EA = T, *P* = 1.0×10^−8^, β = 0.49, 95% CI = 0.33, 0.66), an intergenic SNP located between *LOC105370271* and *SCEL* on chromosome 13q22.3. Followed by rs28617484 (EA = C, *P* = 3.5×10^−8^, β = −0.51, 95% CI = −0.69, −0.33), also an intergenic SNP, between *LINC01920* and *LINC02612* on chromosome 2q23.3. In addition, stratified analysis by measles, mumps and rubella (MMR) vaccination status was performed on the genome-wide and suggestive SNPs. Findings in Table 4 demonstrated that children that had been vaccinated did not contribute largely to the genetic signals identified (*P* > 0.001), with environmental exposure of the measles virus showing greater genetic signal (*P* > 2.6×10^−6^).

**Table 4.** Stratified analyses for antibodies against measles virus at the seven-year clinic by measles, mumps, and rubella (MMR) vaccine status. Abbreviations: Chr, Chromosome; EA, Effect allele; Beta, Beta estimate for the presence allele; L95, lower 95% confidence interval; U95, Upper 95% confidence interval; P, *p*-value.

| Antibody | MMR status (N) | Chr | Position | SNP | EA | Beta | L95 | U95 | P |
| --- | --- | --- | --- | --- | --- | --- | --- | --- | --- |
| Measles | Received (252) | 2 | 151443503 | rs28617484 | C | -0.512 | -0.841 | -0.182 | 0.003 |
|  |  | 3 | 190482826 | rs116249622 | T | 1.088 | 0.216 | 1.960 | 0.015 |
|  |  | 4 | 149791968 | rs1634083 | G | 0.290 | 0.093 | 0.487 | 0.004 |
|  |  | 13 | 78052775 | rs506576 | T | 0.459 | 0.189 | 0.728 | 0.001 |
| | Unknown (430) | 2 | 151443503 | rs28617484 | C | -0.502 | -0.724 | -0.281 | $1.2 \times 10^{-5}$ |
| | | 3 | 190482826 | rs116249622 | T | 0.991 | 0.546 | 1.436 | $1.7 \times 10^{-5}$ |
| | | 4 | 149791968 | rs1634083 | G | 0.387 | 0.229 | 0.546 | $2.6 \times 10^{-6}$ |
| | | 13 | 78052775 | rs506576 | T | 0.514 | 0.300 | 0.728 | $3.7 \times 10^{-6}$ |

For *T*.*gondii*, one intronic SNP was identified, rs36020612 (EA = T, *P* = 1.2×10^−12^, β = 0.20, 95% CI = 0.15, 0.26) located in *GRAM Domain Containing 1B* (*GRAMD1B*) on 11q24.1 (Table 2). Expression of this gene has previously been associated with lymphocyte traits, leukaemia, and eosinophil percentage(31-33). A look-up of rs36020612 was performed using the SAG1 protein domain antibody GWAS, as SAG1 protein domain is an antigen of *T*.*gondii* measured to provide greater precision (EA = T, *P* = 0.78, β = −0.02, 95% CI = −0.14, 0.10). No evidence of an association was identified, with effect size shown in the opposite direction.

We then assessed the consistency of genetic signals from the seven-year clinic at the five-year clinic, 11-year clinic, and the 15-year clinic time points. In summary, six SNPs showed good evidence for association at other timepoints (*P* < 4.3 ×10^−4^, adjusted for the three additional timepoints and the 29 associations tested), with all estimates showing consistency in the direction of effects (Table 5, with comprehensive results of all antibodies shown in Supplementary Table 2).

**Table 5.**
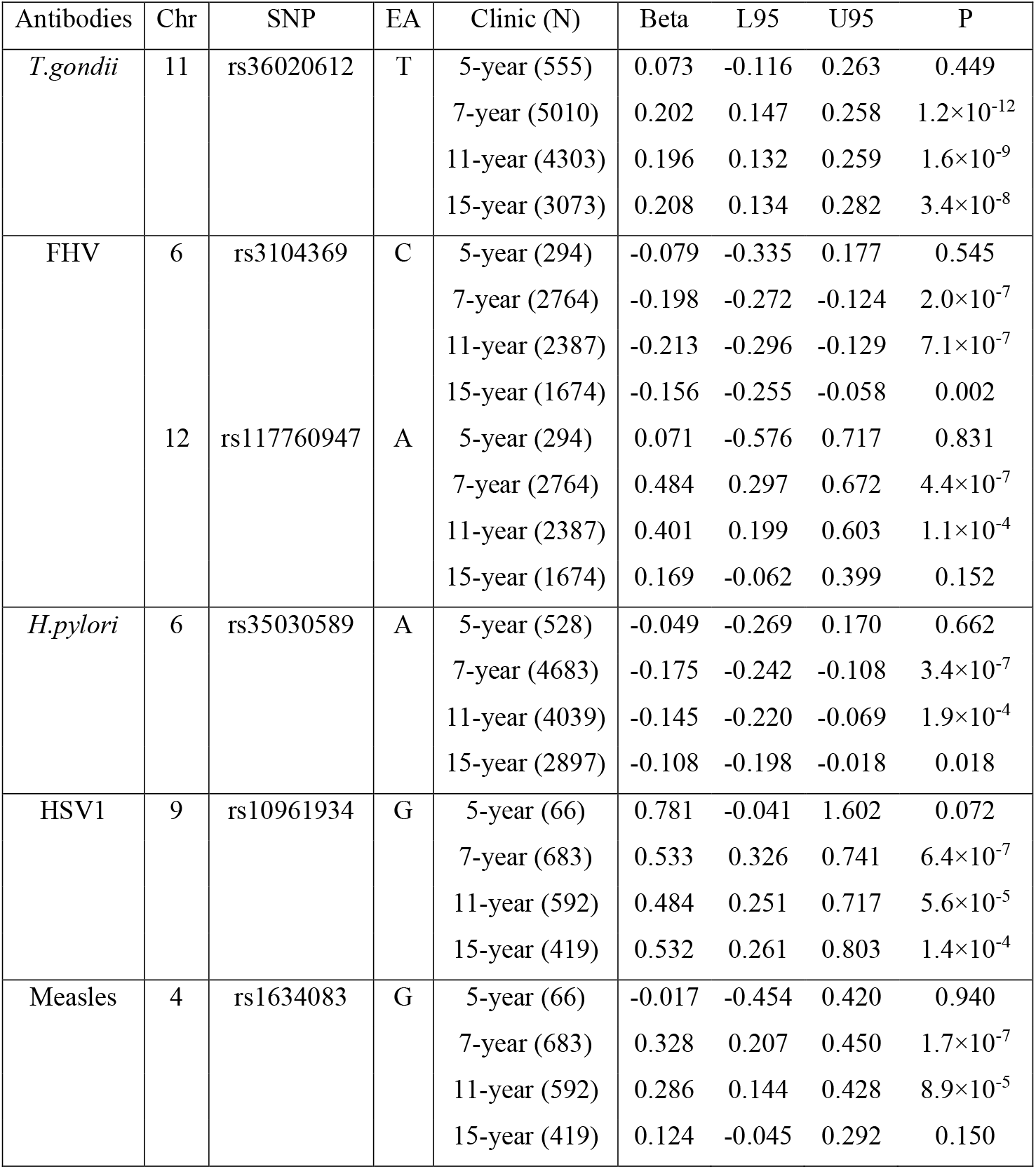
Associations identified using ALSPAC seven-year clinic antibody data (P < 1.0×10^−6^) that also showed evidence for association (*P* < 4.3×10^−4^) at other timepoints. Abbreviations: Chr, Chromosome; EA, Effect allele; Beta, Beta estimate for the presence allele; L95, lower 95% confidence interval; U95, Upper 95% confidence interval; P, *p*-value.

The strongest associations at different time points were demonstrated between rs36020612 (*GRAMD1B*) and antibodies against *T*.*gondii*, with effect sizes similar to that observed at the 7-year clinic (β = 0.20) also seen at the 11-year clinic (β = 0.20, *P* = 1.6 × 10^−9^) and the 15-year clinic (β = 0.21, *P* = 3.4 × 10^−8^), but this association was not seen at the earlier five-year clinic (*P =* 0.45).

For antibodies against feline herpes virus, two SNPs replicated at the 11-year clinic time point: rs3104369 (β = −0.21, *P* = 7.1 × 10^−7^); rs117760947 (β = 0.40, *P* = 1.1 × 10^−4^). These effect sizes were similar to the effect sizes exhibited at the seven-year clinic: rs3104369 (β = −0.19); rs117760947 (β = 0.48), however these associations were not observed at the five-year and 15-year clinic (*P* < 0.83).

Furthermore, rs35030589 was shown to be associated with antibodies against *H*.*pylori* at the seven-year clinic (β = −0.17) and the eleven-year clinic (β = −0.15, *P* = 1.9×10^−4^), with comparable effect sizes shown at the two time points. These associations however were not able to be replicated at the five-year and 15-year clinic.

For antibodies against herpes simplex virus, rs10961934 showed the strongest associations at different timepoints. The effect size at the seven-year clinic (β = 0.53) were similarly demonstrated at the 11-year clinic (β = 0.48, *P* = 5.6×10^−5^) and the 15-year clinic (β = 0.53, *P* = 1.4×10^−4^), although this association was not observed at the five-year clinic (*P* < 0.07).

Lastly, replication of rs1634083 associated with antibodies against measles virus was shown at the 11-year clinic (β = 0.29, *P* = 8.9 × 10^−5^), and the effect size was similar to the seven-year clinic (β = 0.33). Neither of the SNPs that associated strongly with measles showed good evidence for association at other timepoints, with the effect appearing to attenuate with age beyond seven-years.

Furthermore, we attempted to replicate the associations identified in the ALSPAC seven-year clinic in an independent cohort, UK Biobank (Supplementary Methods). In UK Biobank, data was only available for five of the antibodies, allowing replication of 14 of the 29 associations (Table 6). One association, between rs186721582 and antibodies against EBV showed strong evidence of replication (EA = G, *P* = 7.3×10^−10^, β = −0.09, 95% CI = 0.06, 0.12), an intronic variant located in genes *SUGCT* and *LOC105375245*. However, the effect size was in the opposite direction.

**Table 6.** Replication of associations observed in the ALSPAC seven-year clinic GWAS in UK Biobank (*P* < 5.9 × 10^−4^). Abbreviations: Chr, Chromosome; EA, Effect allele; Beta, Beta estimate for the presence allele; L95, lower 95% confidence interval; U95, Upper 95% confidence interval; P, *p*-value.

| Antibodies | Chr | SNP | EA | Discovery: ALSPAC |  |  |  | Replication: UK Biobank |  |  |  |
| --- | --- | --- | --- | --- | --- | --- | --- | --- | --- | --- | --- |
|  |  |  |  | Beta | L95 | U95 | P | Beta | L95 | U95 | P |
| CMV | 4 | rs10028968 | A | -0.112 | -0.157 | -0.067 | $9.5 \times 10^{-7}$ | -0.005 | -0.042 | 0.032 | 0.780 |
| EBV | 6 | rs3096702 | G | -0.116 | -0.161 | -0.071 | $4.3 \times 10^{-7}$ | 0.002 | -0.028 | 0.032 | 0.890 |
| | 7 | rs186721582 | A | 0.429 | 0.264 | 0.594 | $3.6 \times 10^{-7}$ | -0.093 | -0.123 | -0.064 | $7.3 \times 10^{-10}$ |
| | 10 | rs74807157 | A | 0.269 | 0.164 | 0.375 | $6.2 \times 10^{-7}$ | -0.052 | -0.148 | 0.045 | 0.290 |
| | 15 | rs6416551 | G | -0.21 | -0.286 | -0.134 | $7.3 \times 10^{-8}$ | 0.024 | -0.045 | 0.093 | 0.500 |
| *H.pylori* | 6 | rs35030589 | A | -0.175 | -0.242 | -0.108 | $3.4 \times 10^{-7}$ | 0.015 | -0.036 | 0.066 | 0.560 |
| HSV1 | 2 | rs6758808 | T | -0.403 | -0.556 | -0.249 | $3.8 \times 10^{-7}$ | 0.011 | -0.03 | 0.051 | 0.600 |
| | 3 | rs140033700 | A | 1.335 | 0.826 | 1.843 | $3.9 \times 10^{-7}$ | 0.004 | -0.035 | 0.043 | 0.840 |
| | 9 | rs10961934 | G | 0.533 | 0.326 | 0.741 | $6.4 \times 10^{-7}$ | -0.046 | -0.135 | 0.043 | 0.310 |
| SAG1 | 10 | rs117614721 | G | -0.748 | -1.038 | -0.457 | $5.4 \times 10^{-7}$ | 0.026 | -0.025 | 0.077 | 0.320 |
| *T.gondii* | 1 | rs10788882 | C | -0.116 | -0.162 | -0.07 | $8.8 \times 10^{-7}$ | 0.055 | -0.029 | 0.14 | 0.200 |
| | 1 | rs72717048 | G | -0.219 | -0.302 | -0.135 | $2.7 \times 10^{-7}$ | -0.013 | -0.043 | 0.018 | 0.420 |
| | 3 | rs190637453 | C | 0.53 | 0.324 | 0.736 | $4.8 \times 10^{-7}$ | 0.023 | -0.033 | 0.079 | 0.420 |
| | 11 | rs36020612 | T | 0.202 | 0.147 | 0.258 | $1.2 \times 10^{-12}$ | 0.015 | -0.116 | 0.146 | 0.830 |

We also conducted the analysis in the opposite direction, with a GWAS in UK Biobank as a discovery cohort and ALSPAC seven-year clinic as replication. Five antibodies in UK Biobank overlapped with ALSPAC (Cytomegalovirus, Epstein-Barr virus, *Helicobacter pylori*, herpes simplex virus 1, and *Toxoplasma gondii*), and findings are shown in Table 7. In total, 12 associations met the genome-wide threshold (*P* < 5×10^−8^) with Epstein-Barr virus (10 associations), herpes simplex virus 1 (one association) and *T*.*gondii* (one association) in UK Biobank (Table 7). In ALSPAC, five of the associations with antibodies against Epstein-Barr virus replicated (*P* < 0.001): rs9264759, rs1043620, rs2523502, rs3117139 and rs3096695, with effects in the same direction compared to UK Biobank, with the exception of rs1043620 in which the effect was in the opposite direction (Table 7). The remaining two EBV antibody associations did not replicate (*P* > 0.485). In addition, three EBV antibody associations were not able to be replicated as no suitable LD proxy SNP was identified (R^2^ < 0.6) in ALSPAC, and similarly none of the associations with antibodies against herpes simplex virus 1 or *T*.*gondii* were able to be replicated.

**Table 7.**
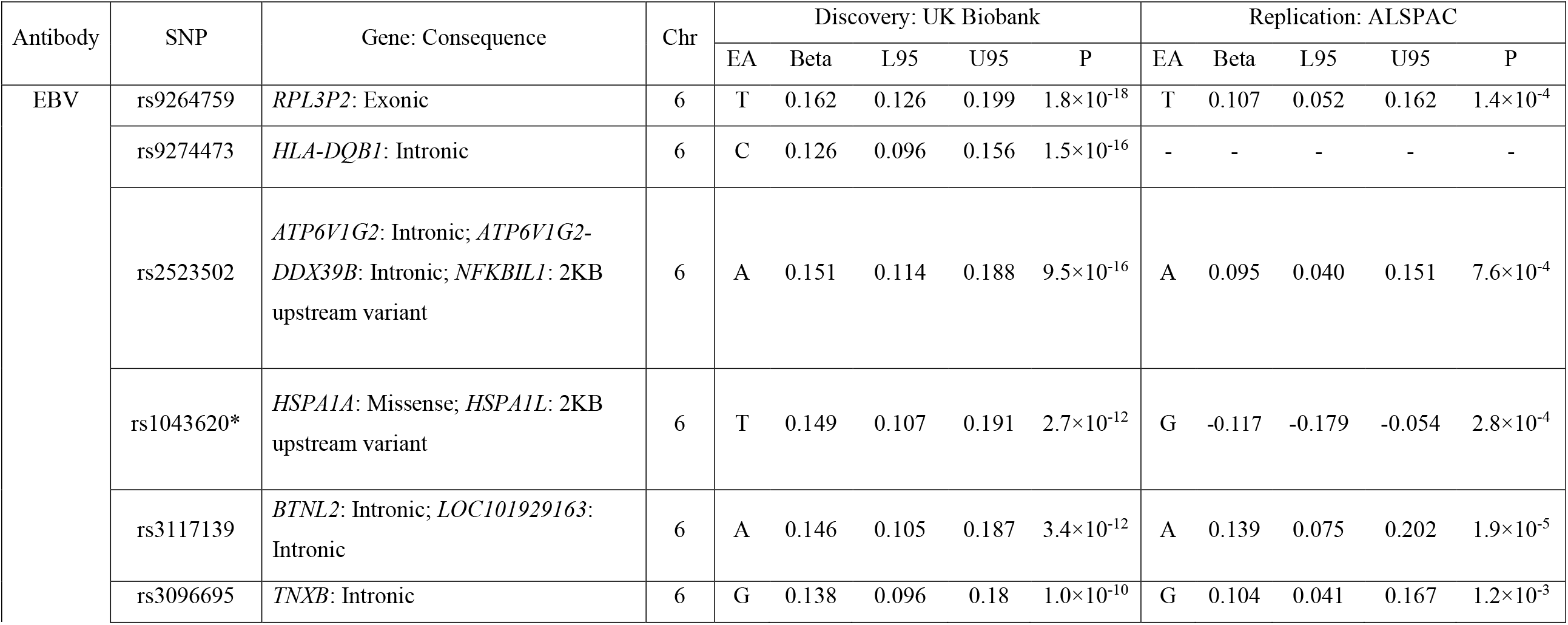
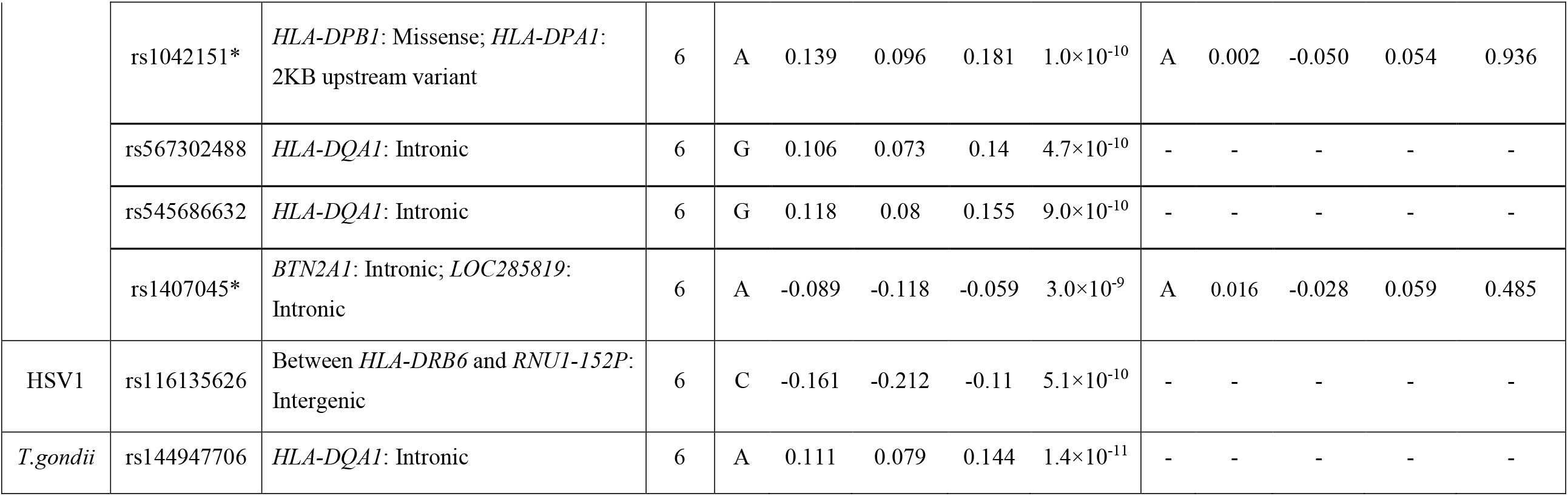
Top SNPs in discovery GWAS in UK Biobank with replication GWAS in ALSPAC seven-year clinic (*P* < 0.001). (-) refers to SNPs that were not present in the reference panel, and no suitable SNP proxy was identified (R^2^ < 0.6); (*) SNP proxies have been used in ALSPAC replication with rs9267551 a proxy for rs1043620 (R^2^ = 0.98), rs3130213 a proxy for rs1042151 (R^2^ = 0.82), and rs2145318 a proxy for rs1407045 (R^2^ = 1). Abbreviations: Chr, Chromosome; EA, Effect allele; Beta, Beta estimate for the presence allele; L95, lower 95% confidence interval; U95, Upper 95% confidence interval; P, *p*-value.

| Antibody | SNP | Gene: Consequence | Chr | Discovery: UK Biobank |  |  |  |  | Replication: ALSPAC |  |  |  |  |
| --- | --- | --- | --- | --- | --- | --- | --- | --- | --- | --- | --- | --- | --- |
|  |  |  |  | EA | Beta | L95 | U95 | P | EA | Beta | L95 | U95 | P |
| EBV | rs9264759 | *RPL3P2*: Exonic | 6 | T | 0.162 | 0.126 | 0.199 | $1.8 \times 10^{-18}$ | T | 0.107 | 0.052 | 0.162 | $1.4 \times 10^{-4}$ |
| | rs9274473 | *HLA-DQB1*: Intronic | 6 | C | 0.126 | 0.096 | 0.156 | $1.5 \times 10^{-16}$ | - | - | - | - | - |
| | rs2523502 | *ATP6V1G2*: Intronic; *ATP6V1G2-DDX39B*: Intronic; *NFKBIL1*: 2KB upstream variant | 6 | A | 0.151 | 0.114 | 0.188 | $9.5 \times 10^{-16}$ | A | 0.095 | 0.040 | 0.151 | $7.6 \times 10^{-4}$ |
| | rs1043620* | *HSPA1A*: Missense; *HSPA1L*: 2KB upstream variant | 6 | T | 0.149 | 0.107 | 0.191 | $2.7 \times 10^{-12}$ | G | -0.117 | -0.179 | -0.054 | $2.8 \times 10^{-4}$ |
| | rs3117139 | *BTNL2*: Intronic; *LOC101929163*: Intronic | 6 | A | 0.146 | 0.105 | 0.187 | $3.4 \times 10^{-12}$ | A | 0.139 | 0.075 | 0.202 | $1.9 \times 10^{-5}$ |
| | rs3096695 | *TNXB*: Intronic | 6 | G | 0.138 | 0.096 | 0.18 | $1.0 \times 10^{-10}$ | G | 0.104 | 0.041 | 0.167 | $1.2 \times 10^{-3}$ |
| | rs1042151* | *HLA-DPBI*: Missense; *HLA-DPA1*:<br>2KB upstream variant | 6 | A | 0.139 | 0.096 | 0.181 | $1.0 \times 10^{-10}$ | A | 0.002 | -0.050 | 0.054 | 0.936 |
| | rs567302488 | *HLA-DQAI*: Intronic | 6 | G | 0.106 | 0.073 | 0.14 | $4.7 \times 10^{-10}$ | - | - | - | - | - |
| | rs545686632 | *HLA-DQAI*: Intronic | 6 | G | 0.118 | 0.08 | 0.155 | $9.0 \times 10^{-10}$ | - | - | - | - | - |
| | rs1407045* | *BTN2A1*: Intronic; *LOC285819*:<br>Intronic | 6 | A | -0.089 | -0.118 | -0.059 | $3.0 \times 10^{-9}$ | A | 0.016 | -0.028 | 0.059 | 0.485 |
| HSV1 | rs116135626 | Between *HLA-DRB6* and *RNU1-152P*:<br>Intergenic | 6 | C | -0.161 | -0.212 | -0.11 | $5.1 \times 10^{-10}$ | - | - | - | - | - |
| *T.gondii* | rs144947706 | *HLA-DQAI*: Intronic | 6 | A | 0.111 | 0.079 | 0.144 | $1.4 \times 10^{-11}$ | - | - | - | - | - |

### Evaluation of genetic overlap of ALSPAC antibodies with published SARS-CoV-2 GWAS meta-analyses

We performed a lookup of genetic signals identified to be genome-wide and suggestively associated with the ALSPAC seven-year clinic antibodies (Table 2 and 3) in a recent COVID-19 Host Genetics Initiative GWAS meta-analyses of SARS-CoV-2 antibodies(8) in European populations (Supplementary Methods). There was no evidence of the 29 SNPs being associated with SARS-CoV-2 (All *P* > 0.1) (Supplementary Table 4) and only 11 SNPs exhibited the same direction of effect.

We also evaluated the seven SNPs identified as strongly associated with SARS-CoV-2 antibodies(8) in the ALSPAC seven-year clinic antibody GWASs. However, there was little evidence for associations between these seven SNPs and any of the 14 antibodies tested in ALSPAC (only three of 98 tested had *P* < 0.05, in line with chance expectations (Supplementary Table 5).

### Association between HLA alleles and antigen specific antibodies

Imputation of the HLA region was conducted in ALSPAC using two methods (HLA:IMP*03 and SNP2HLA). We found very high concordance between posterior probabilities of imputed genotypes between the two methods at two- and four-digit resolutions (Supplementary Table 6 and 7). In addition, very high concordance was shown between allele frequencies for HLA loci measured using the two approaches and compared to the Type 1 Diabetes Genetics Consortium (T1DGC) HLA reference panel (Supplementary Table 8 and 9).

We performed HLA association analyses for 14 antibodies at two- and four-digit resolutions using both HLA imputation methods in ALSPAC. The associations that exceeded the HLA-wide corrected -value thresholds at a 2-digit resolution (*P* < 6.2 × 10^−4^) and at a 4-digit resolution (*P* < 5.0 × 10^−4^) are shown in Table 8 (Full results and Q-Q plots illustrated in Supplementary Table 10 and 11 and Supplementary Figure 3 and 4). Associations for five antibodies with 15 HLA alleles were identified at a two-digit resolution (Table 8), and four antibodies with 23 HLA alleles at a four-digit resolution (Table 8). Findings using the two HLA imputation methods, HLA:IMP*03 and SNP2HLA, produced similar results (Supplementary Table 10 and 11).

**Table 8.**
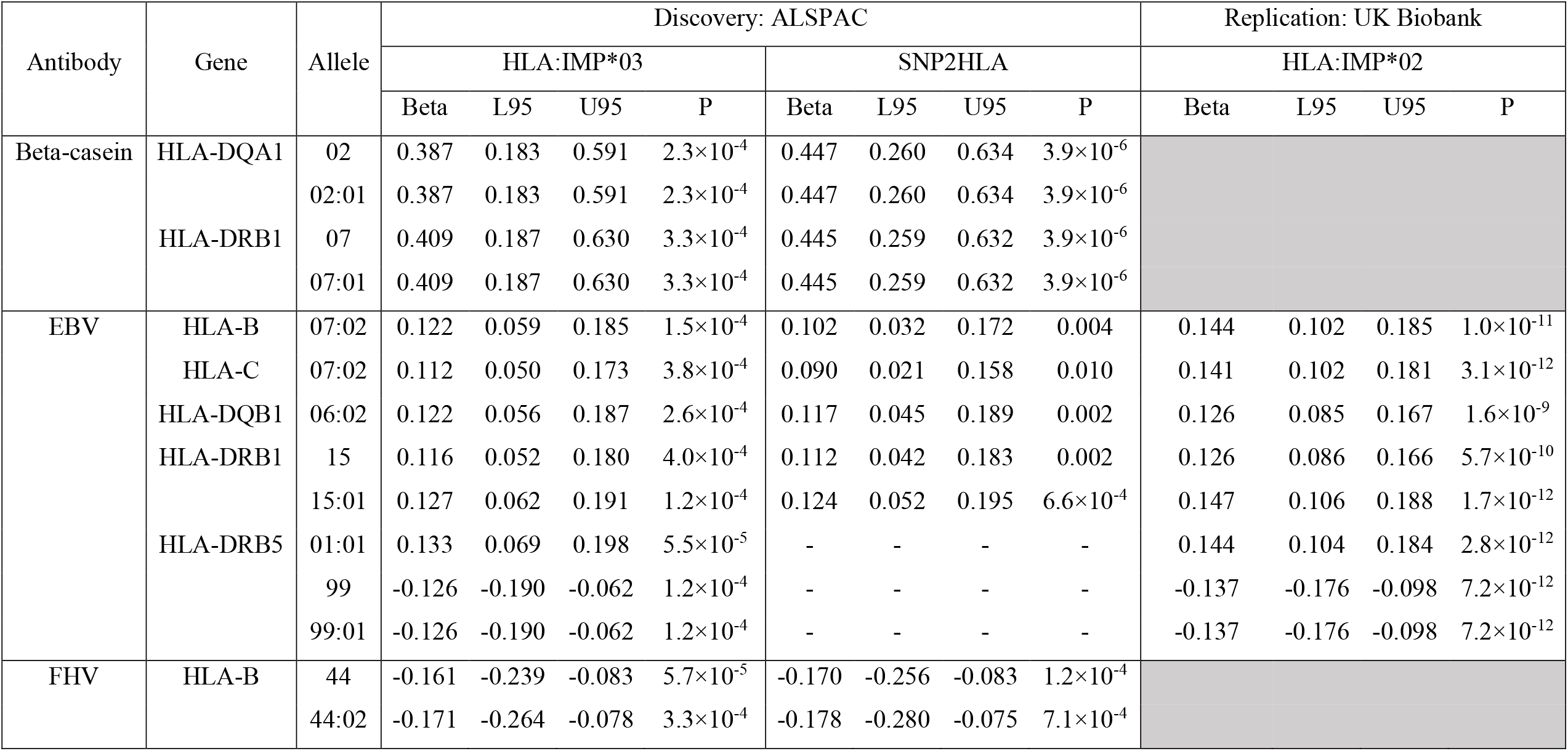
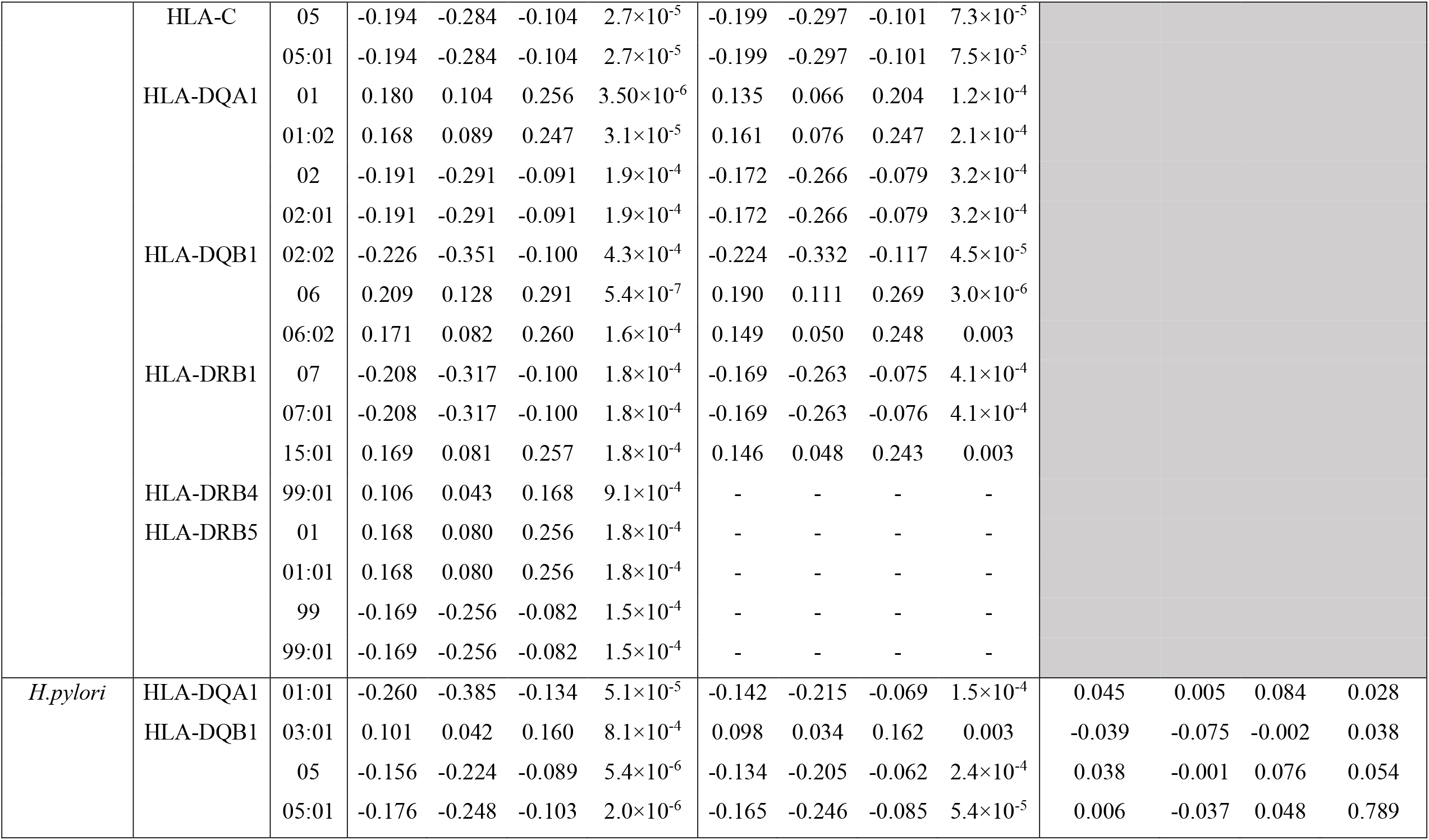
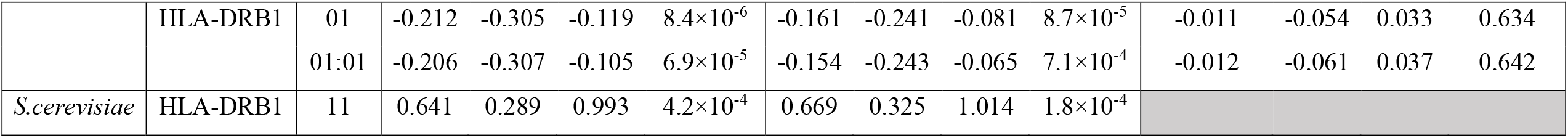
HLA alleles at a 2-digit resolution (*P* < 6.2 × 10^−4^) and 4-digit resolution (*P* < 5.0 × 10^−4^) associated with antibodies measured at the ALSPAC seven-year clinic and replication in UK Biobank (*P* < 0.004). (-) refers to HLA alleles not present in reference panel. Shaded cells refer to antibodies not measured in UK Biobank. Abbreviations: Beta, Beta estimate for the presence allele; L95, lower 95% confidence interval; U95, Upper 95% confidence interval; P, *p*-value.

When further correcting for the 14 antibodies tested, seven HLA alleles at a 2-digit resolution (*P* < 4.4×10^−5^) remained associated with antibodies against beta-casein protein, feline herpes virus and *H*.*pylori*. At a 4-digit resolution (*P* < 3.5×10^−5^), six HLA alleles remained associated with antibodies against beta-casein protein, Epstein-Barr virus, feline herpes virus, and *H*.*pylori*.

We then attempted replication of the ALSPAC HLA associations in UK Biobank (Supplementary Methods) for EBV and *H*.*pylori* antibodies (other infections were not available). All eight Epstein Barr virus antibodies associations showed strong replication signals with the same direction of effect, with *P* <1×10^−9^. However, the four associations with *H*.*pylori* did not replicate (Table 8).

We also performed HLA-wide association testing in UK Biobank with replication in ALSPAC for five overlapping antibodies: Cytomegalovirus, Epstein-Barr virus, *H*.*pylori*, herpes simplex virus 1, and SAG1 protein domain. Full UK Biobank results are shown in Supplementary Table 12 and 13 and Q-Q plots are illustrated in Supplementary Figure 5. In summary, 22 HLA associations were identified with four antibodies at a 2-digit resolution (*P* < 7.1×10^−5^), and 17 HLA associations were identified with five antibodies at a four-digit resolution (*P* < 2.8×10^−5^) in UK Biobank (Supplementary Table 14). Four HLA alleles at a 2-digit resolution (*P* < 0.003) and five HLA alleles at a four-digit resolution (*P* < 0.003) associated with Epstein-Barr virus antibodies replicated in ALSPAC, with all effect estimates consistently showing the same direction of effects (Supplementary Table 14).

## Discussion

In this study, we investigated host genetic contributions to measured antibodies in the ALSPAC cohort. In determining how to analyse antibodies, we found little evidence to support thresholding in order to categorise individuals into seropositive and seronegative. Our analysis suggested that analysing the antibody data as continuous variables is more appropriate.

In genome-wide association analysis, we observed three genetic signals strongly associated (*P* < 5 × 10^−8^) with two antibodies, measles virus and *T*.*gondii*, at the ALSPAC seven-year clinical time point. Associations linking these loci with their respective antibodies have not been previously reported. Assessing the 26 associations that met a suggestive threshold (*P* < 1 × 10^−6^), the association between rs186721582 and Epstein-Barr virus antibodies replicated in UK Biobank. The associations between rs36020612 and *T*.*gondii* did not replicate (*P* > 5.9 × 10^−4^) and antibody data on the measles virus was not available. Stratifying individuals by MMR vaccine status for SNPs associated with antibodies against measles virus at the seven-year clinic showed stronger signal for individuals with an unknown status (Table 4), with comparable distributions between status. This suggests that antibody response to measles virus is more likely through environmental exposure or could be attributable to issues when using ELISAs such as high background and weak signal intensity leading to stochastic antibody levels, or potentially due to the larger sample size in the ‘unknown’ group.

Replication of the SNPs at several clinical time points in ALSPAC (i.e. five, seven-, 11-, and 15-years) showed no associations observed at age five, but several associations persisted beyond age seven (FHV, *H*.*pylori*, HSV1, measles virus, *T*.*gondii*). For FHV antibodies, *H*.*pylori* and measles virus, the observed associations disappeared at the 15-year time point, which could be attributable to the reduction in sample size, lack of phenotypic variance, and possible false-positive associations identified at the seven-year time point.

Of particular interest in the main ALSPAC GWAS, was the association between rs36020612 located on *GRAMD1B* with antibodies against *T*.*gondii* (Table 2). Expression of this gene has been related to lymphocyte, erythrocyte and antibody traits which could suggest a link to immune response. Specifically, chronic lymphocytic leukaemia(32, 34-36), lymphocyte percentage and count(37), mean corpuscular haemoglobin concentration(38, 39), erythrocyte distribution width(38, 40), blood protein levels(41), and immunoglobulin M (IgM) antibody levels(31, 42). However, while this association persisted at later time points in ALSPAC, it did not replicate in the UK Biobank. In addition, rs36020612 was not shown to be associated with antibodies for SAG1 protein domain (*P* = 0.78), an antigen of *T*.*gondii,* with the effect size in the opposite direction. This lack of association could be due to the difference in statistical power contributing to the sample size of SAG1 protein domain (N = 1228) compared to *T*.*gondii* (N = 5010).

We also specifically investigated associations between the major histocompatibility complex (*MHC*) class II region in ALSPAC and identified 20 associations of interest with antibodies against beta-casein protein, Epstein-Barr virus, feline herpes virus, *H*.*pylori*, and *S*.*cerevisiae* (Table 8). Associations with antibodies against Epstein-Barr virus again replicated in UK Biobank.

The strongest association we observed in the HLA region in ALSPAC and UK Biobank was between HLA-DRB1*15:01 and Epstein-Barr virus antibodies, which has been previously reported (43) and is a well-established genetic risk factor associated with increased risk for MS(44-46). HLA-DRB1:15*01 positive humanized mice with EBV infection were similarly shown to demonstrate dysregulation in immune response through attenuated CD4^+^ and CD8^+^T cell activation and decreased efficiency to control EBV viral loads(47). Of the other five 4-digit HLA alleles that showed strong association with EBV, HLA-DRB5*01:01 and HLA:DQB1*06:02 are in strong linkage disequilibrium with the previously discussed HLA:DRB1*15:01(48). HLA-B*07:02 has also been investigated for its association with EBV and the latent-specific CD8^+^T cell response as well as it link to MS(49, 50). However, no current literature has researched HLA-C*07:02 and HLA-DRB5*99:01 in respect to their relationship with EBV.

Four HLA alleles at a four-digit resolution were associated with *H*.*pylori* antibodies, with HLA-DQB1*05:01 showing the strongest association followed by HLA-DQA1*01:01, HLA-DRB1*01:01, and lastly HLA-DQB1*03:01. All four HLA alleles were unable to be replicated in UK Biobank (*P* < 5.0 × 10^−4^) which could possibly be a result of false-positive associations in ALSPAC at the seven-year clinic, and lack of phenotypic variance. To date, no studies have investigated the association of these HLA alleles and *H*.*pylori* in individuals of European ancestry. Studies using other ancestral populations have shown that HLA-DQB1*05:01 strongly increased the risk of gastric cancer in individuals of Mexican descent(51), and similarly in a Chinese population HLA-DRB1*01 was demonstrated to strongly increase in the risk of gastric cancer in *H*.*pylori*-positive individuals(52). This type of cancer is commonly associated with *H*.*pylori* infection, which is the is the strongest known risk factor for the development of this disease(53, 54). At present, no literature has not shown a relationship between both HLA-DQA1*01:01 and HLA-DQB1*03:01 with *H*.*pylori*.

There are several limitations to this study. Firstly, the potential limited exposure to the infections of interest in the ALSPAC sample of children as a result of their young age may have reduced power to detect associations, as viruses such as cytomegalovirus, Epstein-Barr virus and herpes simplex virus 1 show an increase in prevalence into adulthood (27, 55-57). We attempted to address this issue by assessing genetic signals at later clinical time points, and replicate the findings in the adult cohort, UK Biobank. Secondly, the poor evidence in some instances of genetic signal replication at different timepoints in the ALSPAC data may be owing to the small sample sizes of some of the measured antibodies. Eight antigens had a sample size of less than 700 individuals at the seven-year clinic which is the time point with the largest sample size for all measured antibodies. An approach to maximise sample size could have been to keep all individuals with antibody levels measured against infections at all clinical time points and remove any duplicate individuals retaining only individuals with antibody levels measured at the latest time point. This approach could increase potential cases as older individuals may be more likely, by virtue of time, to become infected through environmental exposure, however this approach could potentially introduce heterogeneity.

Thirdly, not all infections of interest had measured antibodies in UK Biobank to test for replication, and furthermore there were differences in assays used to measure antibody response between cohorts. Lastly, the interpretation of the genetic signals is difficult to disentangle using association studies as these signals could be related to susceptibility to infection and persistence of the antibodies. For example, studies have shown a reduction in antibody titers to influenza virus subtype H1N1 a year to 18 months after initial vaccination(58-61). In contrast, the two-dose vaccination programme of the measles virus has been demonstrated to produce a sustained protective antibody persistence(62-64). In addition, if a genetic effect is shown to be associated with higher antibody levels, is the interpretation that it influences the chance of being infected, or that it influences the antibody response to infection, or potentially that it influences a long-lasting antibody response. To disentangle this challenge better approaches to measuring antibody responses against infection are required, such as the development of reliable uninfected baseline population distributions. Furthermore, analyses from *in vivo* and candidate gene studies to provide stronger evidence in support of these findings, and to examine the possible functional roles of the genetic variants and genes to elucidate the underlying biological mechanisms, is required.

In summary, our study has confirmed known HLA allele associations with Epstein-Barr virus in a younger age group (seven-years) and replicated results in the adult population, UK Biobank. In addition, in the discovery phase we have identified four potentially novel genetic associations, with rs36020612 showing strong evidence of association with *T*.*gondii* in children at the seven-year clinic, and at later clinical time-points. The location of this SNP on *GRAMD1B* is of particular interest as expression of this gene has been shown to be related to lymphocyte traits. We also found strong evidence of two SNPs (rs506576 and rs28617484) associated with antibodies against measles virus, however we were unable to test for replication in UK Biobank due to data unavailability. Furthermore, we observed strong evidence of replication of the suggestive SNP (rs186721582) associated with Epstein-Barr virus antibodies in UK Biobank, suggesting the genetic effect is stronger in adults. Our study provides a useful resource for future studies looking to use ALSPAC antibody measurements as well as HLA alleles. It indicates that for future use of the measured antibodies in ALSPAC the most appropriate approach is to use the antibody titer measures as a continuous variable. This study has highlighted the potential for identification of host genetic risk factors for several common infections, and demonstrates that if similar data is collected in other cohorts, future meta-analysis GWAS is likely to be a fruitful endeavour in uncovering the genetic and biological mechanisms of infection susceptibility.

## Data Availability

The study website contains details of all the data that is available through a fully searchable data dictionary and variable search tool.

http://www.bristol.ac.uk/alspac/researchers/our-data/

## Author contributions

The study concept and design was conceived by GH, LP, and RCR. AHWC, LP, and RCR contributed in the acquisition, analysis and/or interpretation of data. AHWC wrote the manuscript and all co-authors contributed to the critical revision of the manuscript before approving its submission. This publication is the work of the authors: AHWC, REM, GH, GDS, RCR, and LP; and will serve as guarantors for the contents of this paper.

## Acknowledgements

We are extremely grateful to all the families who took part in this study, the midwives for their help in recruiting them, and the whole ALSPAC team, which includes interviewers, computer and laboratory technicians, clerical workers, research scientists, volunteers, managers, receptionists and nurses. We thank Ildar Sadreev for his helpful advice in the analysis of this study.

