## Supplementary Methods and Figures for "Genetic analyses of common infections in the Avon Longitudinal Study of Parents and Children cohort"

### *Supplementary Material*

#### Supplementary Methods

<u>UK Biobank</u>

*Cohort profile*

UK Biobank is a population-based heath research resource(1). This cohort consists of approximately 500,000 participants, aged between 38-73 years, who were recruited between the years of 2006 to 2010 across the United Kingdom(1). Comprehensively measured data has been collected on variables related to cognitive function, demographic information, socio-economic measures, physical and mental health measures, health status, and lifestyle measures, with an objective to identify determinants of human diseases(2). Data was collected through various approaches such as anthropometric measures, biological samples (i.e. Blood, urine and saliva), and via questionnaires and interviews(2). A full description of the study design, cohort characteristics and quality control approaches have been previously described(3).

*Phenotype measurements*

Description of the assays and validation protocol for each individual antigen have been described elsewhere(4). In brief, a subset of randomly selected participants (n = 9,695) were chosen and serum samples were taken and assayed using a Multiplex Serology panel. Five antigens were selected: p150 Nter antigen for human cytomegalovirus; viral capsid antigen (VCA) p18 for Epstein-Barr virus; 1gG antigen for Herpes simplex virus 1; Outer membrane protein (OMP) surface antigen for *Helicobacter pylori*; and surface antigen 1 (SAG1) protein domain for *Toxoplasma gondii*. For analyses, the initial assessment time point was selected as it represented the largest sample size for all antigens (N = 9,430), and the continuous measures were standardised with a mean of 2 and a standard deviation of 1. In addition, the standardised measures were rank-based inverse normal transformed to a normal distribution. More details of the antigens can be found on UK Biobank Data Showcase: https://biobank.ctsu.ox.ac.uk/showcase/refer.cgi?id=1348.

*Genotyping and imputation*

In total, the full data release contains 488,377 participants who were successfully genotyped. In particular, 49,979 participants were genotyped using the UK BiLEVE array, and the remaining 438,398 participants were genotyped using the UK Biobank axiom array. A full description of the pre-imputation quality control methods, phasing and imputation have been previously described elsewhere(5). In brief, multiallelic SNPs or SNPs with minor allele frequencies ≤ 1% were removed prior to phasing. The phasing of the genotype data was performed using an amended version of the SHAPEIT2 algorithm(6). Imputation of the genotype data to a reference set consisted of combining the UK10K haplotype and Haplotype Reference Consortium (HRC) reference panels(7) performed using IMPUTE2(8). The analyses described were restricted to autosomal variants within the HRC site list using a graded filtering with varying imputation quality for different allele frequency ranges. For rarer SNPs, these SNPs were included if they had an INFO score > 0.3 for minor allele frequency > 3%; INFO score > 0.6 for minor allele frequency 1-3%; INFO score > 0.8 for minor allele frequency 0.5-1%, INFO score > 0.9 for minor allele frequency 0.1-0.5%), with the INFO scores and minor allele frequencies recalculated on an in-house derived ‘European’ subset(9).

Quality Control filtering of the UK Biobank data was conducted by R.Mitchell, G.Hemani, T.Dudding, L.Corbin, S.Harrison, L.Paternoster as described in the published protocol

(DOI:10.5523/bris.1ovaau5sxunp2cv8rcy88688v). The MRC IEU UK Biobank GWAS pipeline was developed by B.Elsworth, R.Mitchell, C.Raistrick, L.Paternoster, G.Hemani, T.Gaunt (DOI: 10.5523/bris.pnoat8cxo0u52p6ynfaekeigi). Participants were excluded from analysis if they had sex-chromosome aneuploidy or sex-mismatch (N= 814). Furthermore, the sample was restricted to individuals of 'European' ancestry, defined by performing an in-house k-means cluster analyses using the first four principal components provided by UK Biobank using R(10), which resulted in 464,708 included participants used for analyses(9).

*Association analyses*
Genome-wide association analyses was performed using BOLT-LMM (v.2.3)(11) (https://data.broadinstitute.org/alkesgroup/BOLT-LMM/), implemented using the MRC IEU UK Biobank GWAS pipeline (DOI: 10.5523/bris.pnoat8cxo0u52p6ynfaekeigi)(12). Quality control filtering was performed using PLINKv2.00(13) (www.cog-genomics.org/plink/2.0/) SNPs were excluded if they had a genotyping call rate > 0.015, a minor allele frequency > 0.01, exhibited a Hardy-Weinberg equilibrium p-value < 0.0001, and were LD pruned to an $r^2$ threshold of 0.001. Subsequently, population structure was modelled on the 143,006 directly genotyped SNPs. Analyses were adjusted for sex, age, and the first 40 principal components under an additive genetic model.

*HLA association analyses*
Imputation of HLA alleles in UK Biobank has been comprehensively described in more detail on the UK Biobank Data Showcase: https://biobank.ndph.ox.ac.uk/ukb/ukb/docs/HLA_imputation.pdf. In brief, HLA imputation was performed using HLA*IMP:02 (14), with modifications to the multi-population reference panel to accommodate the addition of samples(15). 11 classical HLA genes were imputed, including: HLA-A, HLA-B, HLA-C, HLA-DRB1, HLA-DRB3, HLA-DRB4, HLA-DRB5,HLA-DQA1, HLA-DQB1, HLA-DPA1, HLA-DPB1(14). Prior to analyses, HLA:IMP*02 output was converted into a dosage format by calculating the expected dosage of each allele. The dosage for each individual was calculated by summing the posterior probabilities of each HLA allele. HLA analyses was performed using PLINK(v1.90) (www.cog-genomics.org/plink/1.9/)(16), adjusting for age, sex, and the first 40 principal components.

COVID-19 Host Genetics Initiative
The COVID-19 Host Genetics Initiative(17) is a global collaborative platform initiated to identify SNPs strongly associated with COVID-19 susceptibility, severity, and COVID-19 related outcomes. Publicly available SARS-CoV-2 GWAS meta-analysed summary statistics are accessible through: https://www.covid19hg.org/results/. For the genetic overlap analysis, the GWAS meta-analysis 'C2_ALL_eur' released in January 2021 was selected. 36 cohorts took part in the GWAS meta-analysis investigating COVID-19 positive cases (N = 38,984) versus European population as controls (N = 1,644,784) in participants of European ancestry. Six independent SNPs associated with COVID-19 were identified, and these SNPs were used in the genetic overlap analysis.

#### Supplementary Figures

(A) Alpha-casein protein

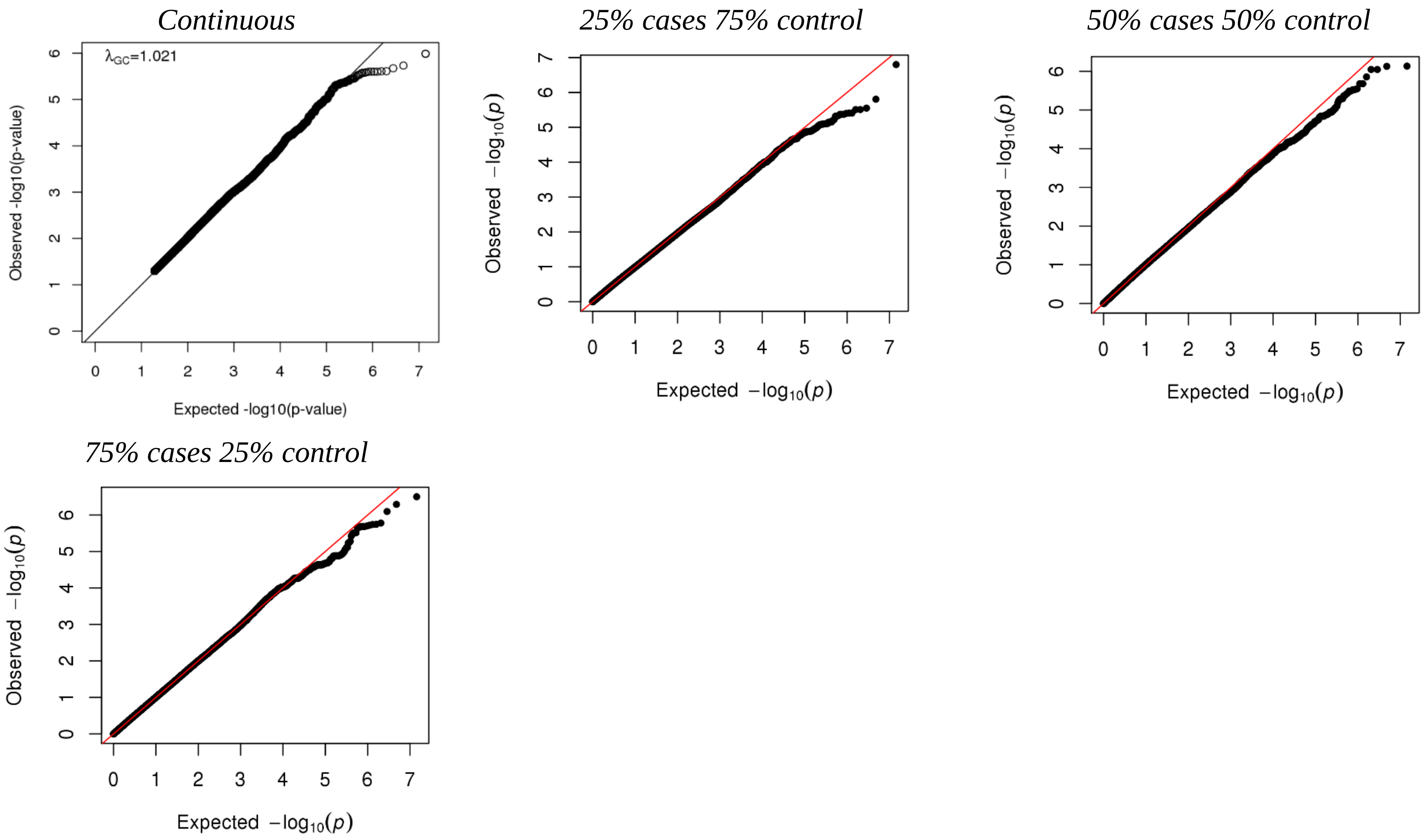

(B) Beta-casein protein

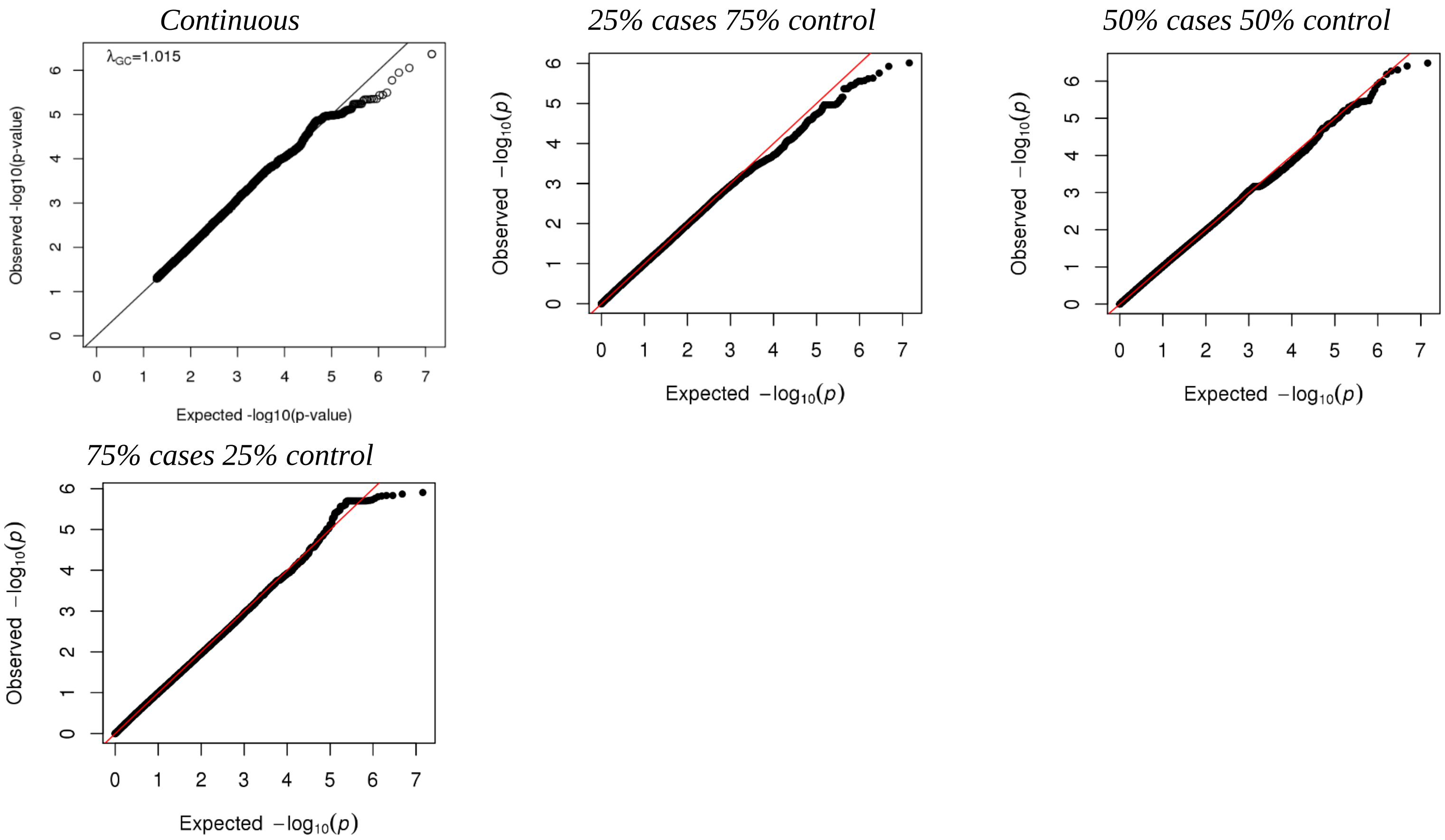

(C) Cytomegalovirus

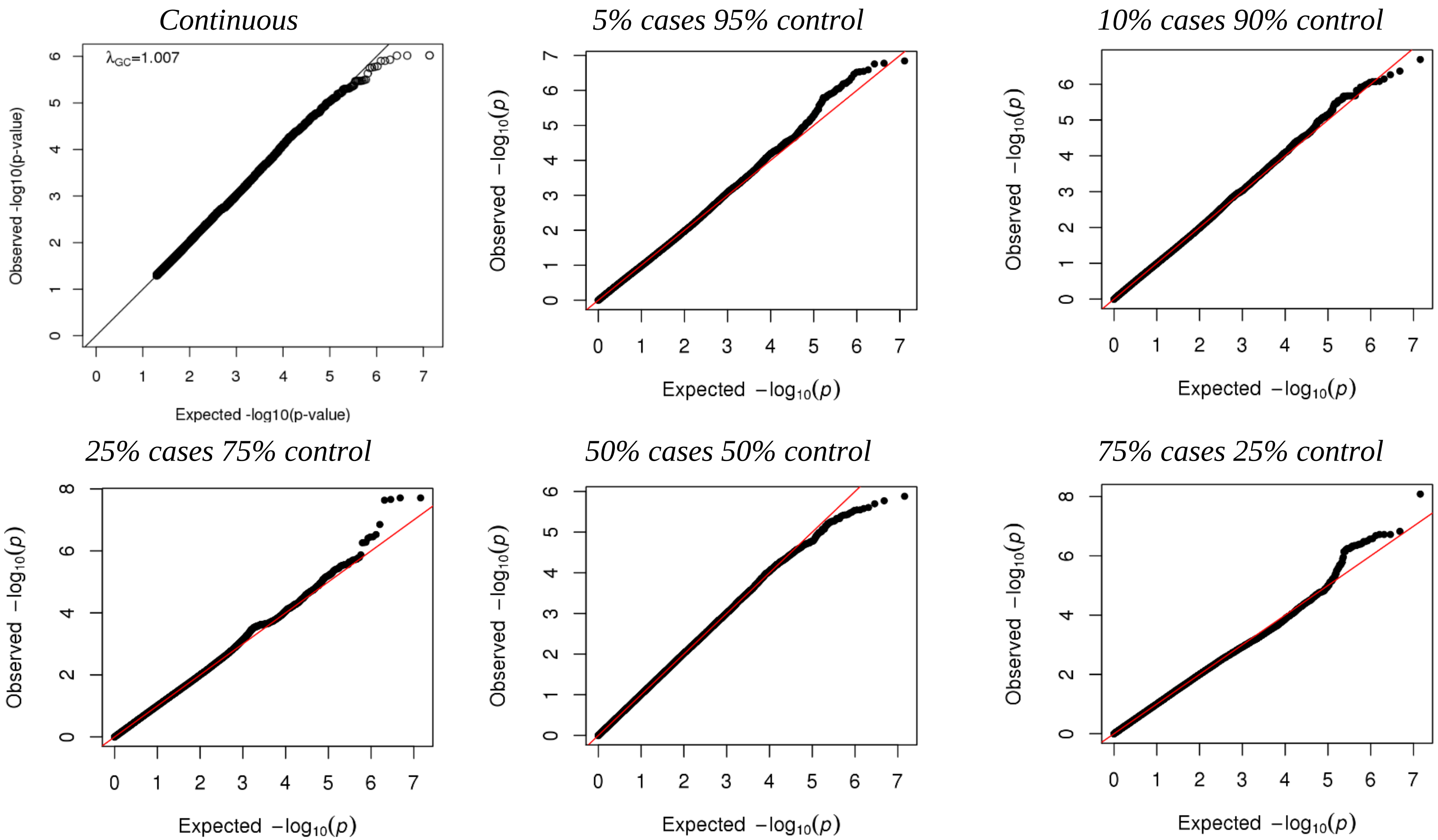

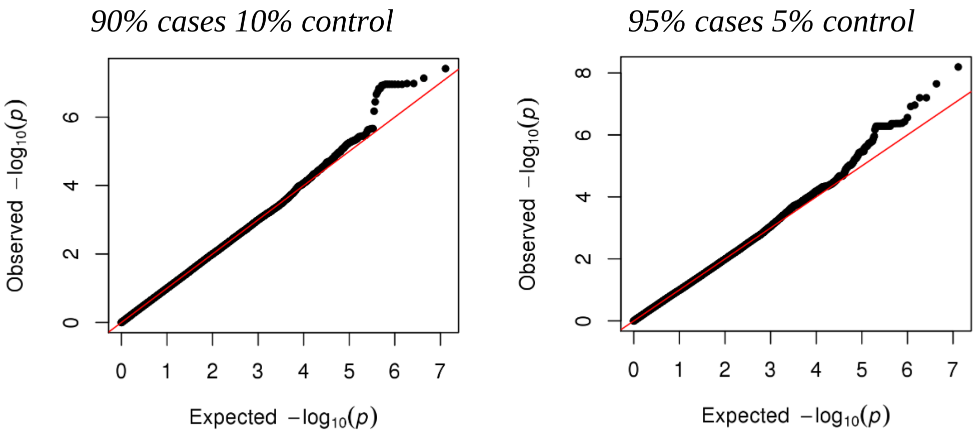


(D) Epstein-Barr virus

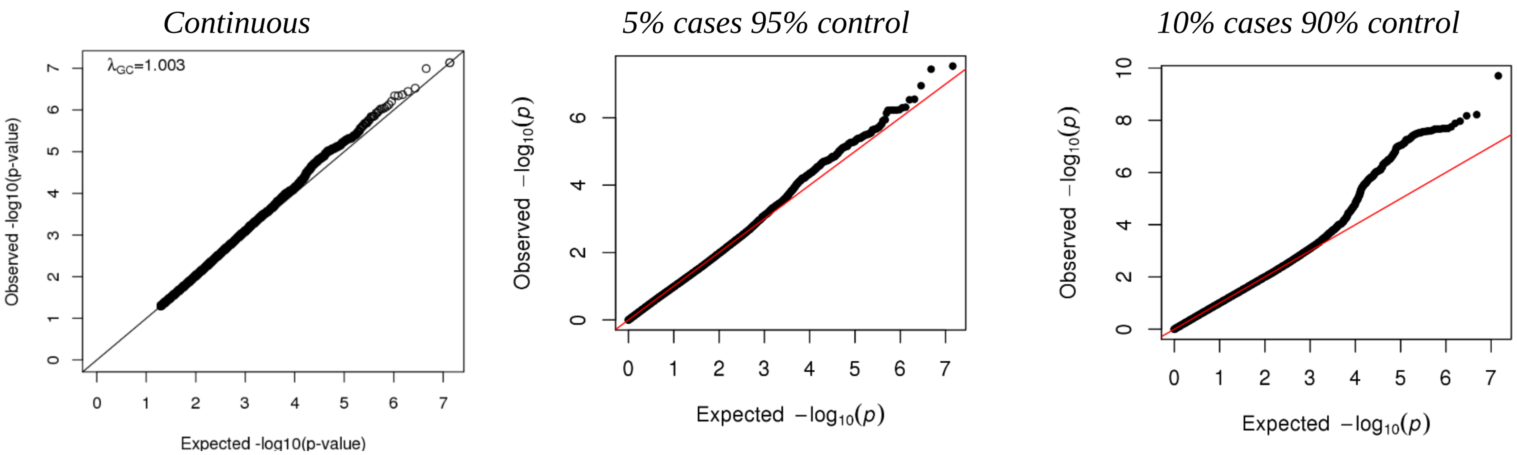

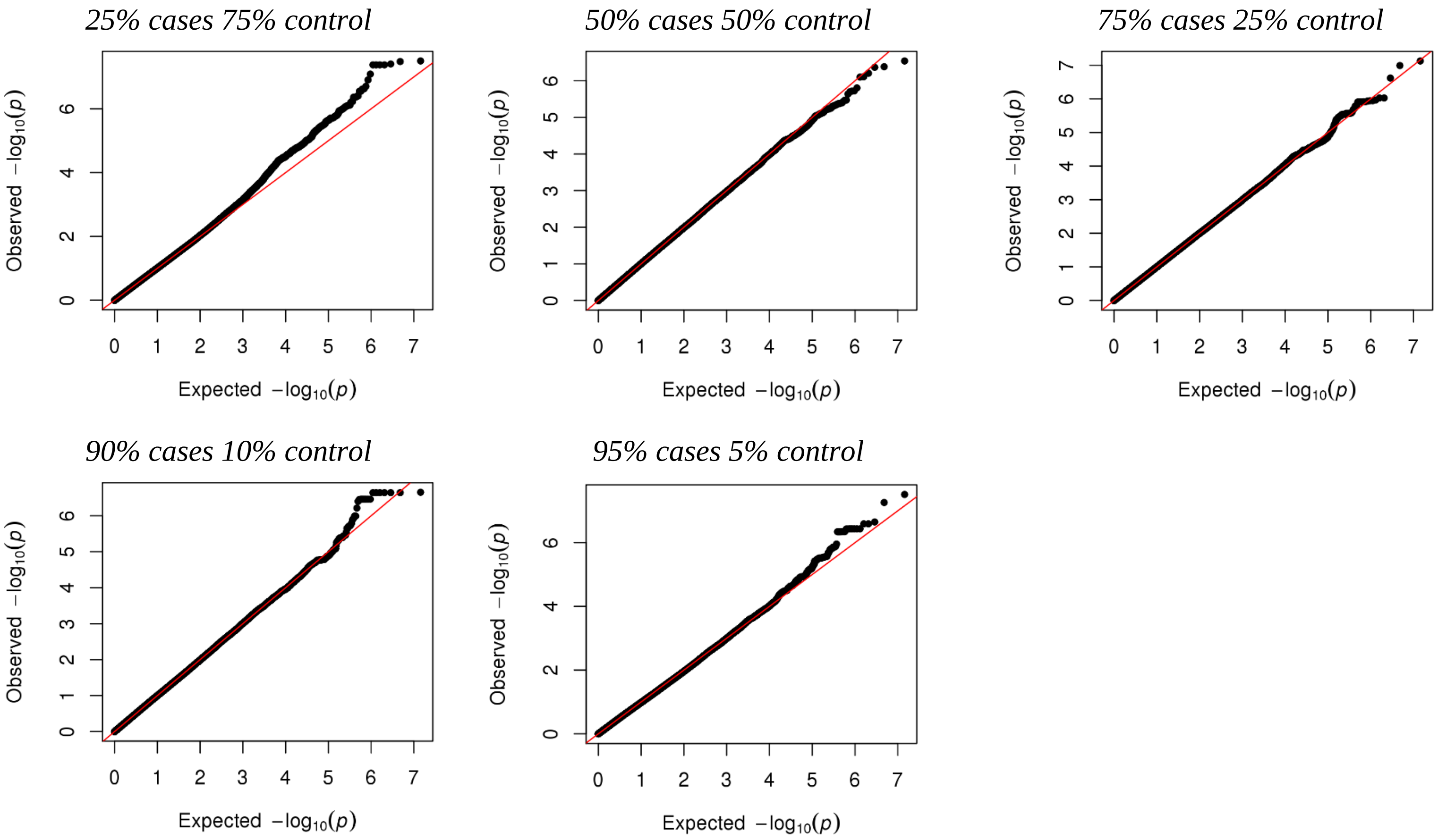
25% cases 75% control
50% cases 50% control
75% cases 25% control
90% cases 10% control
95% cases 5% control
Observed $-\log_{10}(p)$
Expected $-\log_{10}(p)$

(E) Feline herpes virus

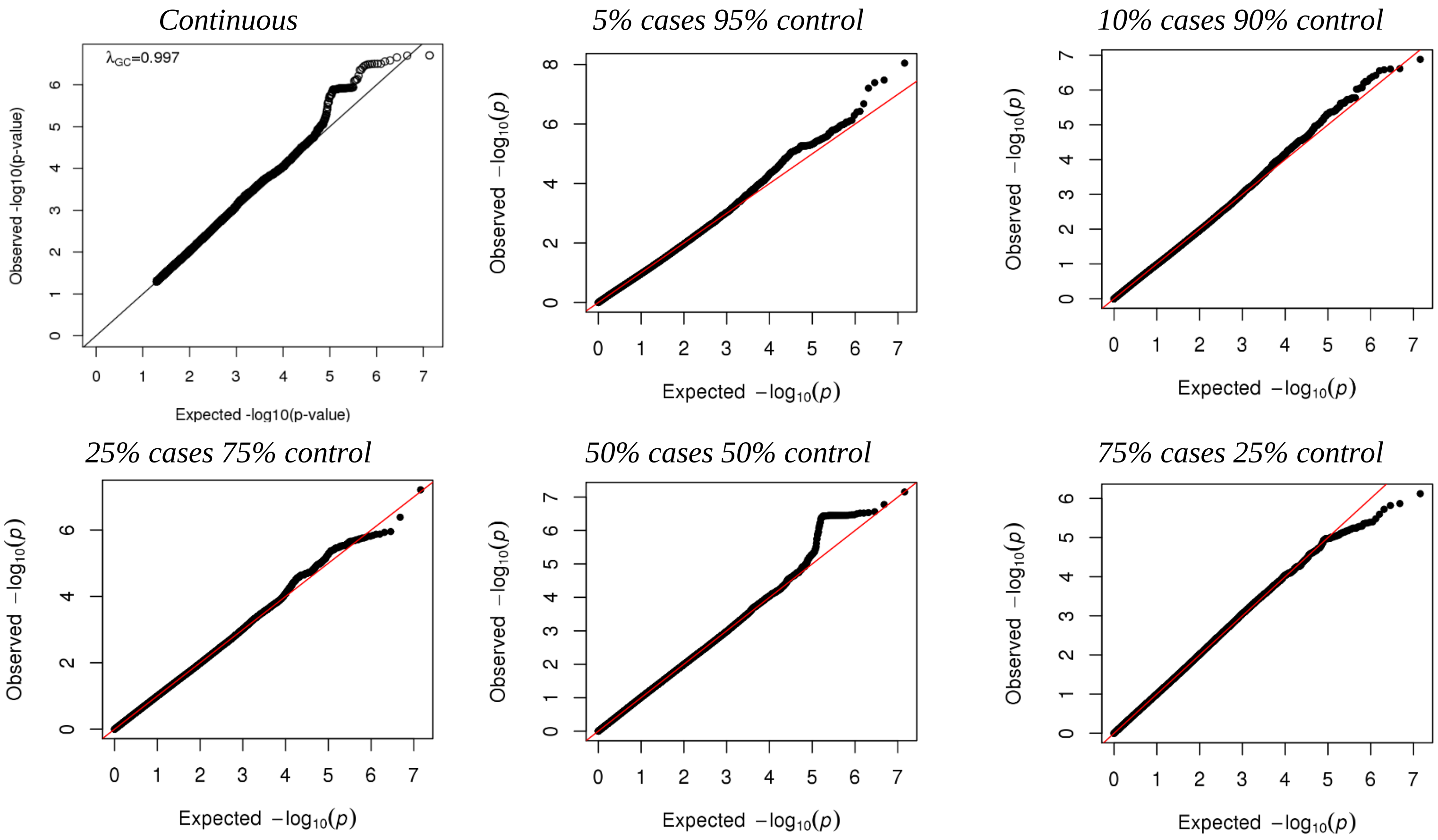

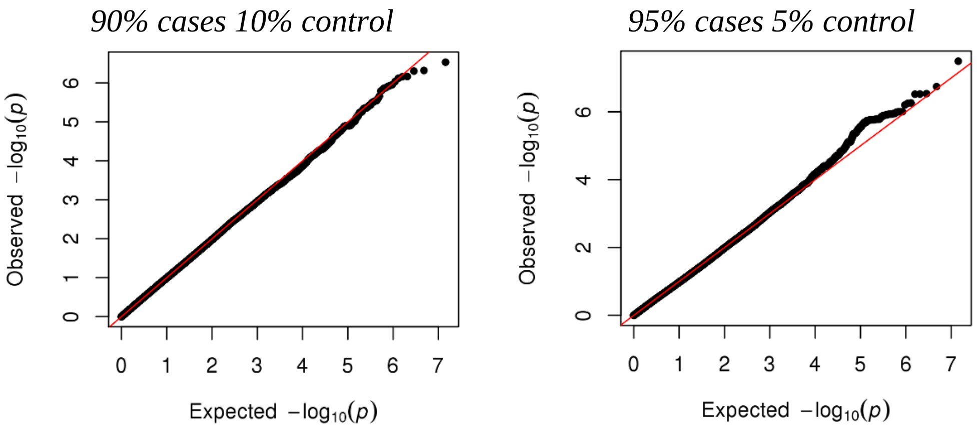


(F) Influenza virus subtype H1N1

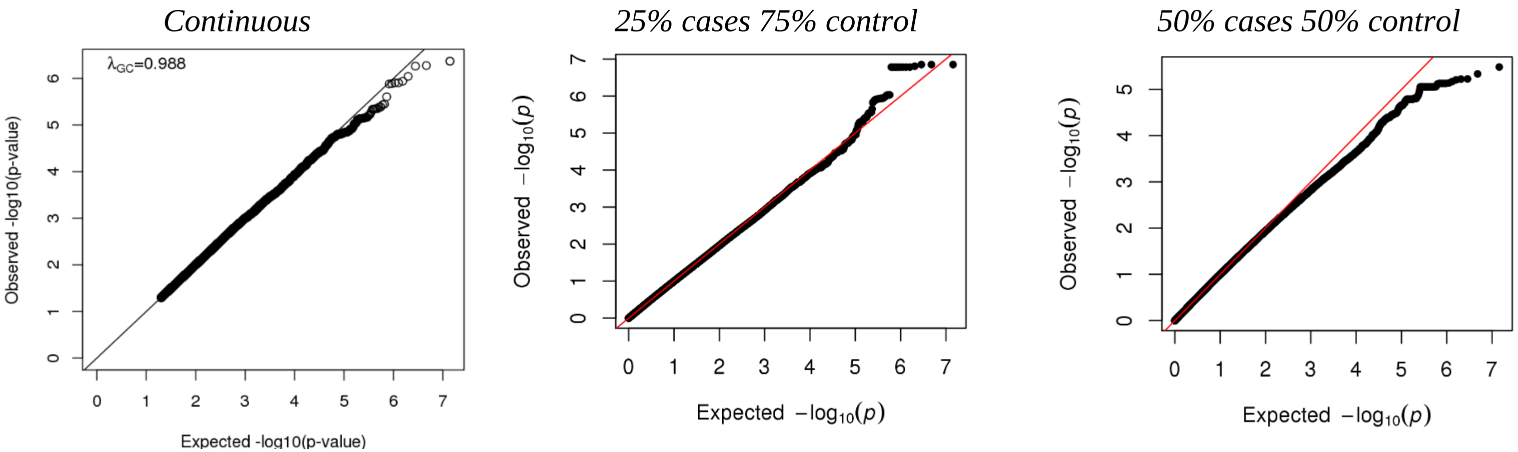

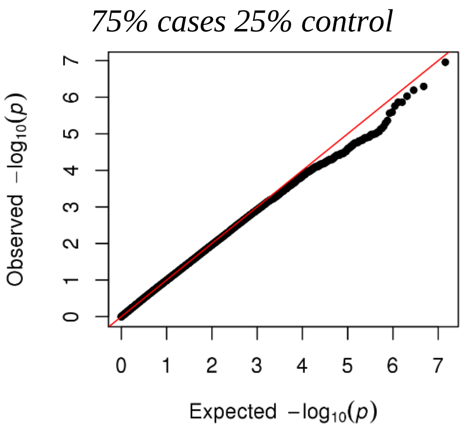


(G) Influenza virus subtype H3N2

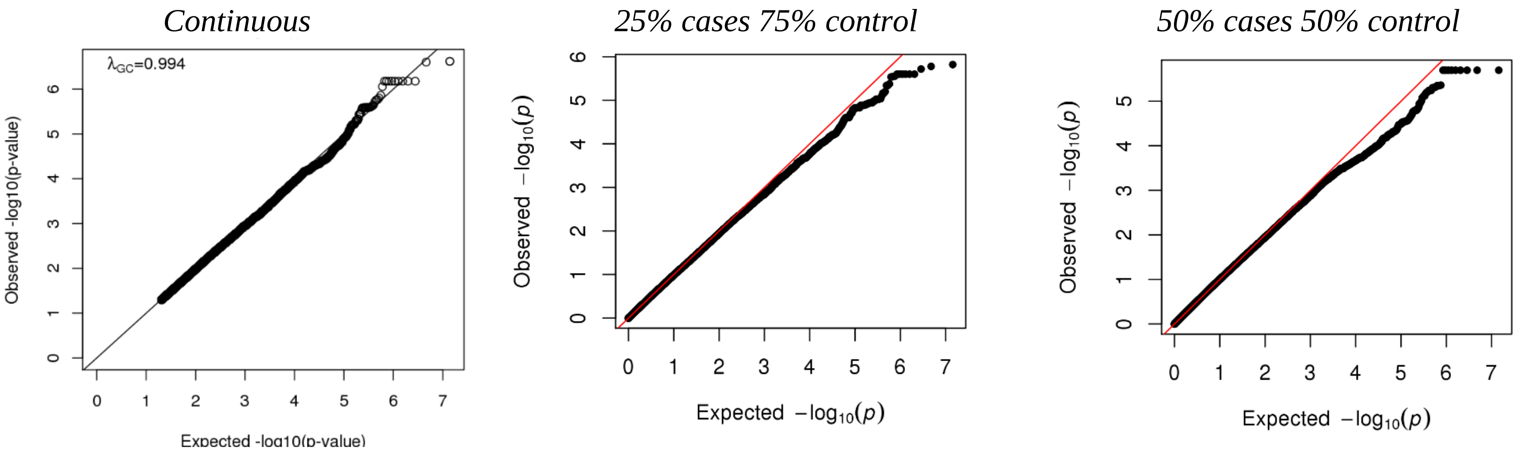

*75% cases 25% control*

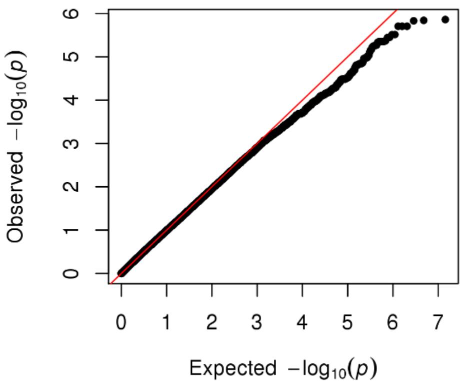


(H) *Helicobacter pylori*

*Continuous*

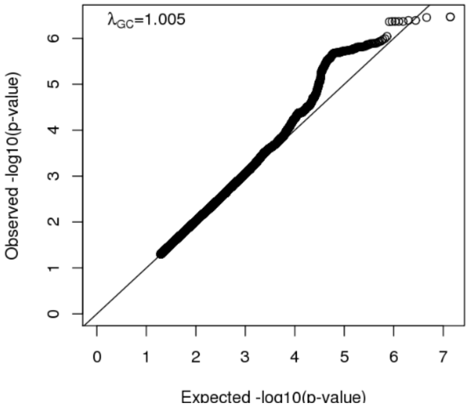


*5% cases 95% control*

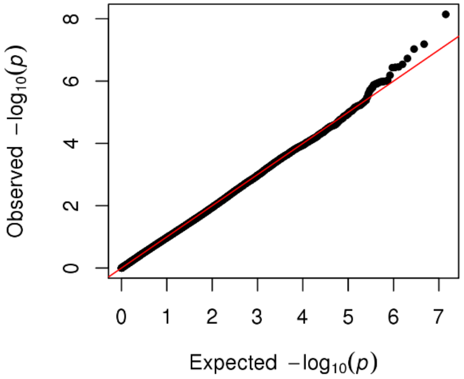


*10% cases 90% control*

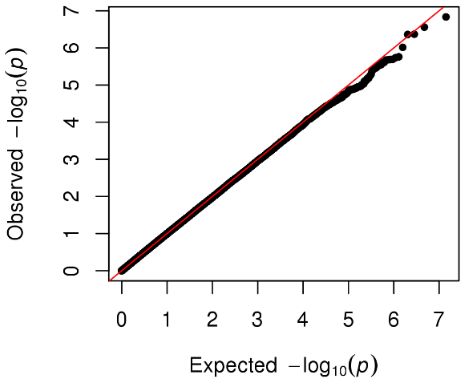

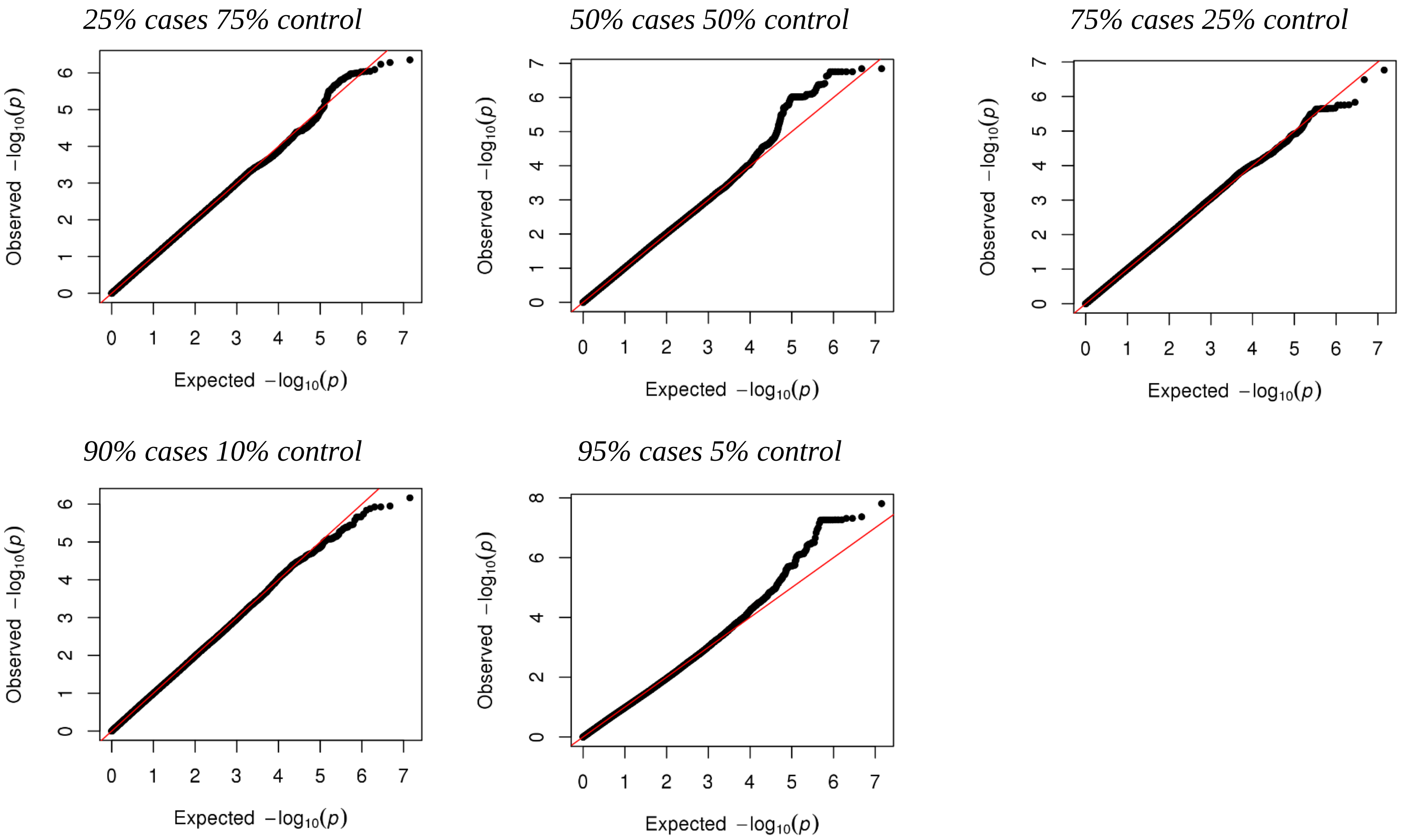
25% cases 75% control
50% cases 50% control
75% cases 25% control
90% cases 10% control
95% cases 5% control
Observed −log10(p)
Expected −log10(p)

(I) Herpes simplex virus 1

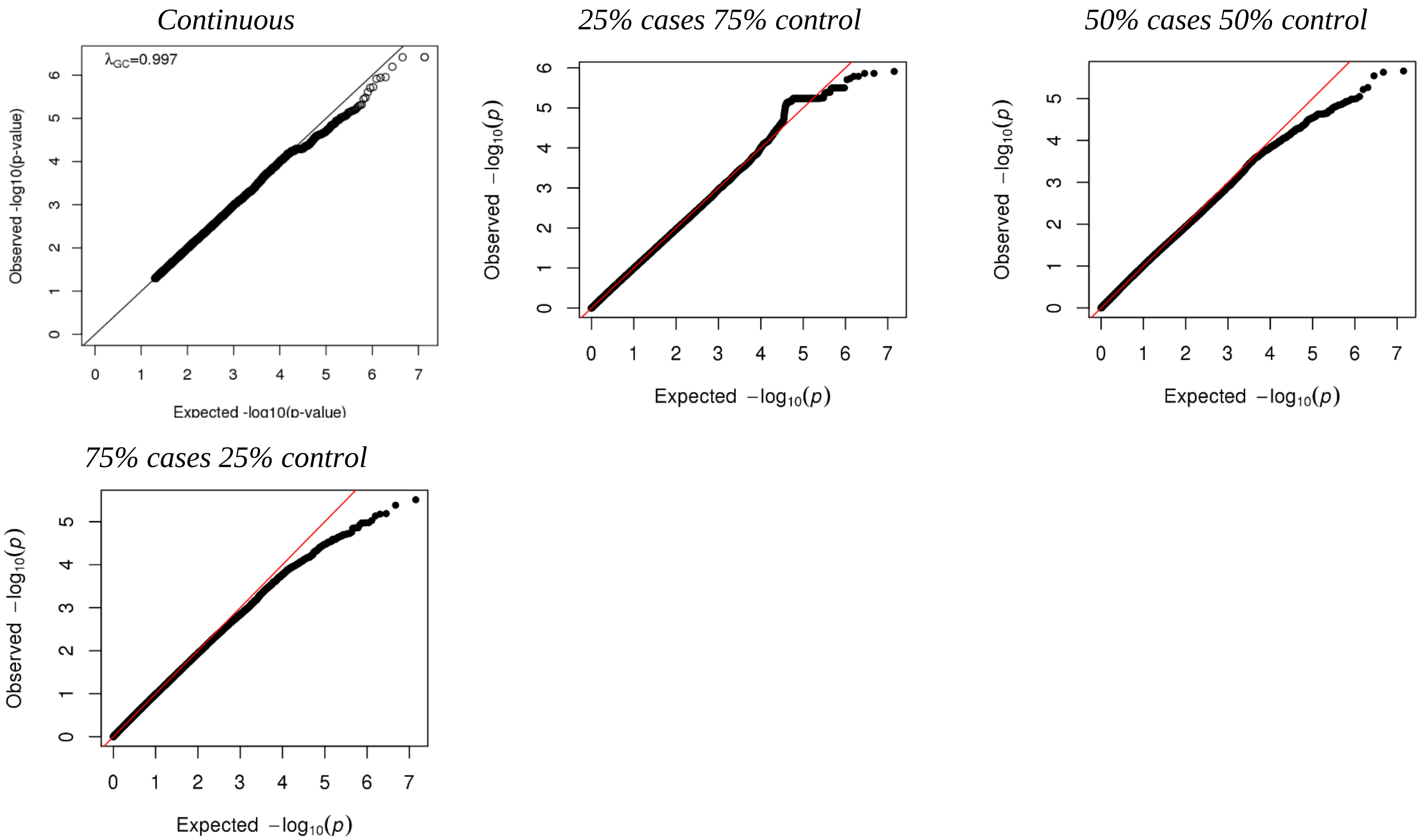

(J) Measles virus

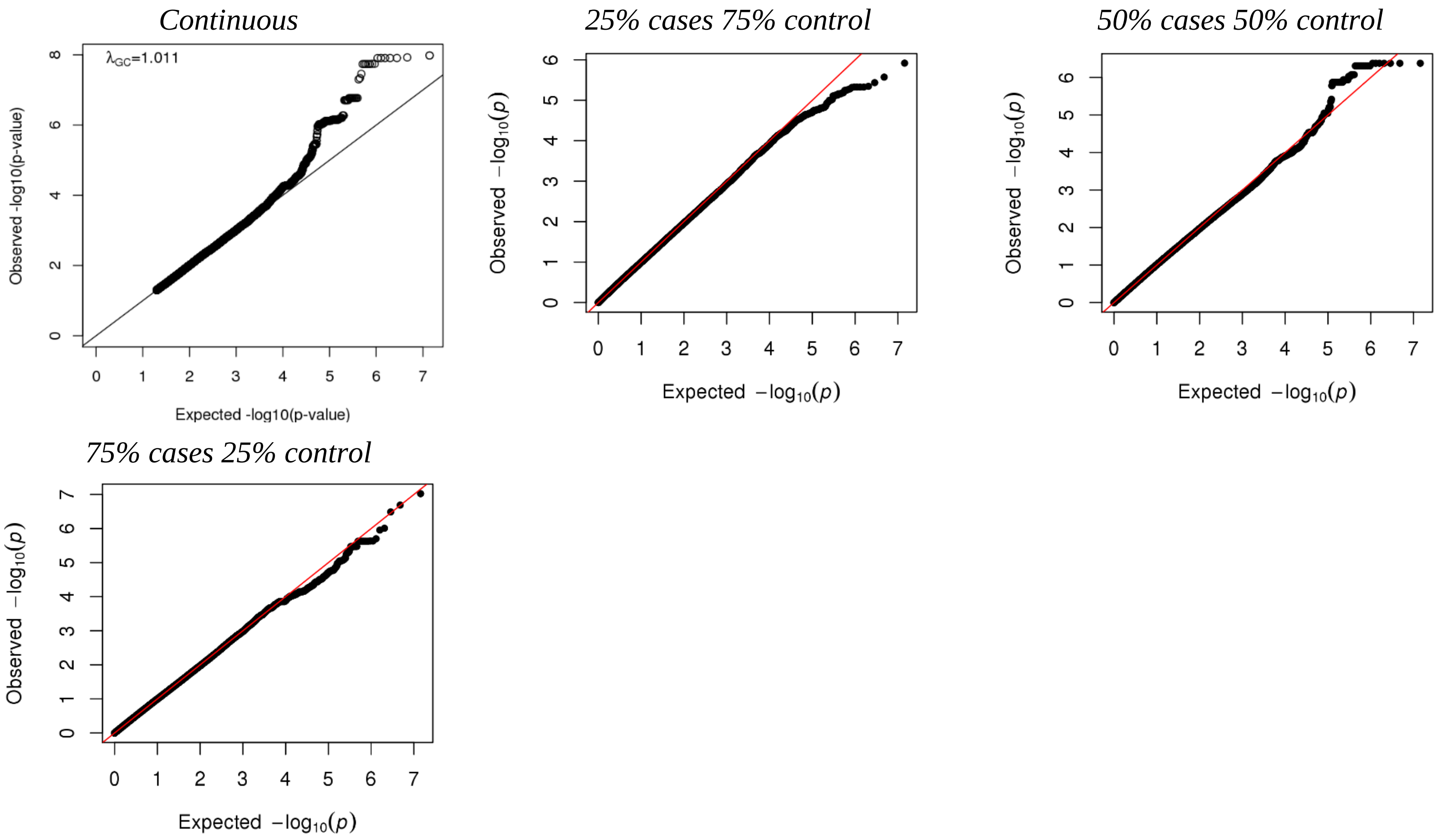

(K) *Saccharomyces cerevisiae*

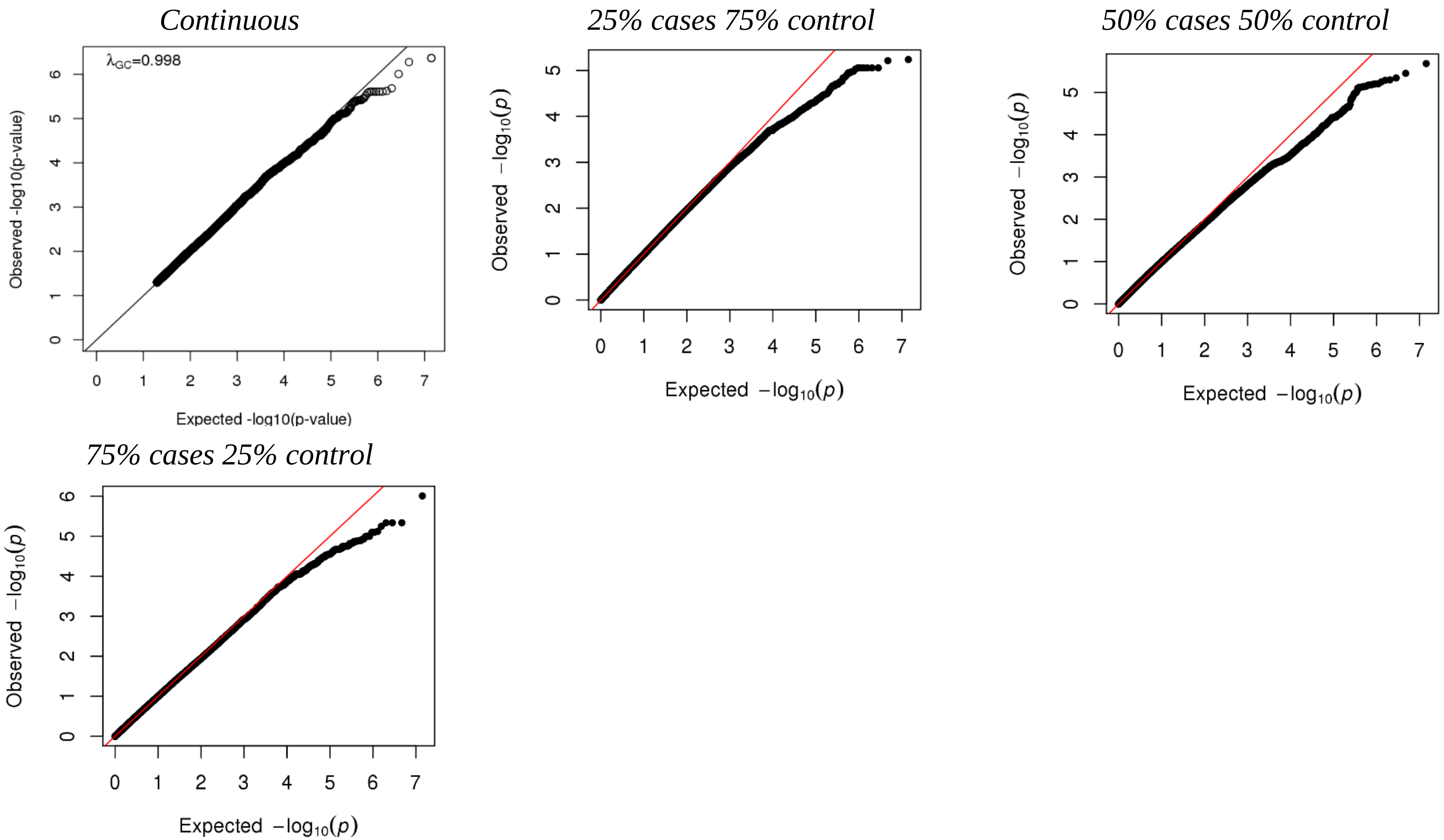

(L) SAG1 protein domain

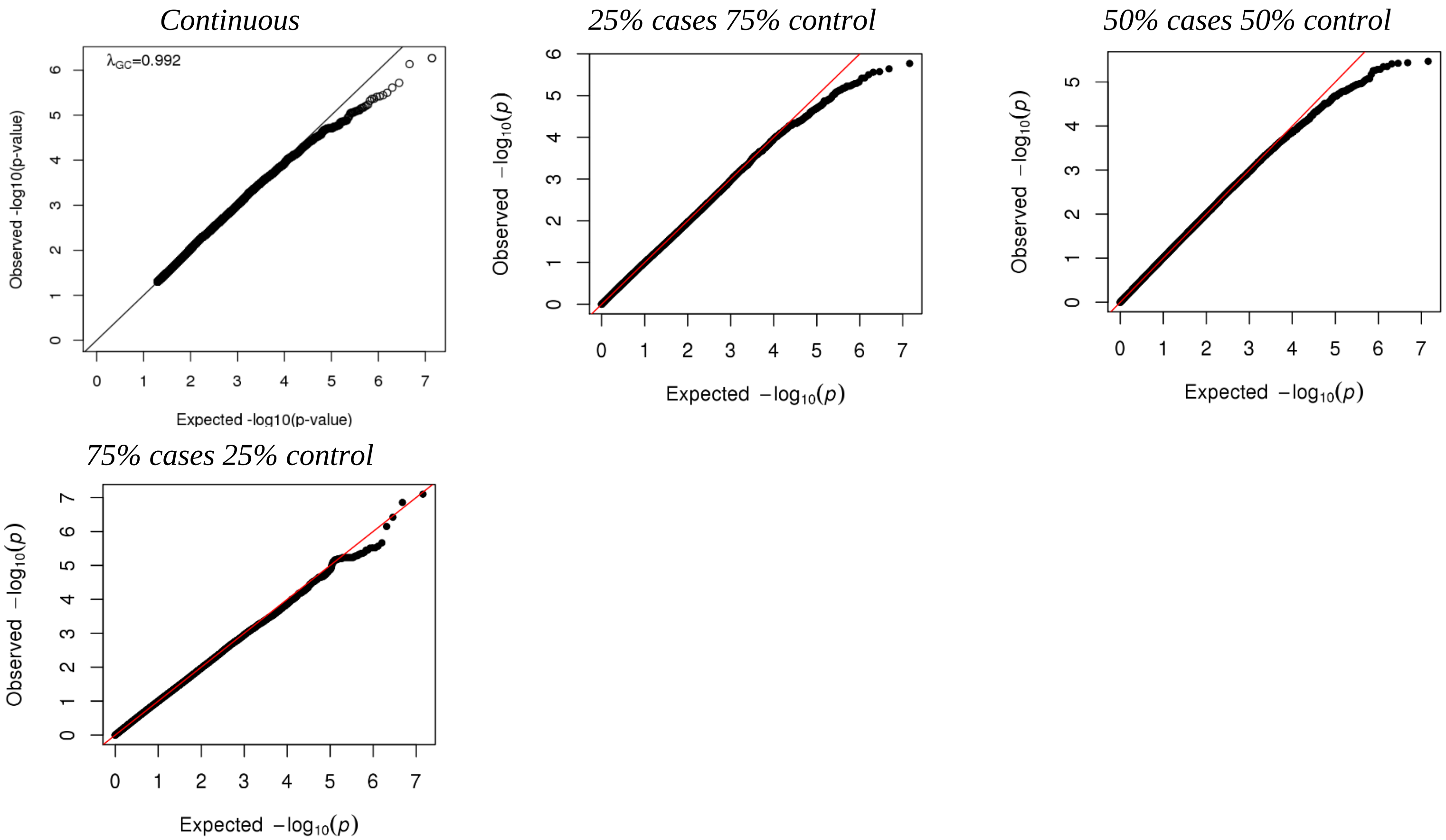

(M) *Toxoplasma gondii*

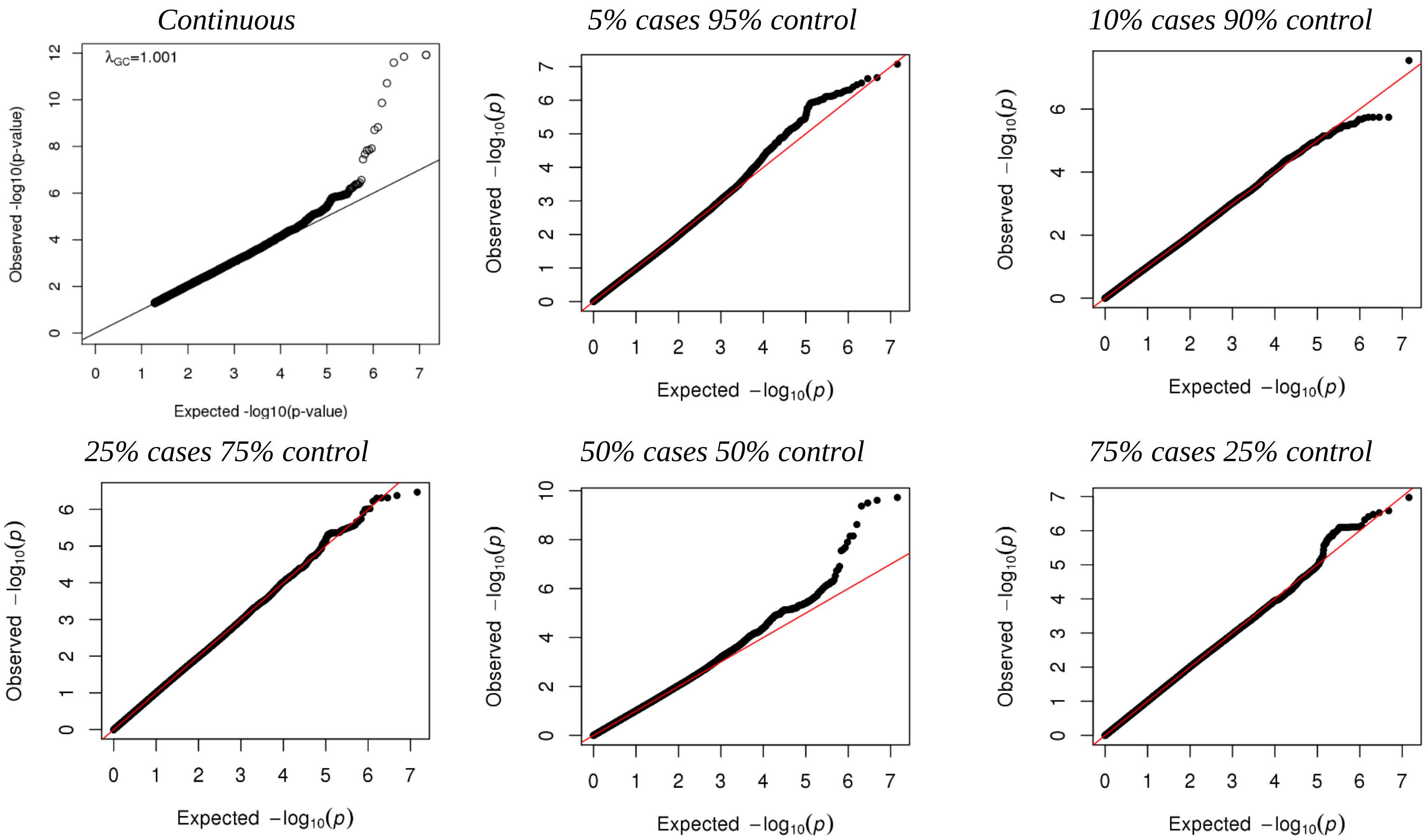

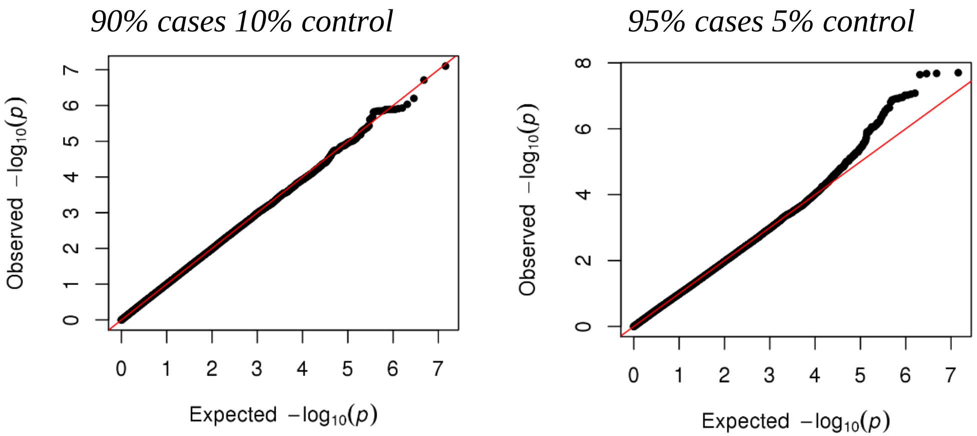


(N) Theiler’s virus

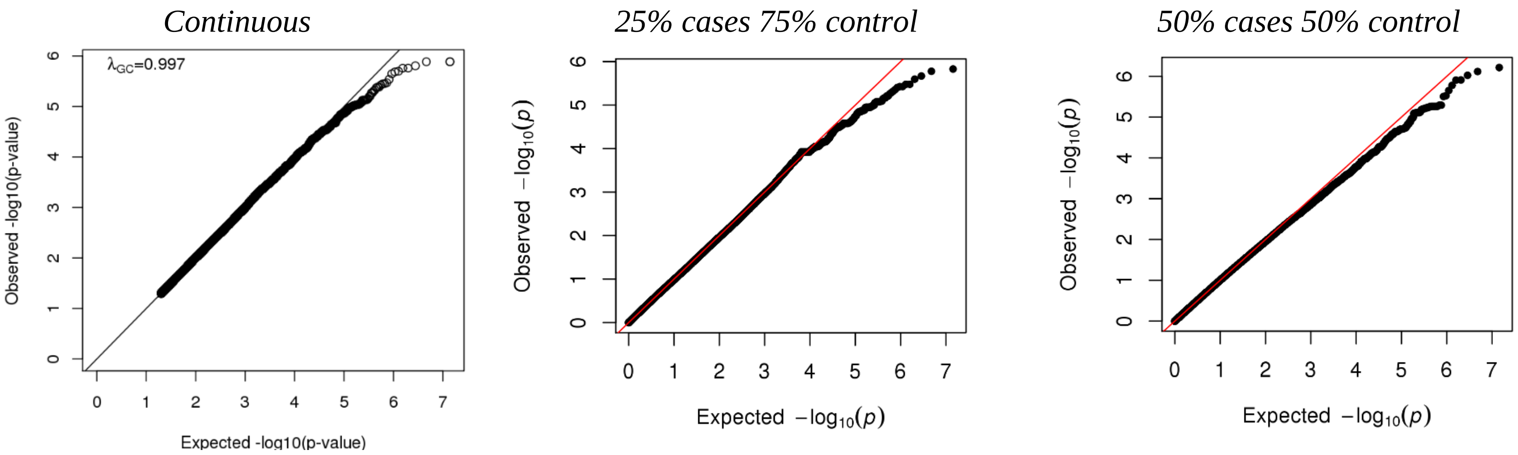

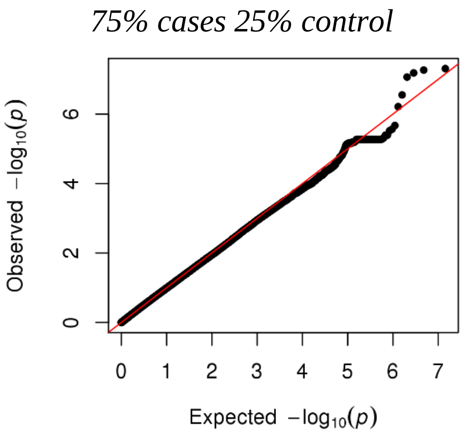


**Supplementary Figure 1** Q-Q plots of several thresholds for antibodies measured at the ALSPAC seven-year clinic.

(A) Alpha-casein protein

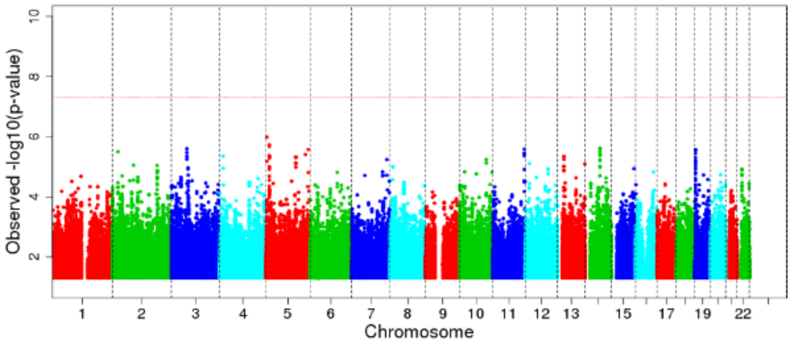


(B) Beta-casein protein

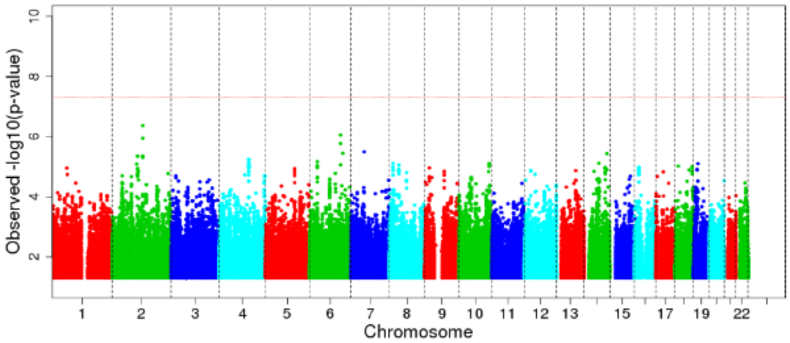


(C) Cytomegalovirus

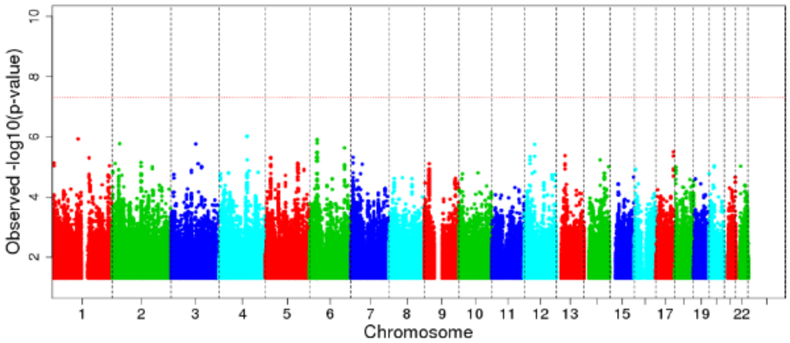


(D) Epstein-Barr virus

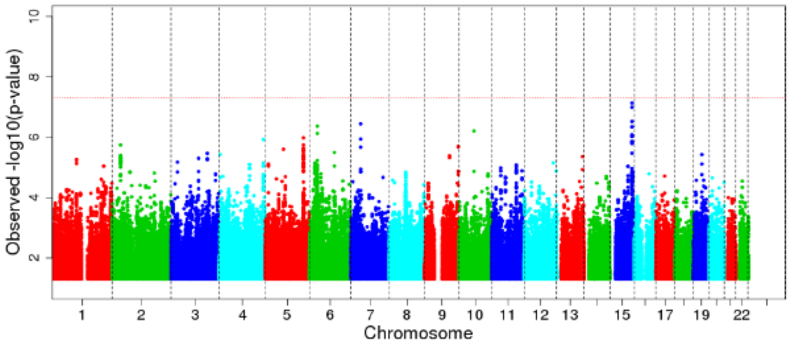


(E) Feline herpes virus

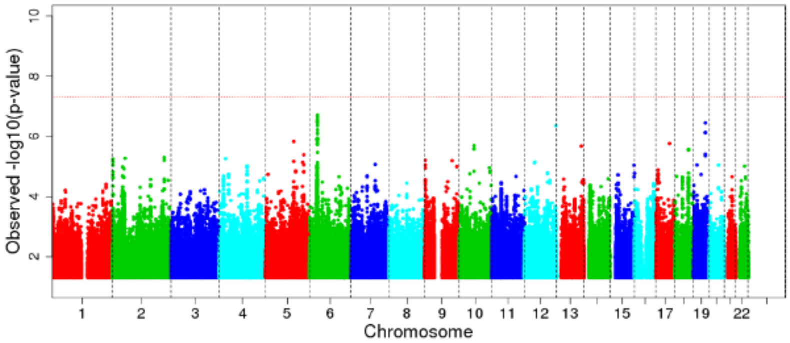

(F) *Helicobacter pylori*

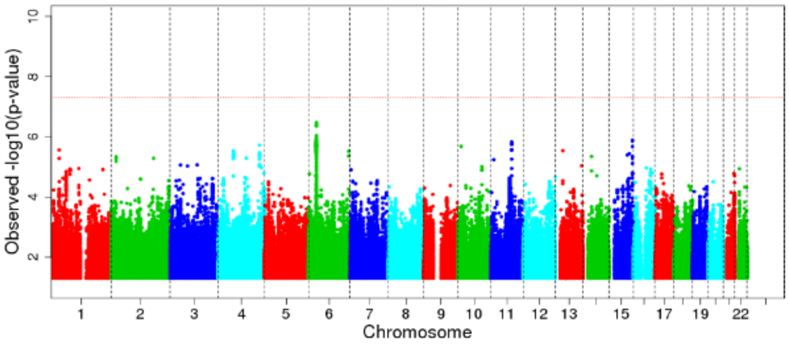


(G) Herpes simplex virus 1

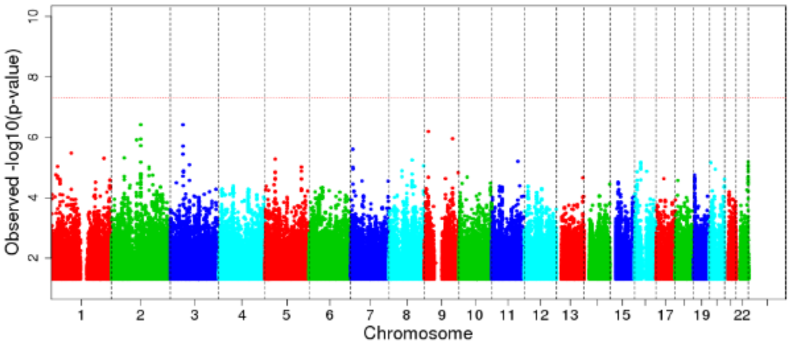


(H) Influenza virus subtype H1N1

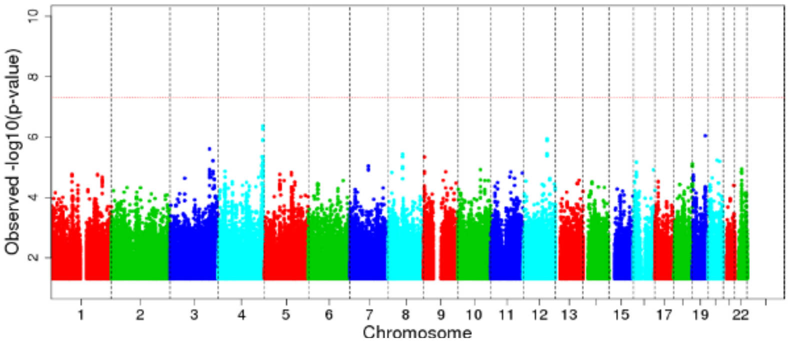


(I) Influenza virus subtype H3N2

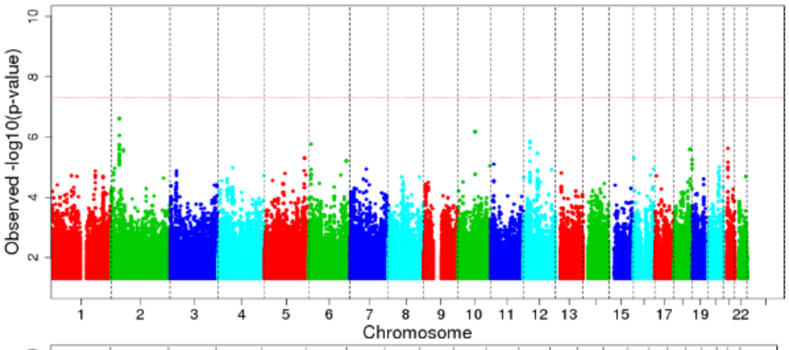


(J) Measles virus

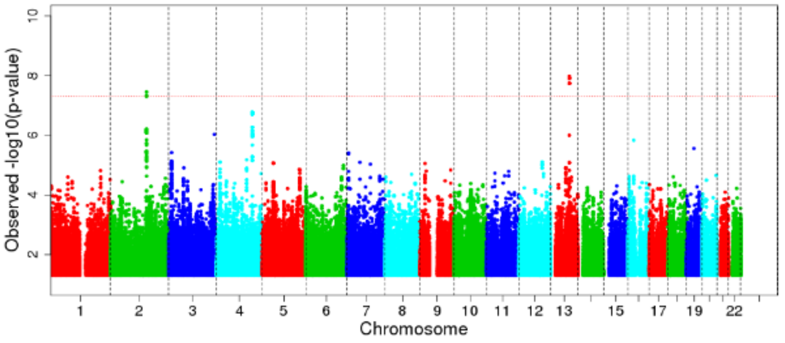

(K) *Saccharomyces cerevisiae*

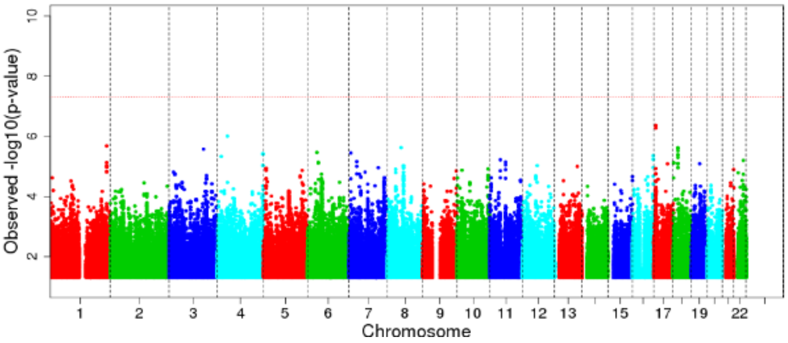


(L) SAG1 protein domain

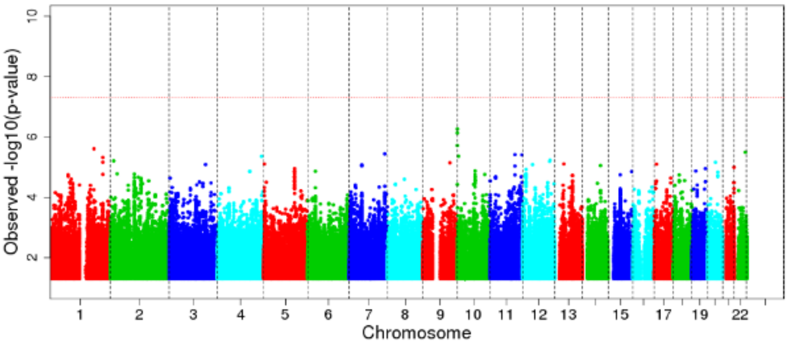


(M) *Toxoplasma gondii*

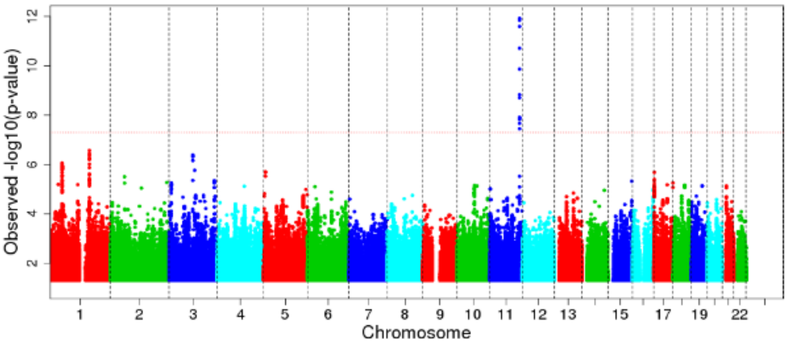


(N) Theiler's virus

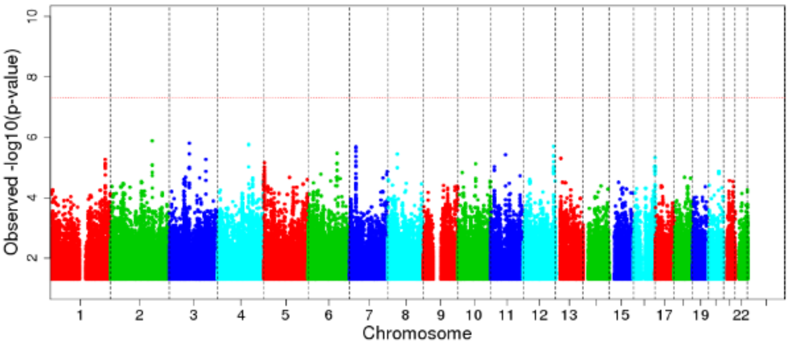


**Supplementary Figure 2** Manhattan plots of rank inverse normal transformed antibodies measured at the ALSPAC seven-year clinic.

(A) Alpha-casein protein

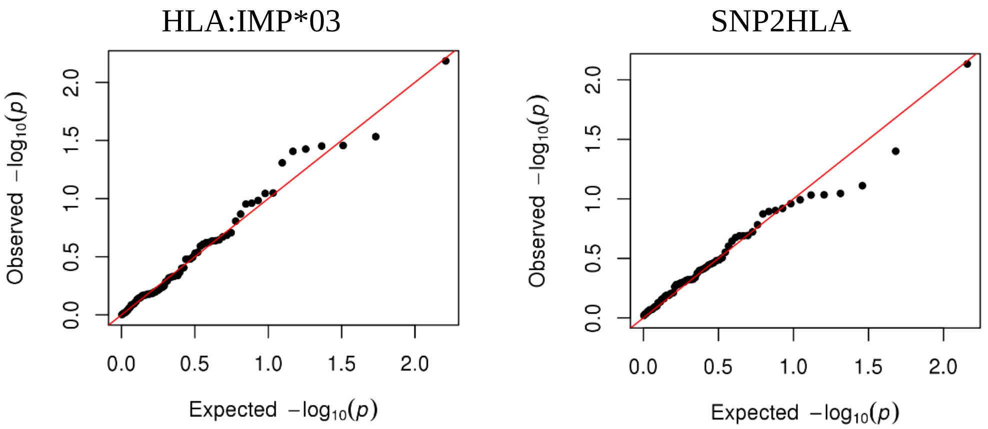


(B) Beta-casein protein

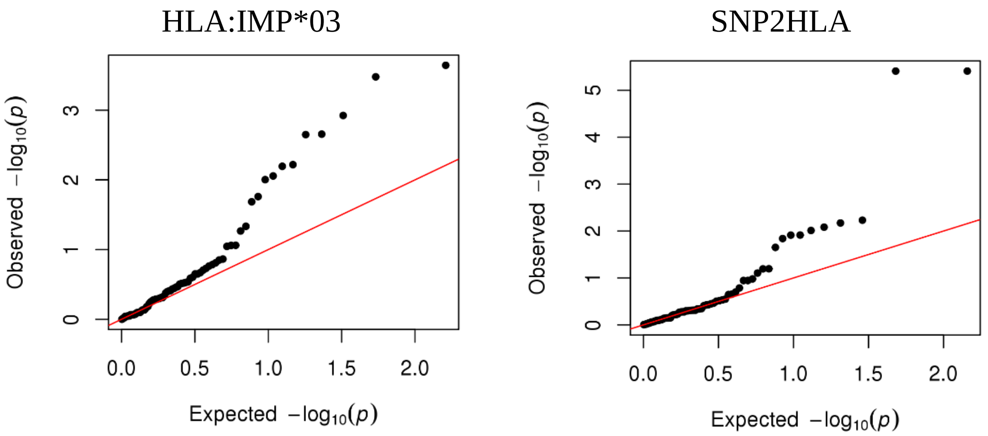


(C) Cytomegalovirus

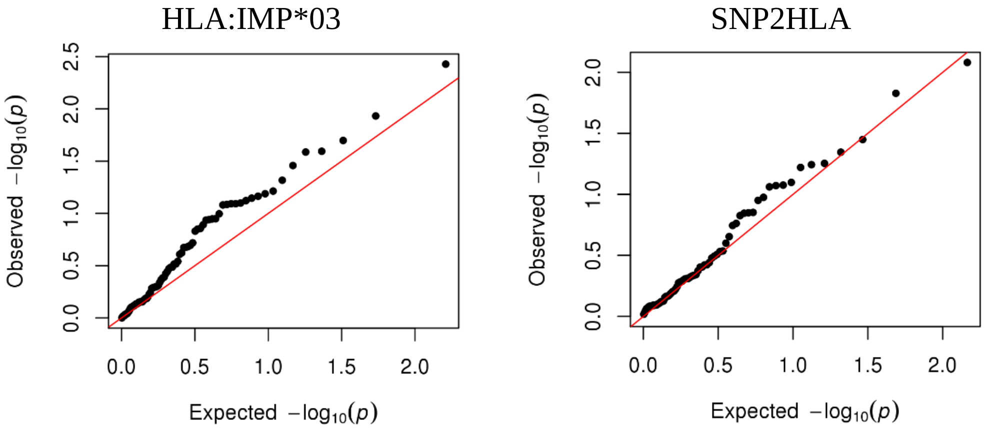


(D) Epstein-Barr virus

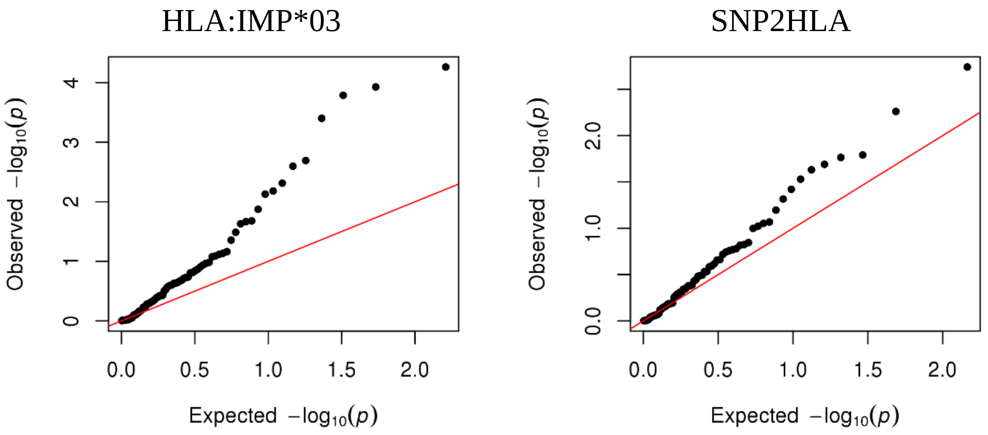

(E) Feline herpes virus

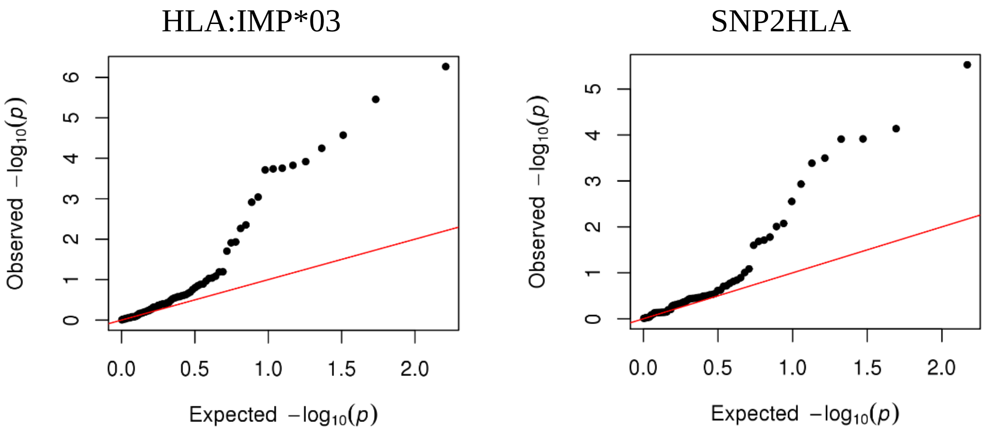


(F) *Helicobacter pylori*

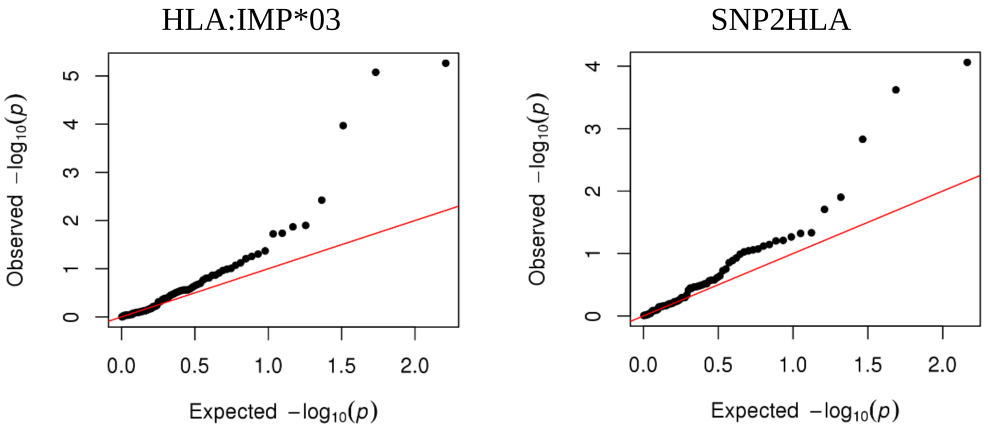


(G) Herpes simplex virus 1

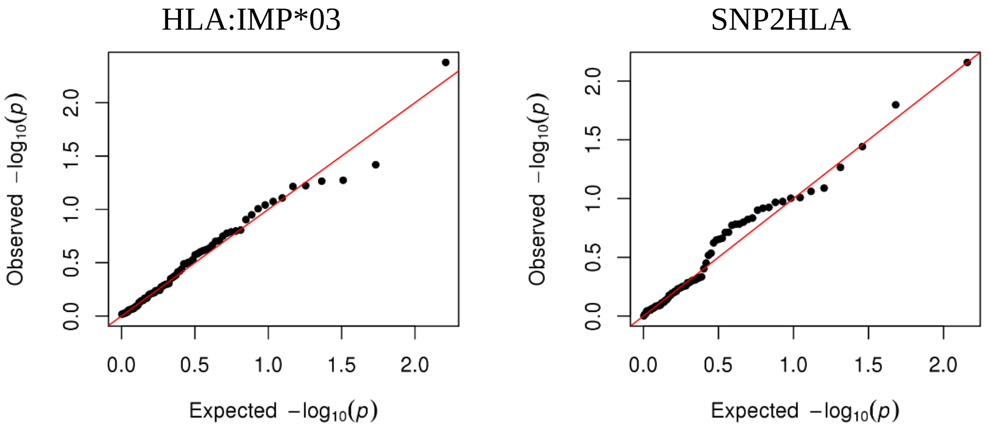


(H) Influenza virus subtype H1N1

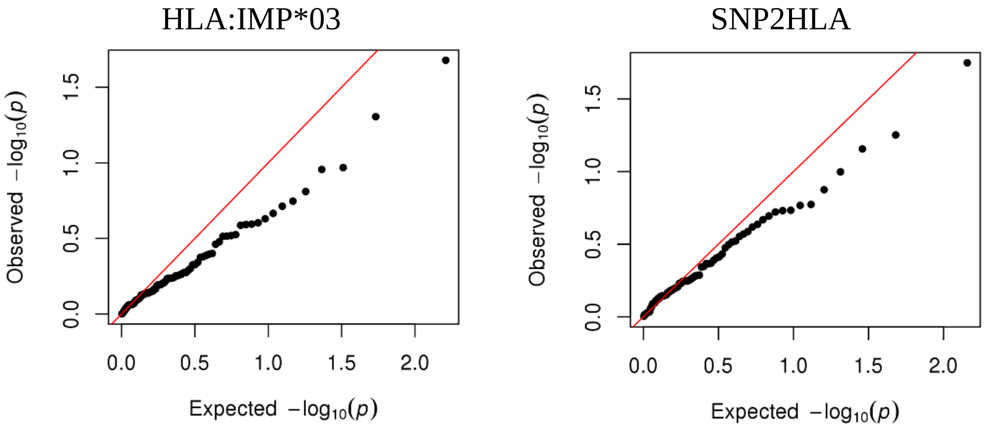

(I) Influenza virus subtype H3N2

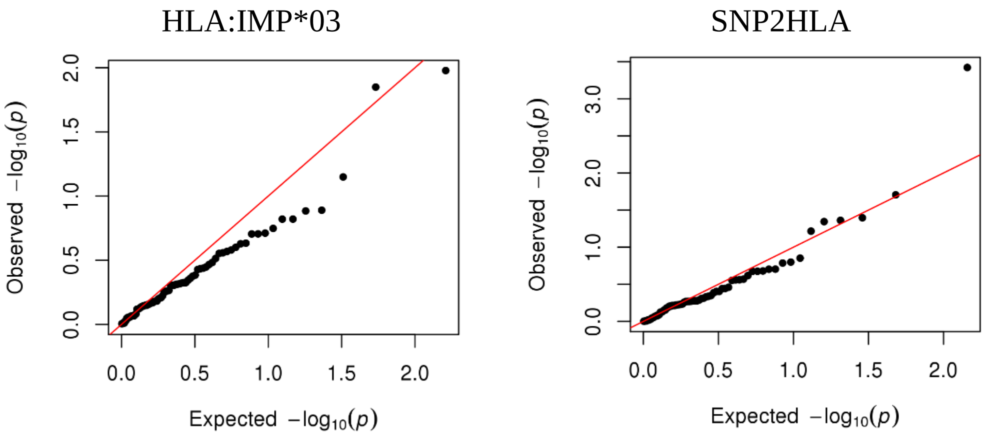


(J) Measles virus

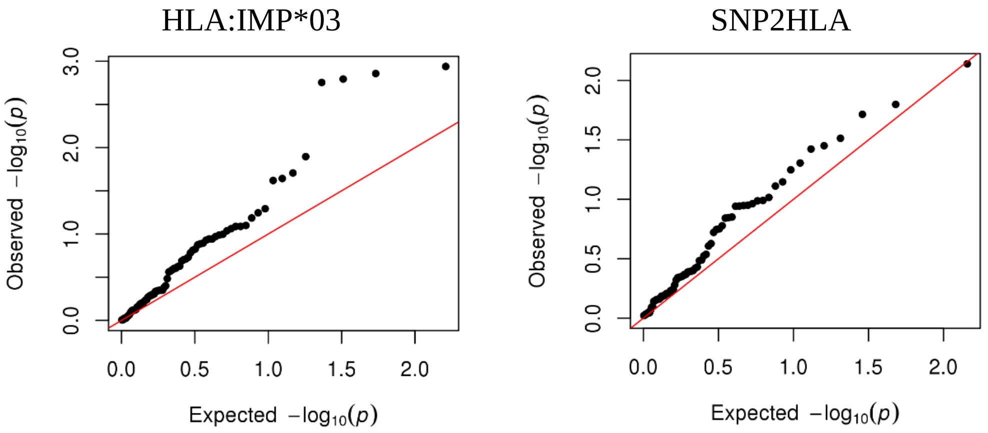


(K) *Saccharomyces cerevisiae*

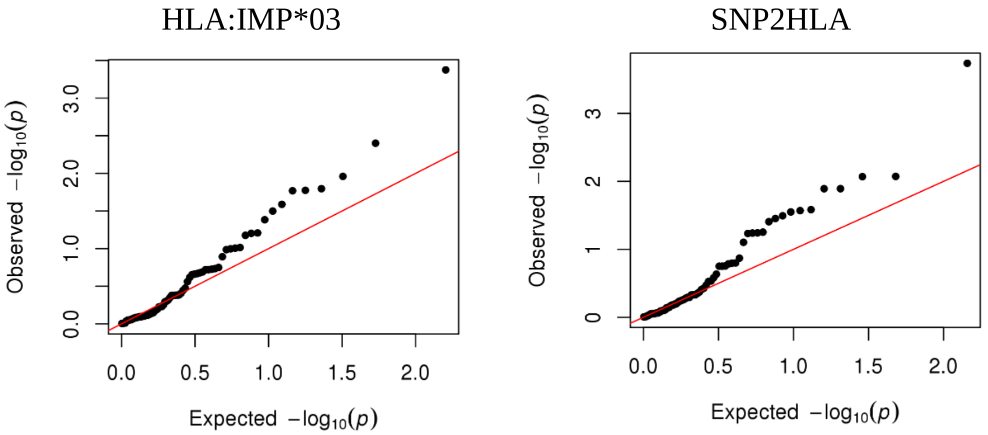


(L) SAG1 protein domain

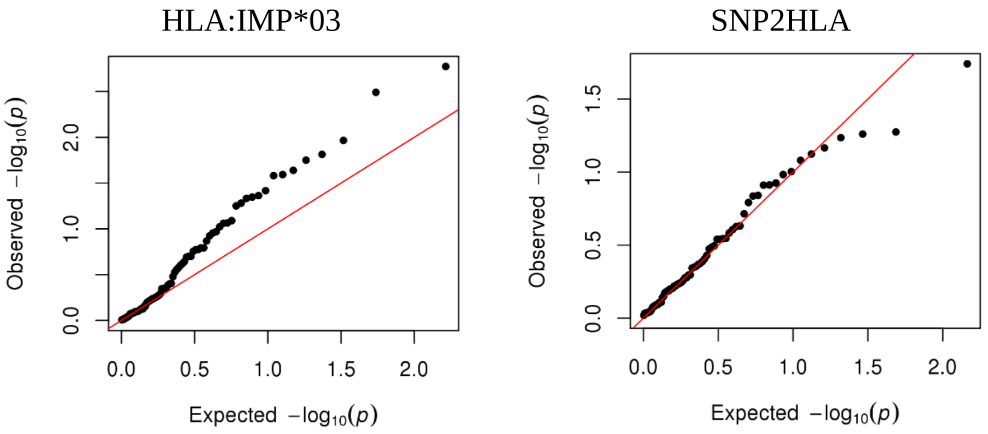

(M) *Toxoplasma gondii*

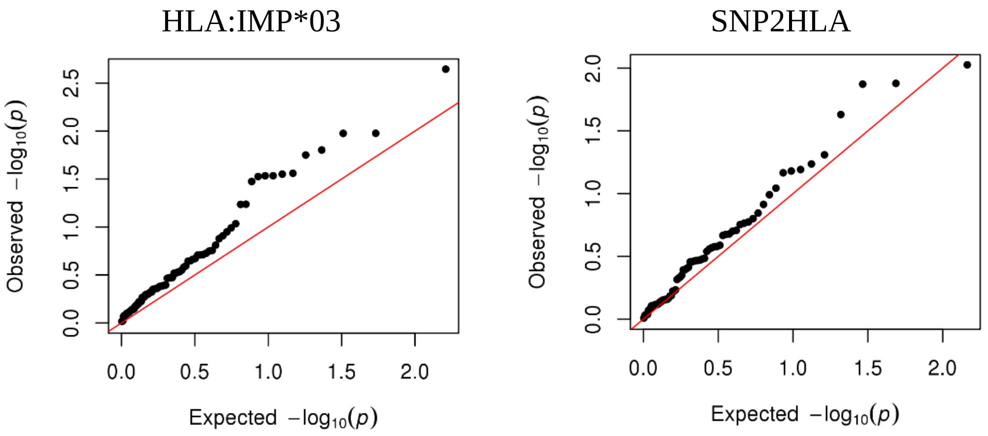


(N) Theiler's virus

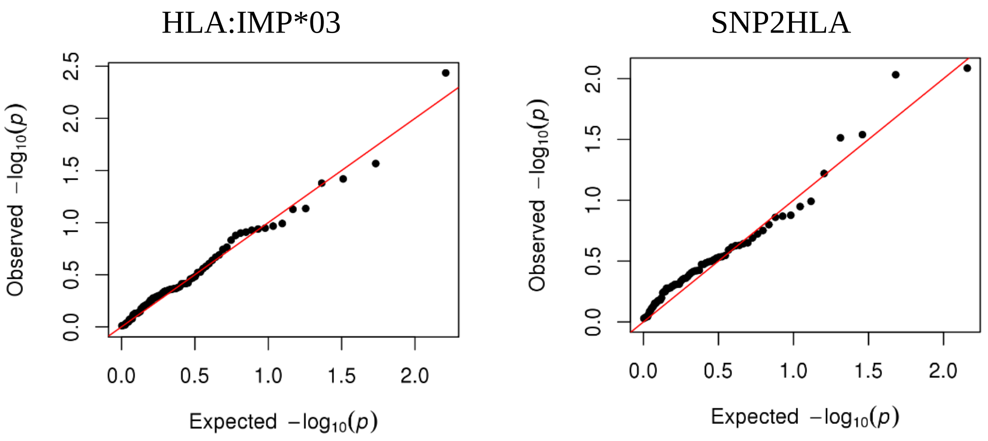


**Supplementary Figure 3** HLA association of antibodies measured at the ALSPAC seven-year clinic: Q-Q plots using two-digit resolution HLA alleles.

(A) Alpha-casein protein

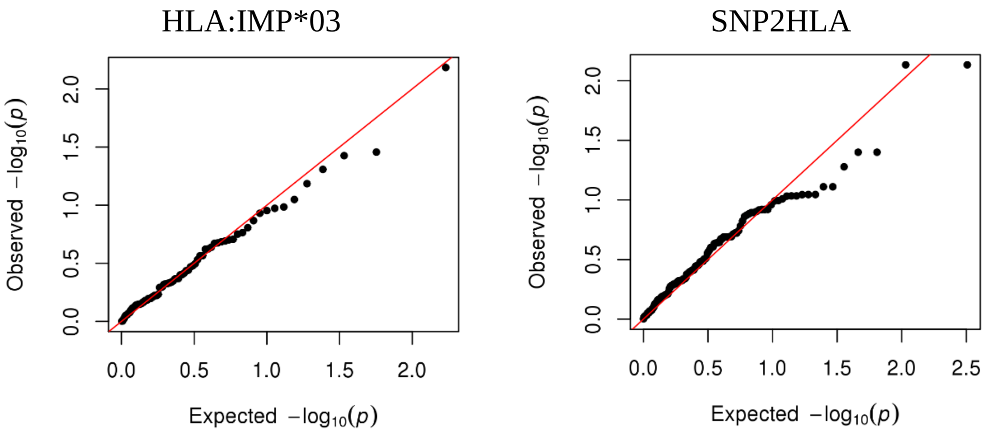


(B) Beta-casein protein

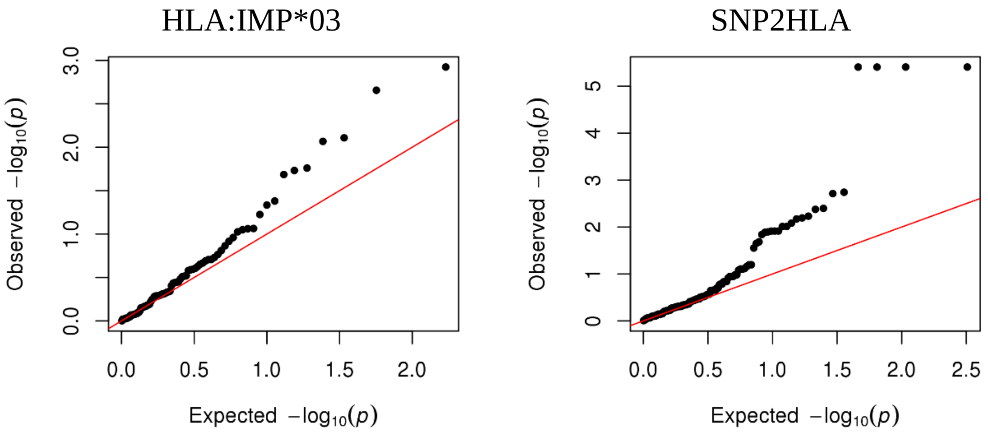


(C) Cytomegalovirus

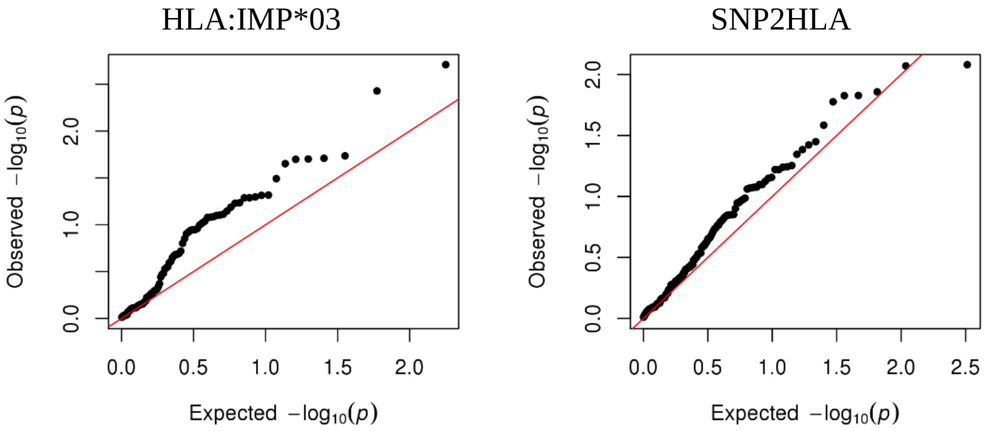


(D) Epstein-Barr virus

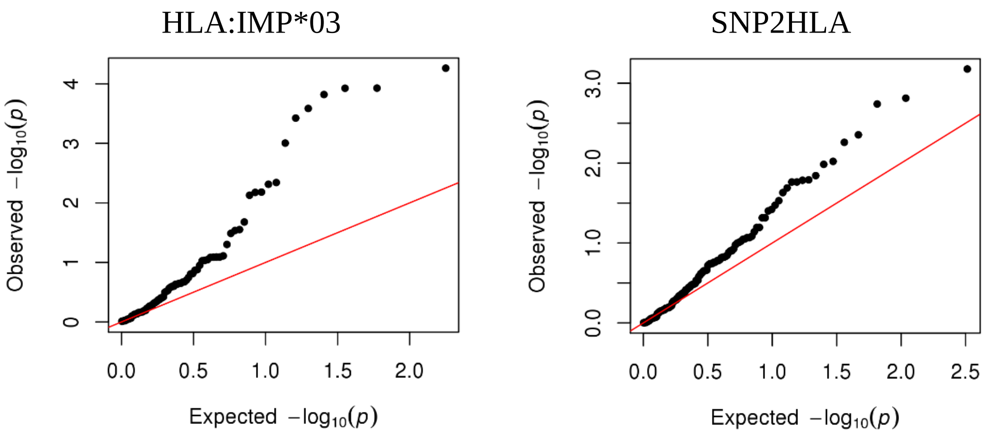

(E) Feline herpes virus

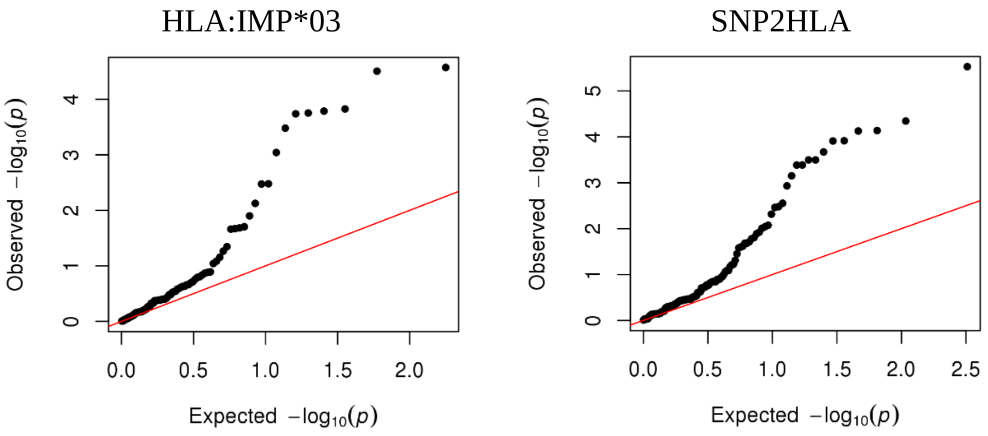


(F) *Helicobacter pylori*

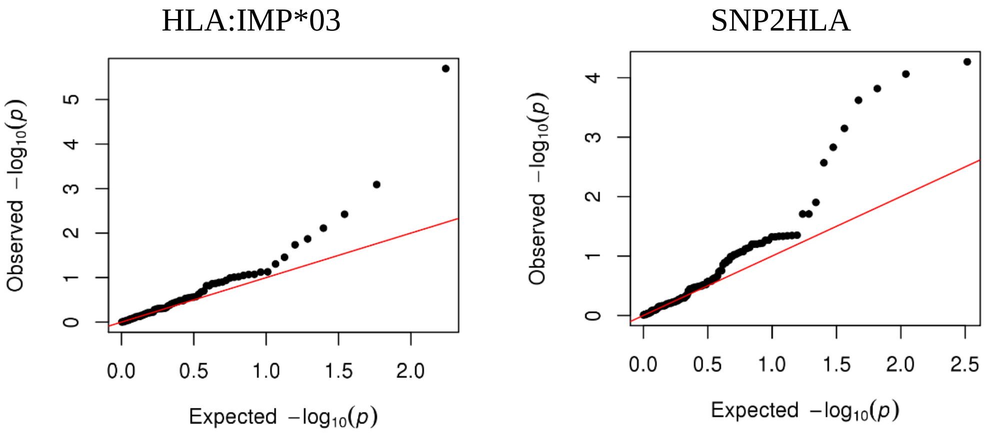


(G) Herpes simplex virus 1

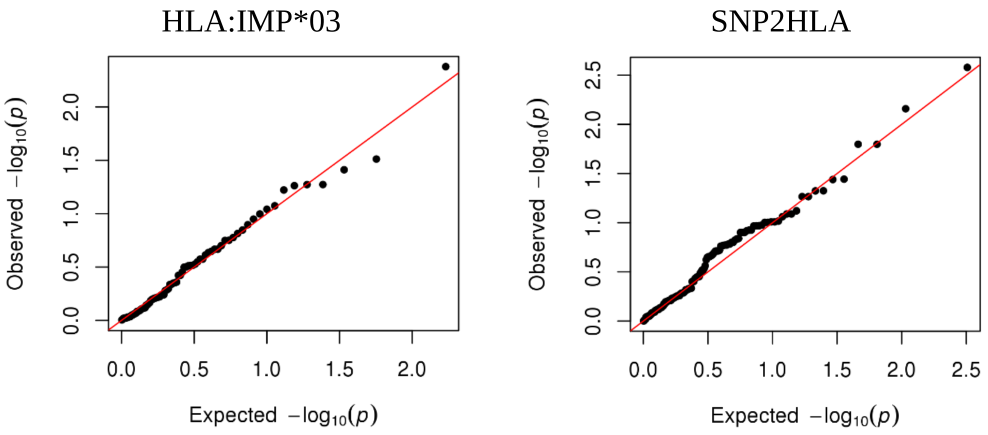


(H) Influenza virus subtype H1N1

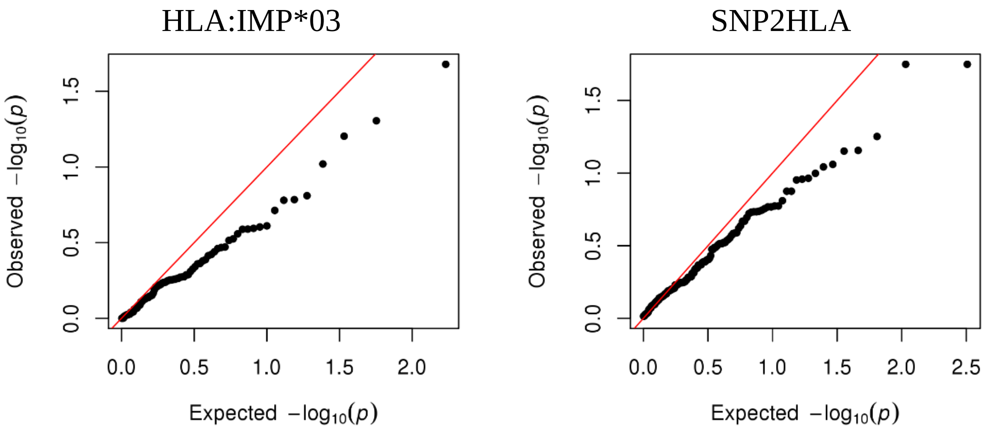

(I) Influenza virus subtype H3N2

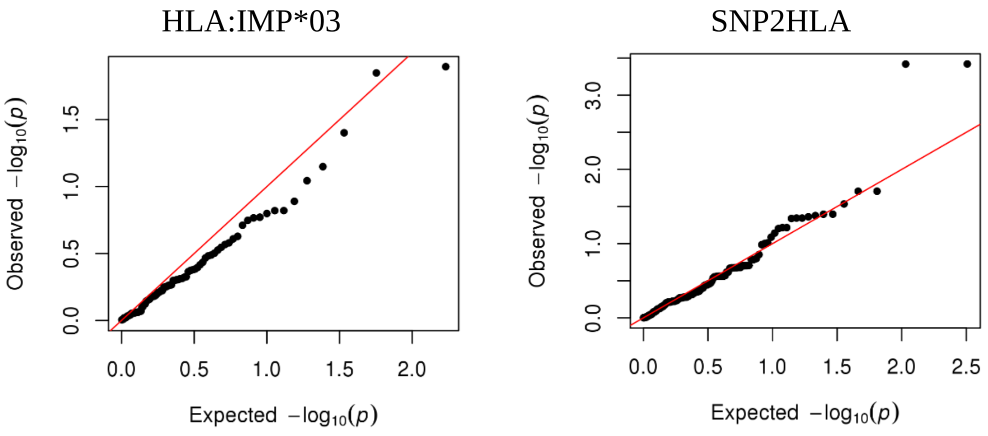


(J) Measles

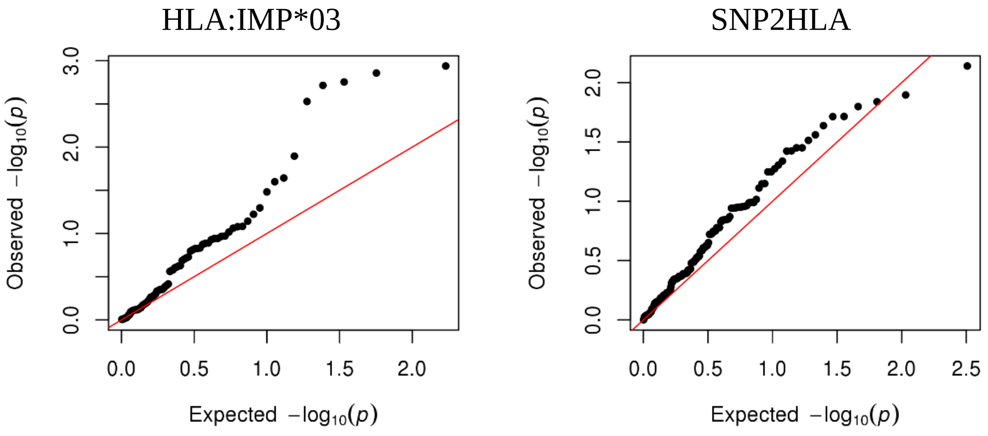


(K) *Saccharomyces cerevisiae*

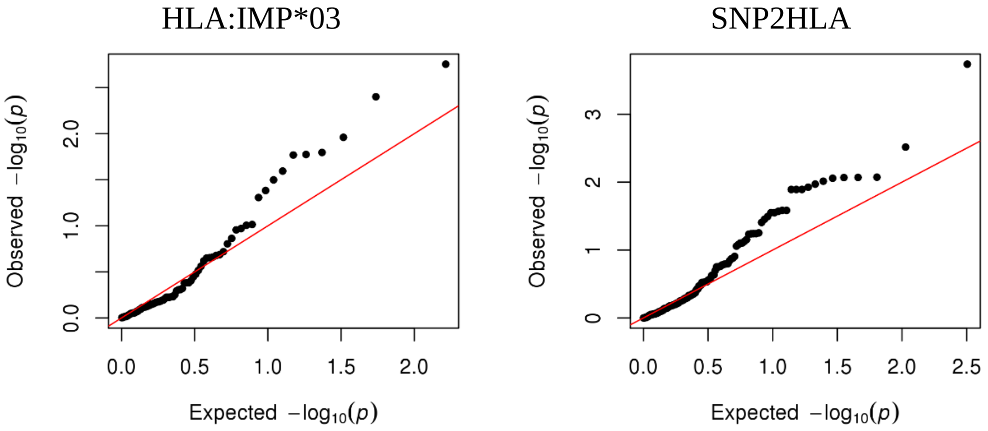


(L) SAG1 protein domain

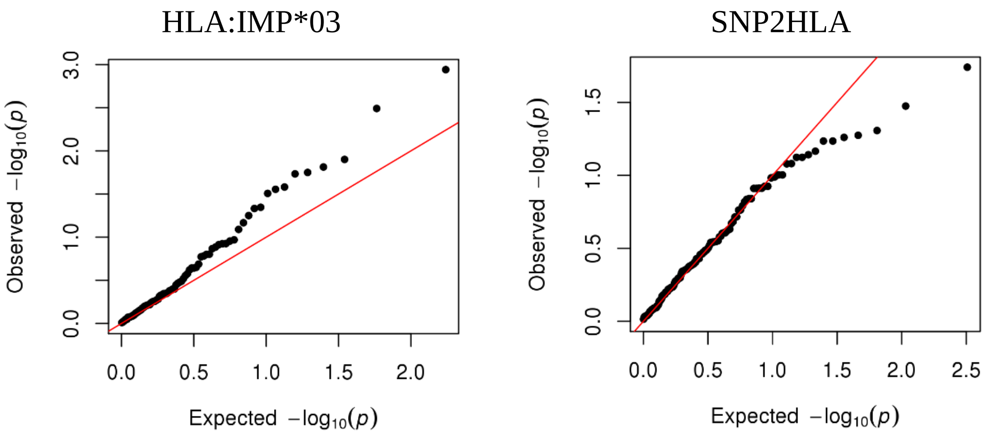

(M) *Toxoplasma gondii*

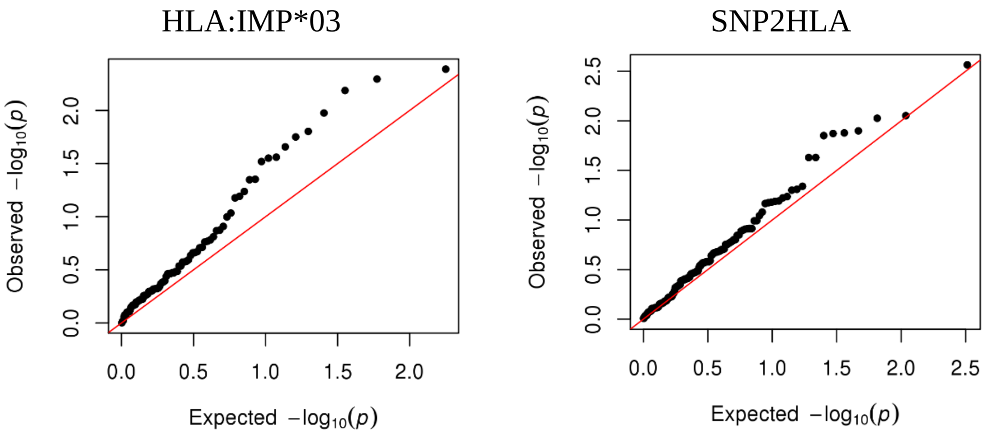


(N) Theiler's virus

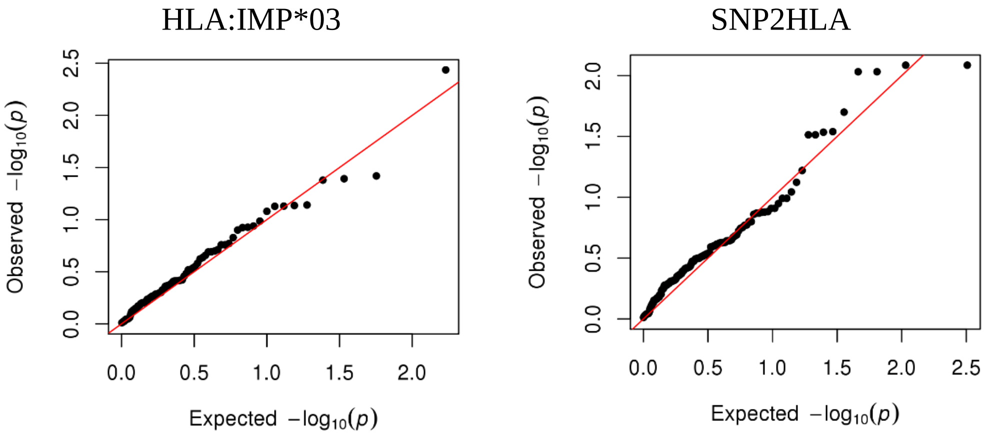


**Supplementary Figure 4** HLA association of antibodies measured at the ALSPAC seven-year clinic: Q-Q plots using four-digit resolution HLA alleles.

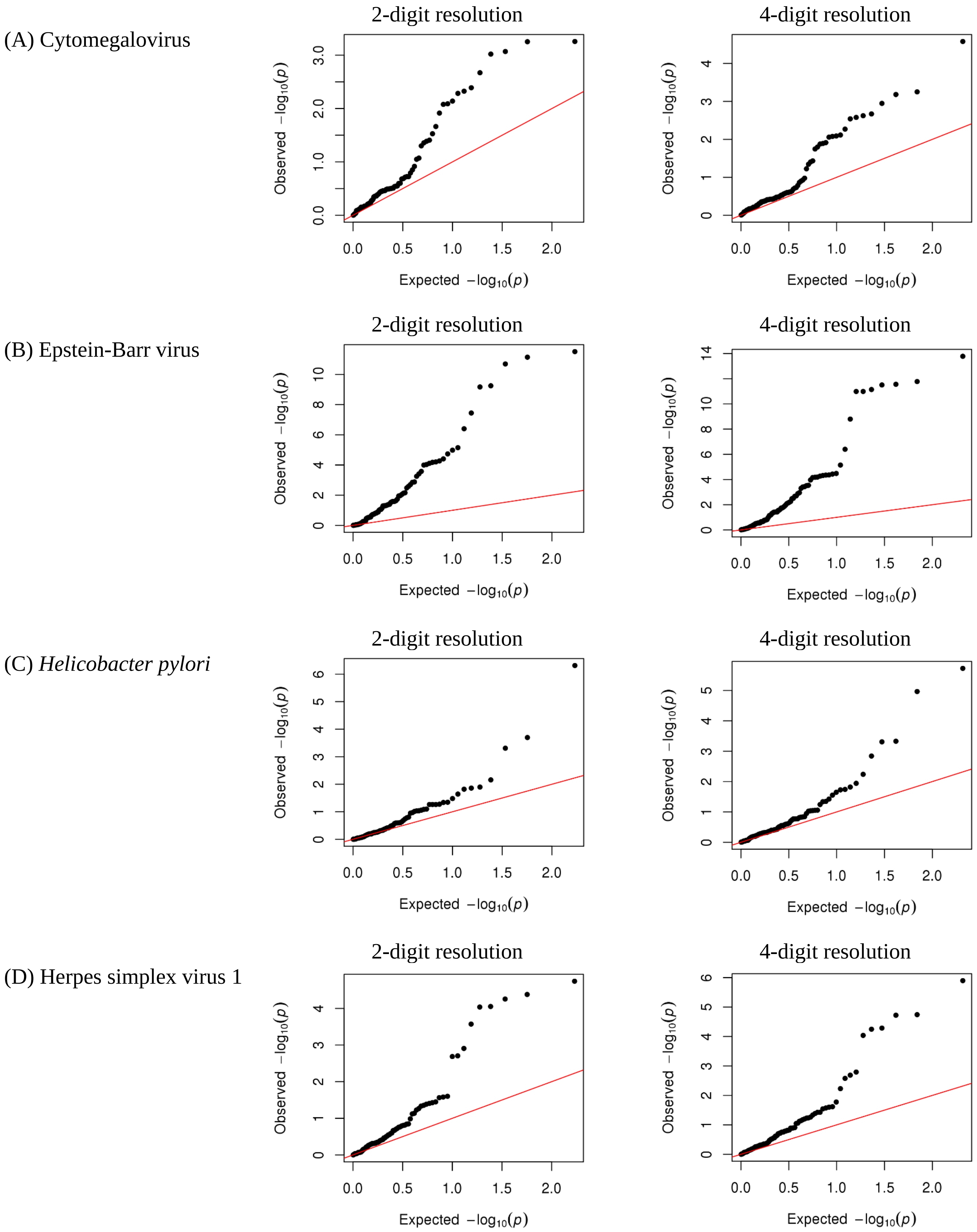
(A) Cytomegalovirus
(B) Epstein-Barr virus
(C) *Helicobacter pylori*
(D) Herpes simplex virus 1
2-digit resolution
4-digit resolution
Observed $-\log_{10}(p)$
Expected $-\log_{10}(p)$

(E) SAG1 protein domain

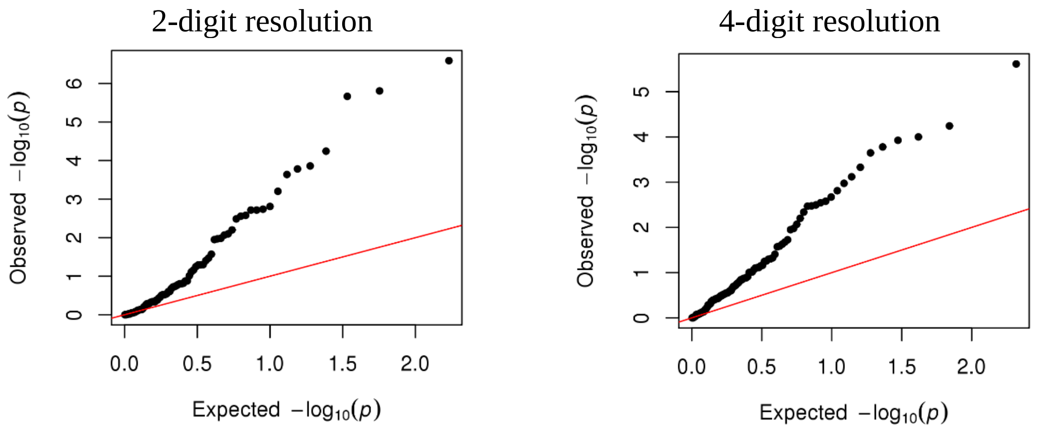


**Supplementary Figure 5** UK Biobank HLA associations of antibodies: Q-Q plots of HLA alleles at 2- and 4-digit resolution.

**Supplementary Tables** (See separate file)

**Table S1. LD clumped SNPs ($P$ > 1×10$^{-6}$) in continuous GWAS for each antibody compared to several thresholded GWASs at the ALSPAC seven-year clinic.** Abbreviations: Beta-casein, Beta-casein protein; CMV, Cytomegalovirus; EBV, Epstein-Barr virus; FHV, Feline herpes virus; *H.pylori*, *Helicobacter pylori*; HSV1, Herpes simplex virus 1; H1N1, Influenza virus subtype H1N1; H3N2, Influenza virus subtype H3N2; Measles, Measles virus; *S.cerevisiae, Saccharomyces cerevisiae*; SAG1, SAG1 protein domain; *T.gondii, Toxoplasma gondii*; Beta, Beta coefficient; log(OR), Logarithm of the odds ratio; L95,Lower 95% confidence interval; U95, Upper 95% confidence interval; P, *p*-value

**Table S2. Replication of genetic signals of measured antibodies from the ALSPAC seven-year clinic at different clinic years ($P$ < 3.5×10$^{-4}$).** SNPs in bold highlight SNPs that exceeded the genome-wide thresholds ($P$ < 5×10$^{-8}$). Abbreviations: Beta, Beta effect estimate for the effect allele; L95, Lower 95% confidence interval; U95, Upper 95% confidence interval; *T.gondii*, *Toxoplasma gondii*; Beta-casein, Beta-casein protein; CMV, Cytomegalovirus; EBV, Epstein-Barr virus; FHV, Feline herpes virus; *H.pylori*, *Helicobacter pylori*; HSV1, Herpes simplex virus 1; H1N1, Influenza virus subtype H1N1; H3N2, Influenza virus subtype H3N2; Measles, Measles virus; *S.cerevisiae*, *Saccharomyces cerevisiae*; SAG1, SAG1 protein domain; *T.gondii*, *Toxoplasma gondii*.

**Table S3. Top SNPs in discovery GWAS in UK Biobank with replication GWAS in ALSPAC seven-year clinic.** Abbreviations: EBV, Epstein-Barr virus; HSV1, Herpes simplex virus 1; EA, Effect allele; Beta, Effect estimate for the effect allele; L95, Lower 95% confidence interval; U95, Upper 95% confidence interval; P, *p*-value. (-) Denotes SNPs missing due to QC filtering; NA denotes SNPs not present in reference panel.

**Table S4. Look-up of the three genome-wide and 26 suggestive genetic signals in ALSPAC associated with measured antibodies across the SARS-CoV-2 GWAS meta-analysis (C2_ALL_eur) ($P$ < 0.002).** The ALSPAC genome-wide and suggestive SNPs are shown in the left-hand column, followed by the results from the SARS-CoV-2 (C2_ALL_eur) GWAS meta-analysis performed by the COVID-19 Host Genetics Initiative in the right-hand column. Abbreviations: EA, Effect allele; Beta, Effect estimate for the effect allele; L95, Lower 95% confidence interval; U95, Upper 95% confidence interval; P, *p*-value. (-) denotes SNPs that were not present in the reference panel or removed due to quality control filtering.

**Table S5. Look-up of the seven SNPs associated with SARS-CoV-2 antibodies across the 14 ALSPAC antibody GWASs.** The SARS-CoV-2 results from the COVID-19 Host Genetics Initiative are shown at the top table, followed by the results for these seven SNPs in ALSPAC. Given multiple testing $P$ < 5.1×10$^{-4}$ was considered evidence for association in ALSPAC. Abbreviations: Chr, Chromosome; Beta, Effect estimate for the effect allele; L95, Lower 95% confidence interval; U95, Upper 95% confidence interval; P, *p*-value. (-) denotes SNPs that were not present in the reference panel or removed due to quality control filtering.

**Table S6. Comparison of HLA imputation methods at a 2-digit resolution using SNP2HLA and HLA*IMP:03 in ALSPAC.** Abbreviations: N, number of individuals; %, percentage of concordance or discordance.

**Table S7. Comparison of HLA imputation methods at a 4-digit resolution using SNP2HLA and HLA*IMP:03 in ALSPAC.** Abbreviations: N, number of individuals; %, percentage of concordance or discordance.

**Table S8. Allele frequency comparison of HLA:IMP*03 and SNP2HLA at a 2-digit resolution with Type 1 Diabetes Genetic Consortium HLA reference panel.** For each HLA allele at a 2-digit resolution, the allele frequency calculated using HLA:IMP*03 (left column) and SNP2HLA (middle column) are compared with Type 1 Diabetes Genetic Consortium HLA reference panel (right column). Abbreviations: T1DGC, Type 1 Diabetes Genetic Consortium HLA reference panel.

**Table S9. Allele frequency comparison of HLA:IMP*03 and SNP2HLA at a 4-digit resolution with Type 1 Diabetes Genetic Consortium HLA reference panel.** For each HLA allele at a 4-digit resolution, the allele frequency calculated using HLA:IMP*03 (left column) and SNP2HLA (middle column) are compared with Type 1 Diabetes Genetic Consortium HLA reference panel (right column). Abbreviations: T1DGC, Type 1 Diabetes Genetic Consortium HLA reference panel.

**Table S10. Comparison of association results at a 2-digit resolution between SNP2HLA and HLA*IMP:03 of ALSPAC antibodies at the seven-year clinic.** SNPs associated with antibodies against an infection of interest are illustrated, with HLA:IMP*03 results shown in the left-hand column, and SNP2HLA results in the right-hand column. Given multiple testing, $P < 6.2 \times 10^{-4}$ was considered evidence of an association in ALSPAC. Abbreviations: Beta, Effect estimate for the effect allele; L95, Lower 95% confidence interval; U95, Upper 95% confidence interval; P, *p*-value.

**Table S11. Comparison of association results at a 4-digit resolution between SNP2HLA and HLA*IMP:03 of ALSPAC antibodies at the seven-year clinic.** SNPs associated with antibodies against an infection of interest are illustrated, with HLA:IMP*03 results shown in the left-hand column, and SNP2HLA results in the right-hand column. Given multiple testing, $P < 5.0 \times 10^{-4}$ was considered evidence of an association in ALSPAC. Abbreviations: Beta, Effect estimate for the effect allele; L95, Lower 95% confidence interval; U95, Upper 95% confidence interval; P, *p*-value.

**Table S12. Discovery genome-wide HLA 2-digit association analyses of antibodies using UK Biobank.** Abbreviations: Beta, Effect estimate for the presence allele; L95, Lower 95% confidence interval; U95, Upper 95% confidence interval; P, *p*-value.

**Table S13. Discovery genome-wide HLA 4-digit association analyses of antibodies using UK Biobank.** Abbreviations: Beta, Effect estimate for the presence allele; L95, Lower 95% confidence interval; U95, Upper 95% confidence interval; P, *p*-value.

**Table S14. HLA alleles at a two-digit resolution ($P < 7.1{\times}10^{-5}$) and at a 4-digit resolution ($P < 2.8{\times}10^{-5}$) associated with antibodies measured in discovery cohort, UK Biobank, and replication performed in ALSPAC ($P < 0.003$).** In the discovery cohort UK Biobank, SNPs exceeding the *P*-value threshold after adjusting for multiple testing are shown in the left-hand column. Replication of these SNPs in ALSPAC using the methods HLA:IMP*03 and SNP2HLA are shown in the middle and right-hand column, respectively. Abbreviations: Beta, Effect estimate for the presence allele; L95, Lower 95% confidence interval; U95, Upper 95% confidence interval; P, *p*-value; NA, No HLA association; (-) HLA alleles not present in the reference panel.
